# Large Language Models Fail to Reproduce Level I Recommendations for Breast Radiotherapy

**DOI:** 10.1101/2024.11.11.24317123

**Authors:** Kathleen Tang, John Han, Shengyang Wu

## Abstract

This study evaluates the reliability of the largest public-facing large language models in providing accurate breast cancer radiotherapy recommendations. We assessed ChatGPT 3.5, ChatGPT 4, ChatGPT 4o, Claude 3.5 Sonnet, and ChatGPT o1 in three common clinical scenarios. The clinical cases are as follows: post-lumpectomy radiotherapy in a 40 year old woman, (2) postmastectomy radiation in a 40 year old woman with 4+ lymph nodes, and (3) postmastectomy radiation in an 80 year old woman with early stage tumor and negative axillary dissection. Each case was designed to be unambiguous with respect to the Level I evidence and clinical guideline-supported approach. The evidence-supported radiation treatments are as follows: (1) Whole breast with boost (2) Regional nodal irradiation (3) Omission of post-operative radiotherapy. Each prompt is presented to each LLM multiple times to ensure reproducibility. Results indicate that the free, public-facing models often fail to provide accurate treatment recommendations, particularly when omission of radiotherapy was the correct course of action. Many recommendations suggested by the LLMs increase morbidity and mortality in patients. Models only accessible through paid subscription (ChatGPT o1 and o1-mini) demonstrated greatly improved accuracy. Some prompt-engineering techniques, rewording and chain-of-reasoning, enhanced the accuracy of the LLMs, while true/false questioning significantly worsened results. While public-facing LLMs show potential for medical applications, their current reliability is unsuitable for clinical decision-making.

## Introduction

Earlier studies have demonstrated large language model’s (LLM) proficiency in successfully navigating a range of standardized examinations in the domain of radiation oncology. Notably, ChatGPT has attained passing marks in radiation physics board exams and the ACR Radiation Oncology in-training exam. [1,2] Additionally, it has offered expert-level medical insights for CNS tumor boards and undergone trials for assisting in chemotherapy selection for solid tumors. [3,4] A distinctive feature of LLM is the ability to provide rationale for its responses and to modify its answers through different user inputs, such as chain of reasoning and rewording prompts. These abilities have generated optimism about the potential incorporation of large language models, such as ChatGPT, into daily clinical decision-making in radiation oncology.

Radiotherapy is among the most evidence-based therapeutic practices in modern medicine. Within radiotherapy, the practice of post-operative breast radiation has been investigated in over dozens of large randomized clinical trials. [5] Consequently, radiation therapy for postoperative breast cancer has been standardized through the adoption of international guidelines by diverse oncology groups including breast surgeons, radiation physicians, and medical oncologists. [6,7]

In this report, we evaluate the accuracy and reliability of the largest public facing large language models, ChatGPT (OpenAI GPT 3.5, June 2023; Open AI GPT 4, August 2024; Open AI GPT 4o, August 2024) and Sonnet (Anthropic Claude 3.5, August 2024) in three common clinical scenarios in breast cancer radiotherapy.

## Methods

We evaluated 3 different prompts presenting standard radiation scenarios (1) Post lumpectomy and sentinel LN dissection in a 40 year old woman early stage IA hormone positive ductal cancer without lymph node involvement. (2) Post mastectomy and axillary LN dissection in a 40 year old woman with stage III hormone positive breast cancer and five lymph nodes (3) Post mastectomy radiation in a 80 year old woman with axillary lymph node dissection with hormone positive breast cancer, no involvement of lymph nodes, negative margins. [Figure 1] These were designed to be in line with level I evidence so that the clear choice for radiation treatment would not be ambiguous. The vast majority of radiation oncology physicians would provide whole breast radiation with boost/conedown to the first woman, post-mastectomy regional nodal irradiation to the second woman, and render no radiation to the final woman.

**Figure 1:**
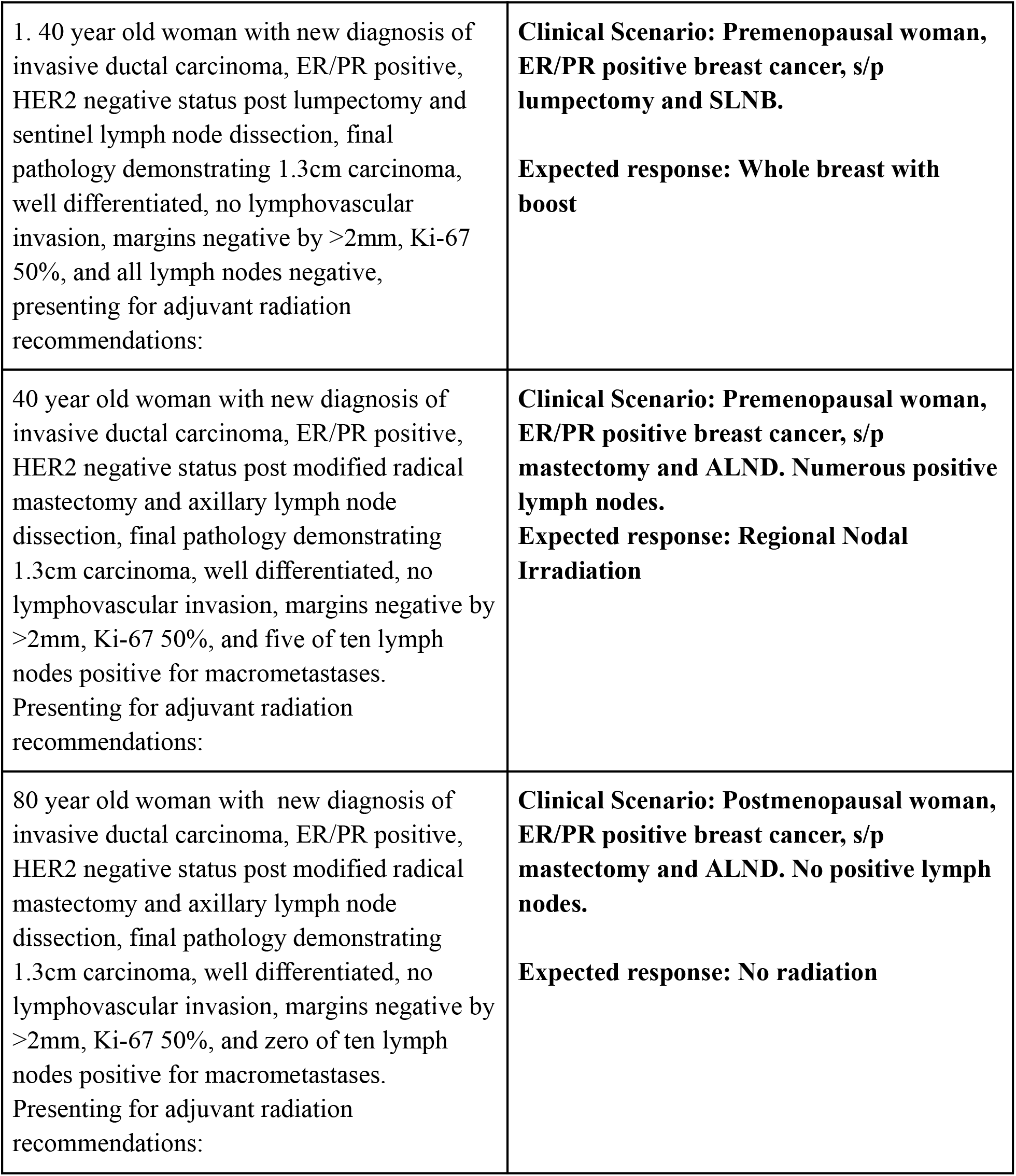
Three Clinical Cases and Respective Category I Evidence-Based Recommendations.

Each prompt was inputted into the LLM (GPT 3.5, GPT 4, GPT 4o, Sonnet 3.5) four-five times and the responses were recorded. A previous study suggested that prompts entered under one chat session could lead to issues with duplicated inquiries to the LLM (Schulte 2023). As a result, our study chose to place each prompt and their duplicate prompts in separate chat sessions to prevent influence of data from different prompts.

The responses were read by researchers and graded as accurate, partially accurate, or inaccurate. If the response did not mention at all the standard of care treatment, it was graded as inaccurate. If the response reproduced the standard care in entirety, it was marked as accurate. Any other responses were marked as partially accurate.

Three previously validated user prompt modifications (rewording the prompt, chain of thought reasoning, true or false) were made to the prompt without changing the overall clinical scenario in order to improve the accuracy of the response. The responses to the modified prompts were again graded on accuracy in the same way (i.e. accurate, partially accurate, and inaccurate).

## Results

Each prompt was inputted four to five times into the LLM. For the first prompt, GPT 3.5’s generated response was accurate 25%, partially accurate 50%, and inaccurate 25%. Accurate responses returned mention of whole breast radiation with boost, which was in line with clinical guidelines and reflected in the widely accepted standard of care. Inaccurate responses were characterized by a lack of any mention of radiotherapy. Partially accurate responses contained radiation as a component of adjuvant care, however named options such as partial breast or failed to mention the inclusion of a radiation boost. The second prompt was accurate 0%, partially accurate 100%, and inaccurate 0%. Partially accurate responses mentioned adjuvant radiation therapy. However, GPT 3.5 failed to mention regional nodal irradiation. The final prompt was accurate 0%, partially accurate 0%, and inaccurate 100%. Inaccurate responses all included mention of adjuvant radiation therapy [Table 1].

**Table 1:**
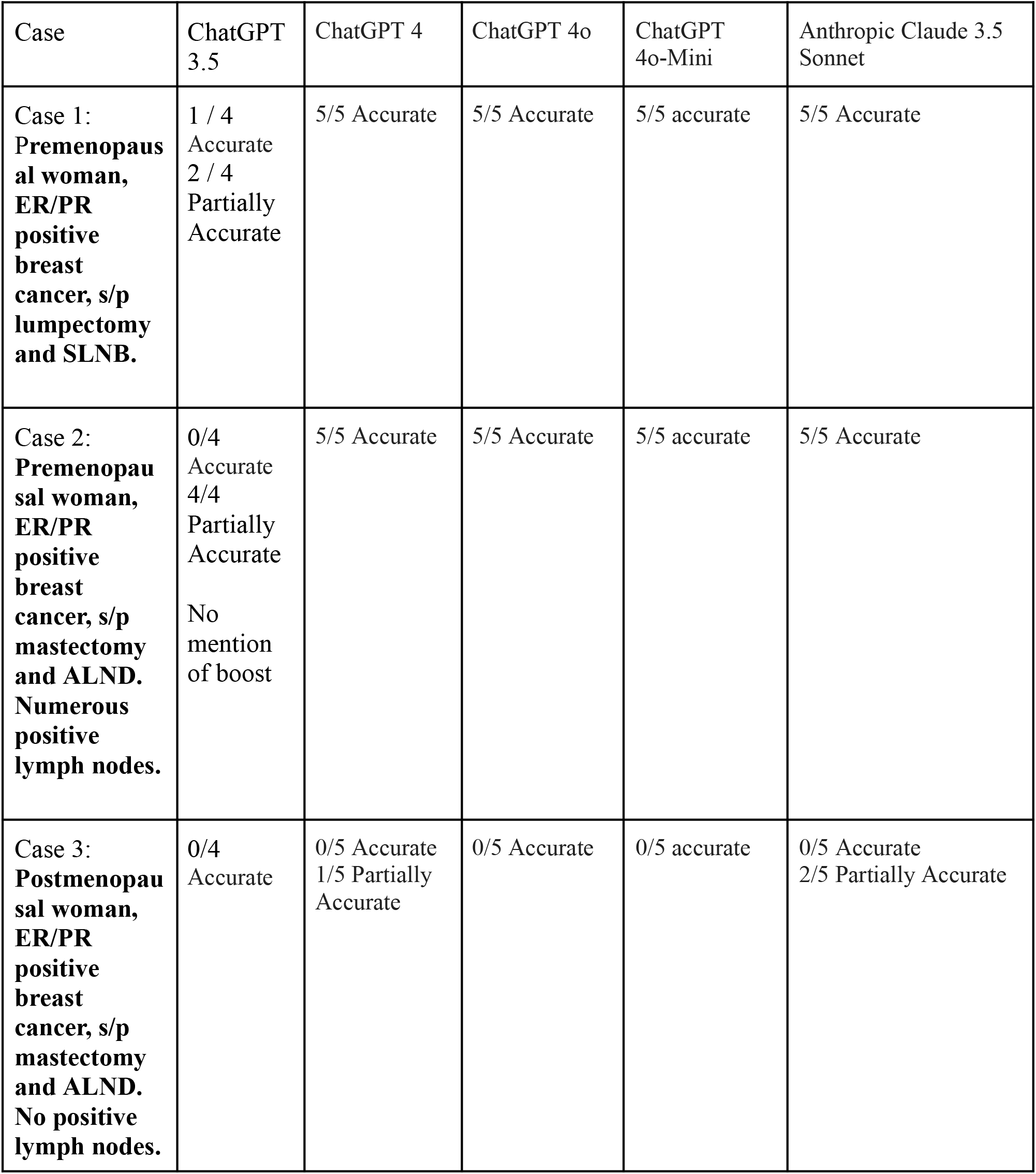
Accuracy of Response to Prompts.

| Case | ChatGPT 3.5 | ChatGPT 4 | ChatGPT 4o | ChatGPT 4o-Mini | Anthropic Claude 3.5 Sonnet |
| --- | --- | --- | --- | --- | --- |
| Case 1:<br>**Premenopausal woman, ER/PR positive breast cancer, s/p lumpectomy and SLNB.** | 1 / 4<br>Accurate<br>2 / 4<br>Partially<br>Accurate | 5/5 Accurate | 5/5 Accurate | 5/5 accurate | 5/5 Accurate |
| Case 2:<br>**Premenopausal woman, ER/PR positive breast cancer, s/p mastectomy and ALND. Numerous positive lymph nodes.** | 0/4<br>Accurate<br>4/4<br>Partially<br>Accurate<br>No<br>mention<br>of boost | 5/5 Accurate | 5/5 Accurate | 5/5 accurate | 5/5 Accurate |
| Case 3:<br>**Postmenopausal woman, ER/PR positive breast cancer, s/p mastectomy and ALND. No positive lymph nodes.** | 0/4<br>Accurate | 0/5 Accurate<br>1/5 Partially<br>Accurate | 0/5 Accurate | 0/5 accurate | 0/5 Accurate<br>2/5 Partially Accurate |

Each prompt was reworded to include the phrase “specific” treatment or recommendations. The first prompt was biased towards the accurate answer of “WBRT with boost” by including terms such as “additional radiation” and “along with boost radiation.” Ten iterations of prompt rewording were performed, yielding answers that were accurate (1/10) 10% of the time, partially accurate (9/10) 90%, and inaccurate (0/10) 0%. For partially accurate answers, GPT 3.5 did not specify boost radiation was recommended or its response specifically stated radiation boost was not required. The second prompt was also reworded similarly. The LLM was accurate 0%, partially accurate (3/3) 100%, and inaccurate 0%. Partially accurate answers were characterized by GPT 3.5 stating that external beam radiation therapy (EBRT) would be the appropriate treatment for the patient but did not mention regional nodal irradiation. The rewording of the third prompt was accurate 0%, partially accurate 0%, and (6/6) 100% inaccurate. All responses mention use of radiation when no radiation should be recommended. [Table 2]

**Table 2:** Rewording the Question - ChatGPT 3.5.

|  | Accurate | Partially Accurate | Inaccurate |
| --- | --- | --- | --- |
| Prompt 1: Reworded as<br>“what specific kind of adjuvant radiation therapy and additional radiation should be done for ...”<br>“What kind of treatment should be provided for a ...”<br>“What specific type of adjuvant radiation therapy should a ... receive and should there be any additional procedures done”<br>“What specific type of adjuvant radiation therapy should a ... receive and along with boost radiation” | 1/10<br>“Whole breast radiation (WBRT) ... and additional radiation boost” | 9/10<br>“Whole breast radiation therapy (WBRT) with or without a radiation boost”<br>“Whole breast radiation therapy (WBRT) ... [and] no additional radiation boost”<br>“WBRT ... may not be any additional procedures necessary”<br>“Adjuvant radiation therapy” | 0/10 |
| Prompt 2: Reworded as<br>“What kind of treatment should a ... do” | 0/3 | 3/3<br>“Adjuvant radiation therapy ... systemic therapy ... hormonal therapy ... | 0/3 |
| “what specific kind of treatment should a 40 year old woman with new diagnosis of ... presenting for adjuvant radiation recommendations receive” |  | chemotherapy”<br>“External beam radiation therapy (EBRT)” |  |
| Prompt 3: Rewording as<br>“what specific kind of treatment should be provided for ...”<br>“what would nccn category 1 specifically recommend for ...” | 0/6 | 0/6 | 6/6<br>All mention postoperative radiation therapy |

Then, a true or false strategy was implemented. This particular strategy was graded as being fully accurate or fully inaccurate due to the model only being able to choose true or false. For the first prompt, the results showed a (2/2) 100% inaccuracy rate in which GPT 3.5 responded that boost radiation was not needed. For the second prompt, the true false strategy resulted in a (8/15) 53% accuracy rate in which the model responded true and a (7/15) 47% inaccuracy rate in which the model responded with false. Prompt 3 yielded a (2/3) 67% accuracy rate and a (1/3) 33% inaccuracy rate when asked if a patient should receive radiation. [Table 3]

**Table 3:** True or False Statements - ChatGPT 3.5.

|  | Accurate | Inaccurate |
| --- | --- | --- |
| Prompt 1: True or False<br>“true or false: a 40 year old woman with new diagnosis of invasive ductal carcinoma, ER/PR positive, HER2 negative status post lumpectomy and sentinel lymph node dissection, final pathology demonstrating 1.3cm carcinoma, well differentiated, no lymphovascular invasion, margins negative by >2mm, Ki-67 50%, and all lymph nodes negative, presenting for | 0/2 | 2/2<br>“False ... does not require boost radiation treatment in addition to ... WBRT” |
| adjuvant radiation recommendations should receive wbrt with boost radiation” |  |  |
| Prompt 2: True or False<br>“True or False, a 40 year old woman ... requires regional nodal irradiation”<br>“Yes or no: ...” | 8/15<br>“True ... regional nodal irradiation (RNI) should be considered ...”<br>“It is reasonable to consider regional nodal irradiation (RNI).”<br>“Yes.” | 7/15<br>“False ... the use of regional nodal irradiation (RNI) is not typically recommended.”<br>“ ... based on the information provided, it is more appropriate to recommend external beam radiation therapy (EBRT) ... rather than RNI.”<br>“No.” |
| Prompt 3: True or False.<br>“True or False, a 80 year old woman ... does not require radiation” | 2/3<br>“True. Based on the provided information, an 80-year-old woman with the described characteristics may not necessarily require adjuvant radiation therapy.” | 1/3<br>“False. The statement that an 80-year old woman with the described characteristics should receive no radiation is not accurate.” |

A chain of reasoning prompting strategy was then tested. This strategy was used with the intention of using the model’s ability to remember past conversations to our advantage, as well as utilize its reasoning capabilities to evaluate its response.

The chain of reasoning for prompt 1 consisted of asking the LLM for factors that would lead to a recommendation of WBRT and boost as the treatment. Then, the data set of the patient would be inputted into the chat session and the LLM was asked what it would recommend for the patient based on the response it gave. If ChatGPT 3.5 gave an answer that was accurate and then switched to one that was partially accurate or inaccurate, it would be asked to explain its reasoning. This method resulted in the LLM being accurate (4/5) 80%, partially accurate (1/5) 20%, and inaccurate 0%. In the partially accurate instance, the LLM stated that it was unsure if a radiation boost would be needed.

GPT 3.5 was then asked about the factors that would lead to a recommendation of regional nodal irradiation as the treatment option. The second prompt was then presented. This strategy yielded a (2/2) 100% partial accuracy rate in which the model stated EBRT as the correct treatment option, but with no mention of regional nodal irradiation. When asked to explain its reasoning, the model stated that regional nodal irradiation was not a common treatment option for early-stage breast cancer cases.

For the final prompt, GPT 3.5 was asked about factors that would lead it to recommend no radiation. Then two choices were given, one being the correct answer of “no radiation” and then “hormone therapy.” A clarification or explanation question would be asked after every incorrect or partially correct answer. Out of five correct responses, only once did the LLM answer that both treatments would be recommended. The other four responses correctly answered no radiation. This resulted in a (5/7) 71% accuracy rate, (2/7) 29% partially accurate rate in which the LLM would choose hormone therapy as being the only correct treatment. There were no inaccurate responses as the LLM did not suggest radiation treatment when prompted this way. [Table 4]

**Table 4:** Chain of Reasoning Strategy - ChatGPT 3.5.

|  | Accurate | Partially Accurate | Inaccurate |
| --- | --- | --- | --- |
| Prompt 1: Chain of reasoning<br>Asked for factors that would lead to a recommendation of WBRT and boost. Then LLM was asked what it would recommend for the patient based on its response. | 4/5<br>“WBRT ... [and] in some cases, a boost radiation treatment may be recommended after WBRT”<br>“Whole-breast radiation therapy (WBRT) with a boost radiation can be | 1/5<br>“WBRT ... the need for an additional boost of radiation may not be indicated.” | 0/5 |
| If it gives the wrong answer, it is asked to explain its reasoning. | considered” |  |  |
| Prompt 2: Chain of reasoning<br>Asked for factors that would lead to a recommendation of regional nodal irradiation. Then, a multiple choice question was posed to the LLM.<br>If it gives the wrong answer, it is asked to explain its reasoning. | 0/2 | 2/2<br>“External beam radiation therapy (EBRT) ... regional nodal irradiation (RNI) is not typically recommended in all cases of early-stage breast cancer.” | 0/2 |
| Prompt 3: Chain of reasoning<br>Asked a multiple choice question to the LLM. Asked for clarification if it got the answer wrong. | 5/7<br>“Both options (a) no radiation and (b) hormone therapy, may be appropriate treatment options ...”<br>“Option (a) no radiation” | 2/7<br>“Would likely involve the following options<br>a) No radiation ...<br>b) Hormone therapy<br>“The correct answer would likely be option (b) hormone therapy.” | 0/7 |

The same strategy was used for the other LLMs. GPT 4 answered with (5/5) 100/% accuracy to both prompt 1 and prompt 2. However, similarly to GPT 3.5, it was unable to accurately answer the third prompt. With (0/5) 0% accurate, (1/5) 20% partially accurate, (4/5) 80% inaccurate, it showed a slight accuracy improvement - the instance where the answer was deemed partially accurate, GPT 4 suggested omitting radiation as a possibility but failed to take a definitive stance.

Accuracy improved notably after alternative prompting. Including the term “specific kind of treatment” resulted in (1/5) 20% accurate responses and (4/5) 80% partially accurate responses. Asking “what would NCCN category I recommend” to GPT 4 resulted in (5/5) 100% accurate responses. True or false prompting resulted in (0/5) 0% accurate and (5/5) inaccurate responses. Our chain of reasoning (CoR) prompting resulted in (5/5) 100% accuracy, and in two instances the model actually answered correctly before CoR prompting was used, indicating inconsistencies in GPT 4’s accuracy.

Just like GPT 4, GPT 4o answered prompt 1 and prompt 2 with (5/5) 100% accuracy. Prompt 3 resulted in (0/5) 0% accurate responses and (5/5) inaccurate responses. In contrast with GPT 4, responses leaned more towards omitting adjuvant radiation; however, the model still falsely considered other radiotherapy options.

Claude 3.5 Sonnet answered with (5/5) 100% accuracy to both prompt 1 and prompt 2. Once again, however, the model was unable to consistently answer prompt 3 with accuracy: 3.5 Sonnet attained (0/5) accurate responses, (2/5) partially accurate responses, and (3/5) inaccurate responses.

GPT 4o-mini, the most accessible public free model as of Fall 2024, answered prompt 1 and prompt 2 with (5/5) 100% accuracy. For prompt 3, the model responded with (0/5) 0% accurate responses and (5/5) inaccurate responses.

As of the time of writing this paper, a new line of ChatGPT models is being developed and is accessible by paying members only: GPT o1-preview and o1-mini. The new models are designed to spend much more time thinking before responding, working through built-in chains of reasoning to produce carefully-considered responses. Both GPT o1-preview and o1-mini answered with 100% accuracy to all three prompts, and even attempts to persuade the two new models that they were incorrect in their responses to prompt 3 were futile.

## Discussion

Previous studies have demonstrated ChatGPT’s inconsistencies in generating responses to queries with objectively evaluable responses, including hallucinations and lack of evidential support, thus raising concerns about large language models’ reliability and potential for harmful treatment suggestions in clinical settings. [8,9] Others believe that ChatGPT has the potential to assist in medicine but its current functionality may be best suited as a tool for medical education like previous data-driven efforts in natural language processing. [10,11] Our current study adds to these findings. We find that none of our tested large language models except GPT o1-preview and GPT o1-mini consistently produce the accurate treatment recommendations, and cannot be trivially modified to improve accuracy, calling attention to issues with clinical LLM applications at present.

Completely accurate statements about breast radiotherapy were not improved despite attempts made to modify the prompts. In fact, when presented with a true or false statement, model performance was consistently worse. It is unclear why true and false prompt modification would worsen the performance of LLMs - but this points to the outsize influence of prompt modifications to the output of the LLM.

We found that one strategy to decrease inaccuracy in responses is chain of reasoning (CoR) prompting. Within this work, CoR prompting involved priming an LLM with the correct recommendation for a general clinical scenario, and oftentimes coercing the LLM to limit its responses with multiple choice questions that do not contain the inaccurate answer. This strategy requires a skilled operator with prior knowledge of the correct answer to guide the LLM. At present, it does not appear to be suitable for clinical workflow.

This study has several limitations. Large language models are always getting updated and refined, with new models boasting training set sizes and various enhancements likely to improve accuracy. Even still, a group of medical physicists working with a majority were able to outperform GPT4. [1] Furthermore, specialized LLMs like MedPalm-E or other fine-tuned domain-specific biology models hold potential for performance improvement. [12] Despite the growing training data and fine-tuning, the identified phenomena of stochastic responses to deterministic prompts (i.e. hallucinations and falsifications) as well as the potential to improve or corrupt responses with prompt modifications are inherent LLM traits unaffected by training data size. Future investigations in clinical applications of LLM might concentrate on devising LLMs with deterministic outputs. In addition, further research may focus on understanding which prompt additions impact prompt accuracy.

## Conclusion

Large language models are unable to consistently reproduce level I recommendations for standard breast cancer radiotherapy, especially when having to choose to omit radiation. True or false prompting strategies worsened the accuracy of responses. Incorporating chain of reasoning prompting improved response accuracy. However, even with modifications, responses often only achieved partial accuracy and oftentimes omitted key aspects of treatment.

## Supporting information

ChatGPT 3.5 Supplemental Analysis

All Model Supplemental Data

## Data Availability

All data produced in the present work are contained in the manuscript

## References

1. Holmes, J., Liu, Z., Zhang, L., Ding, Y., Sio, T. T., McGee, L. A., … & Liu, W. (2023). Evaluating large language models on a highly-specialized topic, radiation oncology physics. Front Onc, (15)2023

2. Huang, Y., Gomaa, A., Weissmann, T., Grigo, J., Tkhayat, H. B., Frey, B., … & Putz, F. (2023). Benchmarking chatgpt-4 on acr radiation oncology in-training exam (txit): Potentials and challenges for ai-assisted medical education and decision making in radiation oncology. Front. Onc. (13)2023

3. Haemmerli, J., Sveikata, L., Nouri, A., May, A., Egervari, K., Freyschlag, C., … & Bijlenga, P. (2023). ChatGPT in glioma adjuvant therapy decision making: ready to assume the role of a doctor in the tumour board?. BMJ Health & Care Informatics, 30(1).

4. Schulte, B. (2023). Capacity of ChatGPT to Identify Guideline-Based Treatments for Advanced Solid Tumors. Cureus, 15(4).

5. Early Breast Cancer Trialists’ Collaborative Group. (1995). Effects of radiotherapy and surgery in early breast cancer—an overview of the randomized trials. New England Journal of Medicine, 333(22), 1444–1456.

6. Smith, Benjamin D., et al. “Radiation therapy for the whole breast: Executive summary of an American Society for Radiation Oncology (ASTRO) evidence-based guideline.” Practical radiation oncology 8.3 (2018): 145–152.

7. Recht, A., Edge, S. B., Solin, L. J., Robinson, D. S., Estabrook, A., Fine, R. E., … & Pfister, D. G. (2001). Postmastectomy radiotherapy: clinical practice guidelines of the American Society of Clinical Oncology. Journal of Clinical Oncology, 19(5), 1539–1569.

8. Alkaissi, Hussam, and Samy I. McFarlane. “Artificial hallucinations in ChatGPT: implications in scientific writing.” Cureus 15.2 (2023).

9. Kassab, J., Nasr, L., Gebrael, G., Chedid El Helou, M., Saba, L., Haroun, E., … & Nasr, F. (2023). AI-based online chat and the future of oncology care: a promising technology or a solution in search of a problem?. Frontiers in Oncology, 13, 1176617.

10. Ebrahimi, B., Howard, A., Carlson, D. J., & Al-Hallaq, H. (2023). ChatGPT: Can a Natural Language Processing Tool Be Trusted for Radiation Oncology Use?. International Journal of Radiation Oncology, Biology, Physics.

11. Wu, S., Langlotz, C. P., Lakhani, P., & Ungar, L. H. (2012). Extracting templates from radiology reports using sequence alignment. International journal of data mining and bioinformatics, 6(6), 633–650.

12. Singhal, K., Azizi, S., Tu, T., Mahdavi, S. S., Wei, J., Chung, H. W., … & Natarajan, V. (2023). Large language models encode clinical knowledge. Nature, 1–9.

