## Supplementary material for "Large Language Models Fail to Reproduce Level I Recommendations for Breast Radiotherapy": All Model Supplemental Data

Details

1) Always get the answer right - ie. ask it in a way that the answer is always in line with NCCN category 1 recommendations
2) Always gets the answer wrong - i.e. ask it in a way that the answer is always wrong.
3) Engineering neutral - Patient data inputted into the machine

- Provided a list of "classical cases" for each category 1 recommendation, and changed the wording to achieve (1) and (2).
- 1000-1500 brief report

ChatGPT Chat Practice Example-
https://chat.openai.com/share/b3ecdf56-09f3-4223-8250-a733ca9db259

First Example - https://chat.openai.com/share/aac0ee57-c0ea-4b2b-9ba7-1afba33405e0

Second Example - https://chat.openai.com/share/f6a95c5f-4930-4a0f-ba1f-dcf38d2342bf

Third Example -https://chat.openai.com/share/af99b940-1469-4fb6-b84c-fbd95a82f092

**Questions/Things I Find Interesting**

- Were some of these correct treatments from the NCCN listed before or after September 2021?
- If you give the AI a very limited question or very limited options, it will be more likely to narrow in on the right answer.

**Practice Example**

A typical Stage IA premenopausal patient
40 year old woman with new diagnosis of invasive ductal carcinoma, ER/PR positive, HER2 negative status post lumpectomy final pathology demonstrating 1.3cm carcinoma, well differentiated, no lymphovascular invasion, margins negative by >2mm, Ki-67 50%, presenting for adjuvant radiation recommendations

- Put the prompt unedited 4-5 times into chatGPT and record the response.

General Observations

- The more times the AI is asked, the number of common considerations decrease
- Consistent considerations are "tumor characteristics and "lumpectomy"
- Age and overall health appears only once
- Mentions adjuvant radiation therapy

| | |
|---|---|
| 1 | I'm not a doctor, but I can provide some general information regarding adjuvant radiation recommendations for invasive ductal carcinoma. It's important to consult with an oncologist who can evaluate your specific case and provide personalized advice. However, here are some common considerations for adjuvant radiation therapy in cases similar to yours:<br><br>1. Tumor Characteristics: In your case, the invasive ductal carcinoma is ER/PR positive and HER2 negative. These factors can influence treatment decisions, and your oncologist will consider them when determining the need for adjuvant radiation therapy.<br>2. Lumpectomy: Since you mentioned a lumpectomy, it suggests that breast-conserving surgery was performed rather than a mastectomy. Adjuvant radiation is typically recommended after lumpectomy to reduce the risk of local recurrence.<br>3. Final Pathology: The size of the carcinoma, well differentiation, absence of lymphovascular invasion, and negative margins (>2mm) are positive factors that generally indicate a lower risk of recurrence. These factors may affect the decision-making process regarding adjuvant radiation therapy.<br>4. Ki-67: Ki-67 is a marker of cellular proliferation, and a higher percentage (such as 50% in your case) indicates a more aggressive tumor. This may influence the recommendation for adjuvant radiation therapy, as it suggests a higher risk of recurrence.<br><br>Based on these factors, your oncologist will assess your overall risk of recurrence and consider various treatment options. Adjuvant radiation therapy is commonly recommended for patients who undergo breast-conserving surgery, particularly if they have certain high-risk features. The radiation treatment aims to target any remaining cancer cells in the breast tissue and decrease the likelihood of local recurrence.<br><br>It's essential to discuss your specific case and treatment options with your oncologist, as they will consider additional factors, such as your overall health, medical history, and any additional treatment plans (e.g., hormonal therapy), to make a comprehensive recommendation for your situation. |

| | |
|---|---|
| 2 | Based on the information provided, here are some possible recommendations for adjuvant radiation therapy for your case:<br><br>1. Age and Overall Health: Your age of 40 may be a relevant factor in treatment decisions. Younger patients sometimes have a higher risk of recurrence, and adjuvant radiation therapy may be considered more strongly in these cases. However, other factors, such as overall health and individual preferences, will also be taken into account.<br>2. Tumor Characteristics: Your tumor is ER/PR positive, HER2 negative, and well-differentiated. These are generally favorable characteristics associated with a lower risk of recurrence. However, the high Ki-67 index of 50% suggests a higher proliferation rate, which could impact treatment decisions. The oncologist will evaluate the overall risk based on these factors.<br>3. Lumpectomy and Negative Margins: Since you had a lumpectomy and the margins are negative by more than 2mm, this indicates that the surgical removal of the tumor was successful. However, adjuvant radiation therapy is typically recommended after lumpectomy to reduce the risk of local recurrence.<br>4. Ki-67: The high Ki-67 index of 50% suggests a higher risk of tumor growth and cell proliferation. This factor may further support the consideration of adjuvant radiation therapy to target any potentially remaining cancer cells.<br><br>Based on these factors, it is likely that your oncologist will recommend adjuvant radiation therapy following your lumpectomy. Radiation therapy is typically delivered to the remaining breast tissue to reduce the risk of local recurrence. However, treatment decisions are individualized, and it is crucial to discuss all options with your healthcare team to make an informed decision. They will consider your specific case, including your overall health, potential side effects of radiation, and any other planned treatments (e.g., hormonal therapy), to create a comprehensive treatment plan tailored to your needs. |

| | |
|---|---|
| 3 | Based on the information provided, here are some possible recommendations for adjuvant radiation therapy for your case:<br><br>1. Tumor Characteristics: Your tumor is ER/PR positive and HER2 negative, which is generally associated with a better prognosis. The fact that it is well differentiated and has no lymphovascular invasion is also positive. However, the high Ki-67 index of 50% suggests a higher proliferation rate, which could indicate a higher risk of recurrence.<br>2. Lumpectomy and Negative Margins: Your lumpectomy was successful, with negative margins by more than 2mm. This means that the tumor was completely removed, reducing the risk of local recurrence. Adjuvant radiation therapy is typically recommended after lumpectomy to further reduce this risk.<br><br>Based on these factors, adjuvant radiation therapy is likely to be recommended in your case. The purpose of radiation therapy is to target any remaining cancer cells in the breast tissue and reduce the risk of local recurrence. The decision will be made based on a combination of factors, including the specific characteristics of your tumor, your overall health, and individual considerations.<br><br>It's important to discuss your case with your oncologist who will evaluate all the relevant factors and provide personalized recommendations based on your specific situation. They will consider the potential benefits of radiation therapy in reducing the risk of recurrence compared to any potential side effects or risks associated with the treatment. They will also take into account any additional treatments, such as hormonal therapy, that may be recommended as part of your overall treatment plan. |

| | |
|---|---|
| 4 | Based on the information provided, here are some possible recommendations for adjuvant radiation therapy for your case:<br><br>1. Tumor Characteristics: Your tumor is ER/PR positive, HER2 negative, and well differentiated. These characteristics indicate a lower risk of recurrence compared to tumors that are ER/PR negative, HER2 positive, or poorly differentiated. However, the high Ki-67 index of 50% suggests a higher proliferation rate, which could indicate a higher risk of recurrence. This factor will be considered in the overall treatment decision.<br>2. Lumpectomy and Negative Margins: The lumpectomy successfully removed the tumor, and the negative margins (>2mm) indicate that no cancer cells were found at the edges of the excised tissue. However, adjuvant radiation therapy is typically recommended after lumpectomy to reduce the risk of local recurrence.<br><br>Based on these factors, your oncologist is likely to recommend adjuvant radiation therapy in your case. The purpose of radiation therapy is to target any remaining cancer cells in the breast tissue and reduce the risk of local recurrence. The decision will be made based on a combination of factors, including the specific characteristics of your tumor, your overall health, and individual considerations.<br><br>It's important to have a detailed discussion with your oncologist to understand the potential benefits and risks of radiation therapy in your situation. They will consider the potential benefits in reducing the risk of recurrence compared to any potential side effects or long-term risks associated with the treatment. They will also take into account your preferences and any additional treatments, such as hormonal therapy, that may be recommended as part of your overall treatment plan. |

| | |
|---|---|
| 5 | Based on the provided information, here are some possible recommendations for adjuvant radiation therapy for your case:<br><br>1. Tumor Characteristics: Your tumor is ER/PR positive and HER2 negative, which is generally associated with a better prognosis. The fact that it is well differentiated and has no lymphovascular invasion is also positive. However, the high Ki-67 index of 50% indicates a higher proliferation rate, suggesting a potentially increased risk of recurrence.<br>2. Lumpectomy and Negative Margins: Your lumpectomy successfully removed the tumor, and the negative margins (>2mm) indicate that no cancer cells were found at the edges of the excised tissue. However, adjuvant radiation therapy is typically recommended after lumpectomy to further reduce the risk of local recurrence.<br><br>Given the characteristics of your tumor and the guidelines for adjuvant radiation therapy, it is likely that your oncologist will recommend radiation treatment. The purpose of radiation therapy is to target any remaining cancer cells in the breast and reduce the risk of local recurrence.<br><br>It is important to have a detailed discussion with your healthcare team, including your oncologist and radiation oncologist, to evaluate the specific risks and benefits in your case. They will consider factors such as your overall health, personal preferences, and any additional treatments (e.g., hormonal therapy) to create an individualized treatment plan.<br><br>Remember to ask any questions you may have to fully understand the recommended treatment and its potential side effects. Your healthcare team is best equipped to provide personalized recommendations based on your unique circumstances. |

- Respond with the correct answer - which is whole breast radiation with boost. Repeat it about 4-5 times to make sure it always gets it correct.

General Observations

- During my 3rd phrasing of the question, the correct answer appeared during the 3rd, 4th, and 5th repeat
- During the 4th phrasing of the question, the correct answer appeared every time except the 2nd repeat
- During 5th phrasing, the correct answer appears each time. However, there are

additional suggestions that appear in certain repeats.

Phrasings

1) What treatment plan is recommended for a 40 year old woman with new diagnosis of invasive ductal carcinoma, ER/PR positive, HER2 negative status post lumpectomy final pathology demonstrating 1.3cm carcinoma, well differentiated, no lymphovascular invasion, margins negative by >2mm, Ki-67 50%, presenting for adjuvant radiation recommendations?
2) What is necessary to treat a 40 year old woman with new diagnosis of invasive ductal carcinoma, ER/PR positive, HER2 negative status post lumpectomy final pathology demonstrating 1.3cm carcinoma, well differentiated, no lymphovascular invasion, margins negative by >2mm, Ki-67 50%, presenting for adjuvant radiation recommendations
3) What specific kind of adjuvant radiation therapy should be done for 40 year old woman with new diagnosis of invasive ductal carcinoma, ER/PR positive, HER2 negative status post lumpectomy final pathology demonstrating 1.3cm carcinoma, well differentiated, no lymphovascular invasion, margins negative by >2mm, Ki-67 50%, presenting for adjuvant radiation recommendations ↓

3rd phrasing

| | |
|---|---|
| 1 | The specific type of adjuvant radiation therapy that would be recommended for a 40-year-old woman with the described diagnosis is external beam radiation therapy (EBRT). EBRT is the most commonly used form of radiation therapy for breast cancer.<br><br>In this case, after lumpectomy, EBRT would be delivered to the remaining breast tissue to target any potential residual cancer cells and reduce the risk of local recurrence. The radiation oncologist will design a treatment plan that includes determining the appropriate dosage, schedule, and duration of radiation therapy.<br><br>The most common approach for adjuvant radiation therapy is whole-breast radiation therapy (WBRT), where the entire breast is treated with radiation. Typically, WBRT is delivered in daily sessions (Monday to Friday) over a period of approximately 3 to 6 weeks.<br><br>In some cases, additional radiation techniques may be considered, such as accelerated partial breast irradiation (APBI) or intensity-modulated radiation therapy (IMRT). APBI involves delivering radiation specifically to the area around the lumpectomy site, while IMRT uses advanced techniques to precisely target the radiation dose to the tumor bed while sparing surrounding healthy tissues. However, the suitability of these techniques would depend on various factors, including the specific characteristics of the tumor and the patient's individual circumstances.<br><br>It's important to note that the precise details of the radiation therapy plan will be determined by the radiation oncologist based on your specific case. They will consider factors such as the size and location of the tumor, your overall health, and individual considerations to design the most appropriate and effective treatment plan for you. |

| | |
|---|---|
| 2 | Based on the information provided, the most common and appropriate adjuvant radiation therapy for a 40-year-old woman with the described diagnosis would be whole-breast radiation therapy (WBRT). WBRT involves delivering radiation to the entire breast to target any remaining cancer cells and reduce the risk of local recurrence.<br><br>The recommended treatment course for WBRT typically involves daily sessions (Monday to Friday) over a period of approximately 3 to 6 weeks. The specific dosage and schedule will be determined by the radiation oncologist based on your individual case and may be influenced by factors such as the characteristics of the tumor, your overall health, and any additional treatment considerations.<br><br>The goal of WBRT is to provide effective radiation coverage to the entire breast, ensuring that any potential residual cancer cells are adequately treated while minimizing the radiation exposure to surrounding healthy tissues.<br><br>It's important to consult with your radiation oncologist to discuss the specific details of your treatment plan, including the dosage, schedule, and any potential side effects. They will be able to provide personalized care and address any concerns you may have regarding your adjuvant radiation therapy. |

| | |
|---|---|
| 3 | Based on the provided information, the specific kind of adjuvant radiation therapy that is typically recommended for a 40-year-old woman with the described diagnosis is whole-breast radiation therapy (WBRT).<br><br>WBRT involves delivering radiation to the entire breast to target any remaining cancer cells and reduce the risk of local recurrence. The radiation is typically administered in daily sessions (Monday to Friday) over a period of several weeks. The specific dosage and duration of treatment will be determined by a radiation oncologist based on your individual case.<br><br>In addition to WBRT, your radiation oncologist may also consider the use of a boost radiation therapy. A boost is an additional dose of radiation that is targeted to the tumor bed, which is the area where the tumor was removed during the lumpectomy. The boost radiation therapy is typically delivered after the completion of WBRT and may require a shorter treatment duration.<br><br>It's important to note that the precise treatment plan, including the specific dosage, schedule, and duration of radiation therapy, will be determined by your radiation oncologist. They will consider factors such as the size and location of the tumor, the characteristics of the breast tissue, your overall health, and individual considerations to design the most appropriate and effective treatment plan for you.<br><br>Make sure to have a detailed discussion with your radiation oncologist to understand the recommended treatment, potential side effects, and any available alternatives. They will provide personalized recommendations based on your unique circumstances and help guide you through the treatment process. |

| | |
|---|---|
| 4 | Based on the information provided, the specific kind of adjuvant radiation therapy that is commonly recommended for a 40-year-old woman with the described diagnosis is external beam radiation therapy (EBRT). EBRT is the standard treatment approach for adjuvant radiation therapy in breast cancer.<br><br>For your specific case, the most appropriate form of EBRT is likely whole-breast radiation therapy (WBRT). WBRT involves delivering radiation to the entire breast to target any remaining cancer cells and reduce the risk of local recurrence. The radiation is typically given in daily sessions (Monday to Friday) over a period of approximately 3 to 6 weeks. The specific dosage and schedule will be determined by the radiation oncologist based on your individual case and may vary depending on various factors such as the tumor characteristics and your overall health.<br><br>In addition to WBRT, your radiation oncologist may consider a boost radiation therapy. A boost delivers an additional dose of radiation specifically to the tumor bed, which is the area where the tumor was removed during the lumpectomy. The boost is typically given after the completion of WBRT and may involve a shorter treatment duration.<br><br>It's important to note that the precise treatment plan, including the specific dosage, schedule, and duration of radiation therapy, will be determined by your radiation oncologist based on your unique case. They will consider factors such as the size and location of the tumor, the characteristics of the breast tissue, and your individual circumstances to develop the most appropriate and effective treatment plan.<br><br>Be sure to have a detailed discussion with your radiation oncologist to understand the recommended treatment, potential side effects, and any available alternatives. They will provide personalized recommendations based on your specific situation and help guide you through the adjuvant radiation therapy process. |

| | |
|---|---|
| 5 | Based on the information provided, the specific kind of adjuvant radiation therapy that is commonly recommended for a 40-year-old woman with the described diagnosis is whole-breast radiation therapy (WBRT).<br><br>WBRT involves delivering radiation to the entire breast to target any remaining cancer cells and reduce the risk of local recurrence. The radiation is typically administered in daily sessions (Monday to Friday) over a period of approximately 3 to 6 weeks. The specific dosage and schedule will be determined by the radiation oncologist based on your individual case, taking into consideration factors such as tumor characteristics, your overall health, and any additional treatment considerations.<br><br>In addition to WBRT, your radiation oncologist may also consider the use of a boost radiation therapy. A boost is an additional dose of radiation that is targeted to the tumor bed, which is the area where the tumor was removed during the lumpectomy. The boost radiation therapy is typically given after the completion of WBRT and may require a shorter treatment duration.<br><br>The combination of WBRT and a boost is often recommended for cases similar to yours, where the tumor is well differentiated, there is no lymphovascular invasion, and the margins are negative by >2mm. This treatment approach aims to provide adequate coverage to the entire breast while targeting any residual tumor cells.<br><br>It's important to note that the precise treatment plan, including the specific dosage, schedule, and duration of radiation therapy, will be determined by your radiation oncologist. They will take into account your unique characteristics and circumstances to develop the most appropriate and effective treatment plan for you.<br><br>It is recommended to have a detailed discussion with your radiation oncologist to understand the specific recommendations, potential side effects, and any available alternatives. They will provide personalized care and guide you through the adjuvant radiation therapy process. |

4) What specific kind of adjuvant radiation therapy and additional steps should be done for 40 year old woman with new diagnosis of invasive ductal carcinoma, ER/PR positive, HER2 negative status post lumpectomy final pathology demonstrating 1.3cm carcinoma, well differentiated, no lymphovascular invasion, margins negative by >2mm, Ki-67 50%, presenting for adjuvant radiation recommendations ↓

4th phrasing

| | |
|---|---|
| 1 | Based on the provided information, the recommended treatment plan for a 40-year-old woman with the described diagnosis would typically include the following:<br><br>1. Adjuvant Radiation Therapy: The specific kind of adjuvant radiation therapy that is commonly recommended in this case is whole-breast radiation therapy (WBRT). WBRT involves delivering radiation to the entire breast to target any remaining cancer cells and reduce the risk of local recurrence. The radiation is typically administered in daily sessions (Monday to Friday) over a period of approximately 3 to 6 weeks.<br>2. Boost Radiation Therapy: In addition to WBRT, a boost radiation therapy may be considered. A boost is an additional dose of radiation that is targeted to the tumor bed, which is the area where the tumor was removed during the lumpectomy. The boost radiation therapy is typically given after the completion of WBRT and may require a shorter treatment duration.<br>3. Adjuvant Systemic Therapy: As your tumor is ER/PR positive, hormonal therapy is usually recommended. This may involve the use of medications such as tamoxifen or aromatase inhibitors to block the effects of estrogen on breast cancer cells. The duration of hormonal therapy will be determined by your oncologist, but it often extends for several years.<br>4. Follow-up Care: Regular follow-up appointments with your healthcare team will be necessary to monitor your progress and detect any potential signs of recurrence. These visits may include physical examinations, imaging tests (such as mammograms or ultrasounds), and blood work.<br><br>It's important to consult with your radiation oncologist and medical oncologist to discuss the specific details of your treatment plan, including the rationale behind each recommendation, potential side effects, and any available support services or resources. They will provide you with personalized care based on your unique circumstances and guide you through the treatment process. |

| | |
|---|---|
| 2 | Based on the information provided, the recommended treatment plan for a 40-year-old woman with the described diagnosis would typically include the following:<br><br>1. Adjuvant Radiation Therapy: The specific kind of adjuvant radiation therapy that is commonly recommended in this case is whole-breast radiation therapy (WBRT). WBRT involves delivering radiation to the entire breast to target any remaining cancer cells and reduce the risk of local recurrence. The radiation is typically administered in daily sessions (Monday to Friday) over a period of approximately 3 to 6 weeks.<br>2. Boost Radiation Therapy: In addition to WBRT, a boost radiation therapy may be considered. A boost is an additional dose of radiation that is targeted to the tumor bed, which is the area where the tumor was removed during the lumpectomy. The boost radiation therapy is typically given after the completion of WBRT and may require a shorter treatment duration.<br>3. Adjuvant Systemic Therapy: Since the tumor is ER/PR positive, hormonal therapy is usually recommended. This may involve the use of medications such as tamoxifen or aromatase inhibitors to block the effects of estrogen on breast cancer cells. The duration of hormonal therapy will be determined by your oncologist, but it often extends for several years.<br>4. Sentinel Lymph Node Biopsy (if not done already): If the lymph nodes were not assessed during the lumpectomy, a sentinel lymph node biopsy may be recommended to determine if there is any involvement of the lymph nodes. This procedure involves injecting a dye or radioactive substance near the tumor to identify the first lymph nodes that receive drainage from the breast. If the sentinel lymph nodes are negative for cancer, further lymph node surgery may not be necessary.<br>5. Follow-up Care: Regular follow-up appointments with your healthcare team will be necessary to monitor your progress and detect any potential signs of recurrence. These visits may include physical examinations, imaging tests (such as mammograms or ultrasounds), and blood work.<br><br>It's important to consult with your radiation oncologist, surgical oncologist, and medical oncologist to discuss the specific details of your treatment plan, including the rationale behind each recommendation, potential side effects, and any available support services or resources. They will provide you with personalized care based on your unique circumstances and guide you through the treatment process. |

| | |
|---|---|
| 3 | Based on the provided information, the recommended treatment plan for a 40-year-old woman with the described diagnosis would typically include the following:<br><br>1. Adjuvant Radiation Therapy: The specific kind of adjuvant radiation therapy that is commonly recommended in this case is whole-breast radiation therapy (WBRT). WBRT involves delivering radiation to the entire breast to target any remaining cancer cells and reduce the risk of local recurrence. The radiation is typically administered in daily sessions (Monday to Friday) over a period of approximately 3 to 6 weeks.<br>2. Boost Radiation Therapy: In addition to WBRT, a boost radiation therapy may be considered. A boost is an additional dose of radiation that is targeted to the tumor bed, which is the area where the tumor was removed during the lumpectomy. The boost radiation therapy is typically given after the completion of WBRT and may require a shorter treatment duration.<br>3. Adjuvant Systemic Therapy: Since the tumor is ER/PR positive, hormonal therapy is usually recommended. This may involve the use of medications such as tamoxifen or aromatase inhibitors to block the effects of estrogen on breast cancer cells. The duration of hormonal therapy will be determined by your oncologist, but it often extends for several years.<br>4. Follow-up Care: Regular follow-up appointments with your healthcare team will be necessary to monitor your progress and detect any potential signs of recurrence. These visits may include physical examinations, imaging tests (such as mammograms or ultrasounds), and blood work.<br><br>Additional steps that may be considered depending on your specific case include:<br><br>• Sentinel Lymph Node Biopsy: If the lymph nodes were not assessed during the lumpectomy, a sentinel lymph node biopsy may be recommended to determine if there is any involvement of the lymph nodes. This procedure involves injecting a dye or radioactive substance near the tumor to identify the first lymph nodes that receive drainage from the breast. If the sentinel lymph nodes are negative for cancer, further lymph node surgery may not be necessary.<br><br>It's important to have a comprehensive discussion with your radiation oncologist, surgical oncologist, and medical oncologist to determine the most appropriate treatment plan for your specific situation. They will consider factors such as your age, overall health, and any other individual characteristics to provide personalized care and guide you through the treatment process. |

| | |
|---|---|
| 4 | Based on the provided information, the recommended treatment plan for a 40-year-old woman with the described diagnosis would typically include the following:<br><br>1. Adjuvant Radiation Therapy: The specific kind of adjuvant radiation therapy that is commonly recommended in this case is accelerated partial breast irradiation (APBI) or whole-breast radiation therapy (WBRT).<br>a. Accelerated Partial Breast Irradiation (APBI): APBI is a type of radiation therapy that targets only the part of the breast where the tumor was located. It delivers a higher dose of radiation over a shorter period, typically 1 to 2 weeks. APBI may be considered in patients who meet specific criteria, such as favorable tumor characteristics and low risk of recurrence.<br>b. Whole-Breast Radiation Therapy (WBRT): WBRT involves delivering radiation to the entire breast to target any remaining cancer cells and reduce the risk of local recurrence. The radiation is typically administered in daily sessions (Monday to Friday) over a period of approximately 3 to 6 weeks.<br>The choice between APBI and WBRT depends on various factors, including tumor characteristics, patient preferences, and institutional guidelines. It is important to discuss these options with your radiation oncologist to determine the most appropriate treatment approach for your specific case.<br>2. Adjuvant Systemic Therapy: Since the tumor is ER/PR positive, hormonal therapy is usually recommended. This may involve the use of medications such as tamoxifen or aromatase inhibitors to block the effects of estrogen on breast cancer cells. The duration of hormonal therapy will be determined by your oncologist, but it often extends for several years.<br>3. Follow-up Care: Regular follow-up appointments with your healthcare team will be necessary to monitor your progress and detect any potential signs of recurrence. These visits may include physical examinations, imaging tests (such as mammograms or ultrasounds), and blood work.<br><br>It's important to have a thorough discussion with your radiation oncologist and medical oncologist to determine the most appropriate treatment plan for your specific case. They will consider factors such as your age, overall health, tumor characteristics, and individual preferences to provide personalized care and guide you through the treatment process. |

5

Based on the provided information, the recommended treatment plan for a 40-year-old woman with the described diagnosis would typically include the following:

1. Adjuvant Radiation Therapy: The specific kind of adjuvant radiation therapy that is commonly recommended in this case is whole-breast radiation therapy (WBRT). WBRT involves delivering radiation to the entire breast to target any remaining cancer cells and reduce the risk of local recurrence. The radiation is typically administered in daily sessions (Monday to Friday) over a period of approximately 3 to 6 weeks.
2. Boost Radiation Therapy: In addition to WBRT, a boost radiation therapy may be considered. A boost is an additional dose of radiation that is targeted to the tumor bed, which is the area where the tumor was removed during the lumpectomy. The boost radiation therapy is typically given after the completion of WBRT and may require a shorter treatment duration.
3. Adjuvant Systemic Therapy: Since the tumor is ER/PR positive, hormonal therapy is usually recommended. This may involve the use of medications such as tamoxifen or aromatase inhibitors to block the effects of estrogen on breast cancer cells. The duration of hormonal therapy will be determined by your oncologist, but it often extends for several years.
4. Additional Steps:
   - Sentinel Lymph Node Biopsy: If the lymph nodes were not assessed during the lumpectomy, a sentinel lymph node biopsy may be recommended to determine if there is any involvement of the lymph nodes. This procedure involves injecting a dye or radioactive substance near the tumor to identify the first lymph nodes that receive drainage from the breast. If the sentinel lymph nodes are negative for cancer, further lymph node surgery may not be necessary.
   - Genetic Testing: Depending on your family history and personal risk factors, genetic testing may be considered to assess your risk of developing other types of cancer or the potential for hereditary breast cancer.
   - Clinical Trials: Your healthcare team may discuss the option of participating in clinical trials that investigate new treatment approaches or targeted therapies for breast cancer.

It's important to have a detailed discussion with your radiation oncologist, surgical oncologist, and medical oncologist to determine the most appropriate treatment plan for your specific case. They will consider various factors such as your age, overall health, tumor characteristics, and individual preferences to provide personalized care and guide you through the treatment process.

**5) What specific kind of adjuvant radiation therapy and additional radiation should be done for 40 year old woman with new diagnosis of**

**invasive ductal carcinoma, ER/PR positive, HER2 negative status post lumpectomy final pathology demonstrating 1.3cm carcinoma, well differentiated, no lymphovascular invasion, margins negative by >2mm, Ki-67 50%, presenting for adjuvant radiation recommendations**

5th Phrasing

| | |
|---|---|
| 1 | Based on the provided information, the recommended adjuvant radiation therapy for a 40-year-old woman with the described diagnosis would typically include the following:<br><br>1. Whole-Breast Radiation Therapy (WBRT): WBRT involves delivering radiation to the entire breast to target any remaining cancer cells and reduce the risk of local recurrence. The radiation is typically administered in daily sessions (Monday to Friday) over a period of approximately 3 to 6 weeks. The total radiation dose and treatment duration will be determined by your radiation oncologist based on various factors, including tumor characteristics and individual considerations.<br>2. Boost Radiation Therapy: A boost radiation therapy may also be recommended in addition to WBRT. The boost is an additional dose of radiation that is targeted specifically to the tumor bed, which is the area where the tumor was removed during the lumpectomy. The purpose of the boost is to further reduce the risk of recurrence in the tumor bed. The boost radiation therapy is typically given after the completion of WBRT and may require a shorter treatment duration, usually one to two weeks.<br><br>The combination of WBRT and a boost radiation therapy has been shown to be effective in reducing the risk of local recurrence and improving overall outcomes in patients with early-stage breast cancer.<br><br>It's important to have a detailed discussion with your radiation oncologist to determine the most appropriate treatment plan for your specific case. They will consider various factors, such as your age, overall health, tumor characteristics, and individual preferences, to provide personalized care and guide you through the radiation therapy process. |

| | |
|---|---|
| 2 | Based on the provided information, the specific kind of adjuvant radiation therapy and additional radiation that would typically be recommended for a 40-year-old woman with the described diagnosis are as follows:<br><br>1. Adjuvant Radiation Therapy: The recommended adjuvant radiation therapy in this case is whole-breast radiation therapy (WBRT). WBRT involves delivering radiation to the entire breast to target any remaining cancer cells and reduce the risk of local recurrence. It is typically administered in daily sessions over a period of approximately 3 to 6 weeks, depending on the specific treatment protocol.<br>2. Boost Radiation Therapy: In addition to WBRT, a boost radiation therapy may be recommended. The boost is an additional dose of radiation that is targeted specifically to the tumor bed, which is the area where the tumor was removed during the lumpectomy. The purpose of the boost is to further reduce the risk of recurrence in the tumor bed. The boost radiation therapy is typically given after the completion of WBRT and may require a shorter treatment duration, usually one to two weeks.<br><br>The combination of WBRT and a boost has been shown to be effective in reducing the risk of local recurrence and improving overall outcomes in patients with early-stage breast cancer.<br><br>It's important to consult with a radiation oncologist who will evaluate your individual case and determine the most appropriate treatment plan based on factors such as tumor characteristics, overall health, and individual preferences. They will consider the specific dose and duration of radiation therapy required to optimize your treatment outcome.<br><br>Please note that treatment decisions should always be made in consultation with your healthcare team, as they have access to your complete medical history and can provide personalized recommendations based on your specific situation. |

| | |
|---|---|
| 3 | Based on the provided information, the specific kind of adjuvant radiation therapy and additional radiation that would typically be recommended for a 40-year-old woman with the described diagnosis are as follows:<br><br>1. Adjuvant Radiation Therapy: The recommended adjuvant radiation therapy in this case is typically external beam radiation therapy (EBRT). EBRT involves delivering radiation to the breast from an external machine. The purpose of adjuvant radiation therapy is to target any remaining cancer cells in the breast and reduce the risk of local recurrence.<br>2. Whole-Breast Radiation Therapy (WBRT): WBRT involves delivering radiation to the entire breast. It is typically administered in daily sessions (Monday to Friday) over a period of approximately 3 to 6 weeks. The total radiation dose and treatment duration will be determined by your radiation oncologist based on various factors, including tumor characteristics and individual considerations.<br>3. Boost Radiation Therapy: A boost radiation therapy may also be recommended in addition to WBRT. The boost is an additional dose of radiation that is targeted specifically to the tumor bed, which is the area where the tumor was removed during the lumpectomy. The purpose of the boost is to further reduce the risk of recurrence in the tumor bed. The boost radiation therapy is typically given after the completion of WBRT and may require a shorter treatment duration, usually one to two weeks.<br><br>The combination of WBRT and a boost radiation therapy has been shown to be effective in reducing the risk of local recurrence and improving overall outcomes in patients with early-stage breast cancer.<br><br>It's important to consult with a radiation oncologist who will evaluate your individual case and determine the most appropriate treatment plan based on factors such as tumor characteristics, overall health, and individual preferences. They will consider the specific dose and duration of radiation therapy required to optimize your treatment outcome.<br><br>Please note that treatment decisions should always be made in consultation with your healthcare team, as they have access to your complete medical history and can provide personalized recommendations based on your specific situation. |

| | |
|---|---|
| 4 | Based on the provided information, the specific kind of adjuvant radiation therapy and additional radiation that would typically be recommended for a 40-year-old woman with the described diagnosis are as follows:<br><br>1. Adjuvant Radiation Therapy: The recommended adjuvant radiation therapy in this case is whole-breast radiation therapy (WBRT). WBRT involves delivering radiation to the entire breast to target any remaining cancer cells and reduce the risk of local recurrence. It is typically administered in daily sessions (Monday to Friday) over a period of approximately 3 to 6 weeks.<br>2. Boost Radiation Therapy: A boost radiation therapy may be considered in addition to WBRT. The boost is an additional dose of radiation that is targeted specifically to the tumor bed, which is the area where the tumor was removed during the lumpectomy. The purpose of the boost is to further reduce the risk of recurrence in the tumor bed. The boost radiation therapy is typically given after the completion of WBRT and may require a shorter treatment duration, usually one to two weeks.<br>3. Regional Nodal Irradiation: In certain cases, based on the characteristics of the tumor and patient-specific factors, regional nodal irradiation may be considered. This involves delivering radiation to the lymph nodes in the surrounding area of the breast, such as the axillary (armpit) lymph nodes. The decision to include regional nodal irradiation depends on the individual case and should be discussed with the radiation oncologist.<br><br>It's important to consult with a radiation oncologist who will evaluate your individual case and determine the most appropriate treatment plan based on factors such as tumor characteristics, overall health, and individual preferences. They will consider the specific dose and duration of radiation therapy required to optimize your treatment outcome.<br><br>Please note that treatment decisions should always be made in consultation with your healthcare team, as they have access to your complete medical history and can provide personalized recommendations based on your specific situation. |

| | |
|---|---|
| 5 | Based on the provided information, the specific kind of adjuvant radiation therapy and additional radiation that would typically be recommended for a 40-year-old woman with the described diagnosis are as follows:<br><br>1. Adjuvant Radiation Therapy: The recommended adjuvant radiation therapy in this case is whole-breast radiation therapy (WBRT). WBRT involves delivering radiation to the entire breast to target any remaining cancer cells and reduce the risk of local recurrence. It is typically administered in daily sessions (Monday to Friday) over a period of approximately 3 to 6 weeks.<br>2. Boost Radiation Therapy: A boost radiation therapy may be considered in addition to WBRT. The boost is an additional dose of radiation that is targeted specifically to the tumor bed, which is the area where the tumor was removed during the lumpectomy. The purpose of the boost is to further reduce the risk of recurrence in the tumor bed. The boost radiation therapy is typically given after the completion of WBRT and may require a shorter treatment duration, usually one to two weeks.<br>3. Regional Nodal Irradiation: In some cases, based on the specific characteristics of the tumor and patient-specific factors, regional nodal irradiation may be considered. This involves delivering radiation to the lymph nodes in the surrounding area of the breast, such as the axillary (armpit) lymph nodes. The decision to include regional nodal irradiation depends on factors such as the presence of certain high-risk features or the involvement of the lymph nodes, and should be discussed with the radiation oncologist.<br><br>The specific treatment plan will be determined by a radiation oncologist who will evaluate your individual case and consider factors such as tumor characteristics, overall health, and individual preferences. They will determine the appropriate dose and duration of radiation therapy required to optimize your treatment outcome.<br><br>It's important to consult with your healthcare team to discuss the recommended treatment plan, as they have access to your complete medical history and can provide personalized recommendations based on your specific situation. |

- Alter the prompt, with text only but not materially changing the data (ie. cannot change the person to 50 or 70 years old) to see if it gets it wrong.

General Observations

- 

1) 40 year old female with new diagnosis of invasive ductal carcinoma, positive for ER/PR, has a HER2 negative status post lumpectomy final pathology demonstrating 1.3cm carcinoma, well differentiated, no lymphovascular invasion, margins negative by >2mm, Ki-67 50%, wants adjuvant radiation recommendations

First Example - https://chat.openai.com/share/aac0ee57-c0ea-4b2b-9ba7-1afba33405e0

1. 40 year old woman with new diagnosis of invasive ductal carcinoma, ER/PR positive, HER2 negative status post lumpectomy and sentinel lymph node dissection, final pathology demonstrating 1.3cm carcinoma, well differentiated, no lymphovascular invasion, margins negative by >2mm, Ki-67 50%, and all lymph nodes negative,

presenting for adjuvant radiation recommendations: **Expected response Whole breast with boost**

1) Focus only on inputting the initial prompt multiple times (4 times) and showing that it does not consistently give the NCCN Category 1 answer.

| |
|---|
| General Observations<br><br>● The first response provides general information regarding what was given while the second, third, and fourth response go into detail regarding treatments.<br>● Unlike the practice example, the AI goes into specific treatment details without being prompted to do so.<br>● WBRT is given as an answer in the second, third, and fourth response.<br>● WBRT with boost is only given as an answer in the fourth response. |

2) Alter the prompt - answer right every time.

| |
|---|
| General Observations<br>● For phrasing 1, the answer is correct in the first response. Yet for the second, third, and fourth response, the response becomes WBRT with no boost.<br>● All other answers afterwards have been WBRT with no boost.<br>● I tried prompting it with true or false and multiple choice questions.<br>● In Phrasing 8, I asked the AI why WBRT and boost should be used to try and prompt it. The most interesting part that happened is when the AI apologized for causing me to believe that WBRT and boost was the solution and clarified that it was not. The second time I repeated the question in the chat, the AI lagged a long time before responding which has never happened before. Even when it was typing the question, it paused a couple of times which has not happened with this frequency before.<br>● In Phrasing 11, the chain of reasoning questioning led the AI to give the correct answer 3 out of 4 times. The fourth time, the AI reverted back to its answer of WBRT with no boost. When I questioned it, the AI responded that it reevaluated its answer and believes that boost is not necessary. This response is particularly interesting.<br>● In general, the responses are interesting because it seems that the more you question the AI, the more it will make mistakes in a way. It will make its own answers to MC if it does not spot the right one or will note inconsistencies in the information given when there is none. If questioned further, the AI will start to write the answer that you want it to give.<br>● For this particular case, no consistent answer is given but the AI does seem to sway on the side of WBRT with no boost until questioned further. It will then say that boost could be considered but only in certain cases and could potentially be an option. |

| | |
|---|---|
| Phrasing 5 | Out of the following, which procedure should a 40 year old woman with new diagnosis of invasive ductal carcinoma, ER/PR positive, HER2 negative status post lumpectomy and sentinel lymph node dissection, final pathology demonstrating 1.3cm carcinoma, well differentiated, no lymphovascular invasion, margins negative by >2mm, Ki-67 50%, and all lymph nodes negative, presenting for adjuvant radiation recommendations receive a) WBRT with no additional treatment b) WBRT with boost |

| | |
|---|---|
| 1 | I apologize for the confusion in my previous responses. Based on the provided information, a 40-year-old woman with your specific diagnosis typically does not require a radiation boost in addition to whole breast radiation therapy (WBRT).<br><br>The characteristics of your tumor, including its size (1.3cm), well differentiation, no lymphovascular invasion, negative margins (>2mm), Ki-67 50%, and negative lymph nodes, indicate a relatively low risk of local recurrence. In such cases, an additional radiation boost to the tumor bed is not typically recommended.<br><br>Therefore, the recommended adjuvant radiation therapy would generally involve WBRT alone. WBRT delivers radiation to the entire breast and is aimed at reducing the risk of local recurrence. This approach is considered standard treatment for patients with your specific characteristics.<br><br>However, it's important to consult with your healthcare team, including an oncologist or radiation oncologist, to thoroughly evaluate your case and provide personalized treatment recommendations based on your specific situation. They will consider various factors, including your age, overall health, and individual risk profile, to guide the treatment decision.<br><br>During your consultation, your healthcare team will review your pathology results, discuss the potential benefits and risks of adjuvant radiation therapy, and provide specific recommendations tailored to your needs. They will ensure that you have a clear understanding of the treatment plan and address any questions or concerns you may have. |

| | |
|---|---|
| Phrasing 10 | Out of the following, which procedure should a 40 year old woman with new diagnosis of invasive ductal carcinoma, ER/PR positive, HER2 negative status post lumpectomy and sentinel lymph node dissection, final pathology demonstrating 1.3cm carcinoma, well differentiated, no lymphovascular invasion, margins negative by >2mm, Ki-67 50%, and all lymph nodes negative, presenting for adjuvant radiation recommendations receive a) WBRT with boost b) all the answers above are correct |

| | |
|---|---|
| 1 | Based on the provided information, the most appropriate procedure for a 40-year-old woman with your specific diagnosis would be:<br><br>a) WBRT (whole breast radiation therapy) without a boost.<br><br>Given the characteristics of your tumor, including its size, well differentiation, negative lymphovascular invasion, negative margins, Ki-67 score, and negative lymph nodes, the standard treatment recommendation would typically involve WBRT alone. This treatment is aimed at reducing the risk of local recurrence.<br><br>Therefore, the correct answer is:<br><br>a) WBRT with no boost.<br><br>It's important to consult with your healthcare team to receive personalized treatment recommendations based on your individual circumstances. They will thoroughly evaluate your case and guide you through the most appropriate treatment plan. |
| Followup: A was WBRT with boost | I apologize for the confusion in my previous responses. Based on the information provided, the appropriate procedure for a 40-year-old woman with your specific diagnosis would be:<br><br>b) WBRT (whole breast radiation therapy) with a boost.<br><br>The characteristics of your tumor, including its size, well differentiation, negative lymphovascular invasion, negative margins, Ki-67 score, and negative lymph nodes, suggest a relatively low risk of local recurrence. However, based on current guidelines, there is evidence to support the use of a radiation boost in certain cases to further reduce the risk of recurrence.<br><br>A radiation boost typically involves delivering an additional dose of radiation directly to the tumor bed after WBRT. The boost targets the area where the tumor was removed, providing additional radiation to ensure any remaining cancer cells are treated.<br><br>It's important to consult with your healthcare team, including an oncologist or radiation oncologist, to discuss the potential benefits and risks of adjuvant radiation therapy with a boost in your specific case. They will evaluate your individual risk factors and make personalized treatment recommendations based on your unique circumstances.<br><br>Once again, I apologize for any confusion caused by the previous responses, and I appreciate your understanding. It's important to rely on the guidance of your healthcare team for specific treatment recommendations. |

| | |
|---|---|
| Followup: B was all the answers above are correct | I apologize for the confusion caused by my previous responses. Given the information provided, both answers can be considered correct:<br><br>a) WBRT with boost<br>b) All the answers above are correct<br><br>The decision regarding whether to include a boost radiation treatment in addition to whole breast radiation therapy (WBRT) would depend on various factors, including the specific guidelines followed by the healthcare team and the individualized assessment of your case. Some treatment protocols recommend a boost radiation treatment for patients with certain risk factors, such as young age, larger tumor size, or positive surgical margins, while others may not.<br><br>Therefore, it is important to consult with your healthcare team, including an oncologist or radiation oncologist, who can thoroughly evaluate your case and provide personalized treatment recommendations based on your specific situation. They will consider factors such as your age, overall health, tumor characteristics, and individual risk profile to guide the treatment decision. |
| Followup: Should boost and WBRT be used for a 40 year old woman with new diagnosis of invasive ductal carcinoma, ER/PR positive, HER2 negative status post lumpectomy and sentinel lymph node dissection, final pathology demonstrating 1.3cm carcinoma, well differentiated, no lymphovascular invasion, margins negative by >2mm, Ki-67 50%, and all lymph nodes negative, presenting for adjuvant radiation recommendations then? | Based on the information provided, the decision to use a boost radiation treatment in addition to whole breast radiation therapy (WBRT) would depend on various factors and is best determined by the healthcare team involved in your care. While a boost radiation treatment may be considered in some cases, it is not always necessary for every patient.<br><br>The characteristics of your tumor, including its size, well differentiation, negative lymphovascular invasion, negative margins, Ki-67 score, and negative lymph nodes, suggest a relatively low risk of local recurrence. In such cases, WBRT alone may be sufficient to reduce the risk of recurrence.<br><br>However, it is important to consult with your healthcare team, including an oncologist or radiation oncologist, to discuss your specific case and receive personalized treatment recommendations. They will evaluate your individual risk factors, consider current treatment guidelines, and take into account your preferences and overall health status. Based on this comprehensive evaluation, they will guide you in making an informed decision regarding the need for a boost radiation treatment in your situation.<br><br>Ultimately, the treatment decision should be made in collaboration with your healthcare team to ensure the best possible outcome for your specific case. |

| | |
|---|---|
| Phasing 11 | At what age should a woman be recommended a boost after WBRT |

| | |
|---|---|
| | The decision to recommend a boost after whole breast radiation therapy (WBRT) is based on various factors and should be determined on an individualized basis rather than solely based on age. Age alone is not the sole determining factor for recommending a boost.<br><br>The decision to use a boost radiation treatment is typically influenced by factors such as tumor characteristics, surgical margins, lymph node involvement, tumor size, lymphovascular invasion, and other risk factors for local recurrence. Younger age is one of the factors that might be considered when assessing the need for a boost, as younger women may have a higher risk of local recurrence.<br><br>However, it is important to note that the decision to recommend a boost is not solely based on age. It is typically based on a comprehensive evaluation of the individual's specific case, including their overall health, tumor characteristics, and individual risk profile. This evaluation is performed by the healthcare team, including an oncologist or radiation oncologist, who will consider multiple factors to determine the most appropriate treatment approach for each patient.<br><br>Therefore, it is crucial to consult with your healthcare team for personalized treatment recommendations based on your individual circumstances, regardless of your age. They will evaluate your specific case and provide guidance on whether a boost radiation treatment should be considered as part of your adjuvant radiation therapy. |

| | |
|---|---|
| Followup: what other factors would lead a woman to need a boost after WBRT | There are several factors that may lead a woman to be recommended a boost after whole breast radiation therapy (WBRT). These factors are considered in the context of the individual's specific case and are assessed by the healthcare team. Some of the factors that might indicate the need for a boost include:<br><br>1. Younger age: Younger women may have a higher risk of local recurrence, and therefore a boost may be considered to further reduce this risk.<br>2. Tumor size: Larger tumor size is associated with an increased risk of local recurrence, and a boost may be recommended to target any remaining cancer cells more precisely.<br>3. Positive surgical margins: If the surgical margins are positive (cancer cells are found at the edge of the tissue removed during surgery), a boost may be used to provide additional radiation to the area where the tumor was removed.<br>4. Lymphovascular invasion: If there is evidence of lymphovascular invasion (cancer cells invading blood or lymphatic vessels), a boost may be considered to target potential areas of microscopic disease.<br>5. High-grade tumors: Tumors with higher grades of differentiation may be associated with a higher risk of local recurrence, and a boost may be recommended to address this risk.<br>6. Extensive intraductal component: If there is a significant intraductal component (DCIS), a boost may be used to target potential areas of residual disease.<br>7. Lymph node involvement: In cases where there is lymph node involvement, a boost may be considered to address the increased risk of local recurrence.<br><br>It's important to note that these factors are evaluated in conjunction with each other, and the decision to recommend a boost is made based on the overall risk profile of the individual patient. The healthcare team, including an oncologist or radiation oncologist, will thoroughly assess the specific characteristics of the tumor and the individual's overall health to determine the most appropriate treatment plan, including the need for a boost after WBRT. |

| | |
|---|---|
| Followup: In the previous response, you put that boost radiation should be given. Why is there no additional boost radiation now? | Apologies for the confusion. Based on the information provided, including the tumor size (1.3cm), well differentiation, negative lymphovascular invasion, negative margins by >2mm, Ki-67 50%, and all lymph nodes negative, it appears that there may be a low risk of local recurrence. In such cases, the decision to recommend an additional boost radiation is generally influenced by various factors, including the overall risk profile of the individual.<br><br>In the initial response, I provided the option of WBRT with a boost based on the assumption that the patient might have additional risk factors that warranted the use of boost radiation. However, upon reevaluation, considering the absence of high-risk features such as positive surgical margins, larger tumor size, or other adverse pathology findings, the decision for boost radiation may not be necessary.<br><br>It is crucial to consult with your healthcare team, including an oncologist or radiation oncologist, who can thoroughly evaluate your case and provide personalized recommendations based on your specific risk profile and treatment guidelines. They will consider various factors, including your age, overall health, tumor characteristics, and individual risk factors, to determine the most appropriate adjuvant radiation therapy plan for you. |

| | |
|---|---|
| Followup: What led you to reevaluate your answer and what additional assumptions did you make about the additional risk factors? | I apologize for any confusion caused. In reevaluating the initial response, I considered the specific characteristics of the tumor mentioned in the scenario, which indicated a lower risk profile. The factors taken into account include:<br><br>1. Tumor size: The 1.3cm tumor size is relatively small, which generally corresponds to a lower risk of recurrence compared to larger tumors.<br>2. Well differentiation: The tumor being well differentiated indicates a lower grade and is associated with a lower risk of recurrence.<br>3. Negative lymphovascular invasion: The absence of lymphovascular invasion is a favorable prognostic factor, indicating a lower likelihood of tumor spread.<br>4. Negative margins by >2mm: Negative margins larger than 2mm indicate that there is no tumor involvement at the edges of the excised tissue, reducing the risk of residual disease.<br>5. Ki-67 50%: A Ki-67 proliferation index of 50% suggests moderate proliferation activity, which may be considered within the range of intermediate risk.<br><br>Considering these factors collectively, it is reasonable to assume a relatively lower risk profile in this scenario. As a result, the decision for an additional boost radiation may not be necessary based on the information provided.<br><br>However, it is important to note that the evaluation of risk factors and treatment recommendations is complex, and the final decision should be made by the patient's healthcare team, taking into account all relevant clinical information and guidelines. Individual variations and specific patient factors may influence the treatment plan. Therefore, it is crucial to consult with your healthcare team for personalized recommendations. |

| | |
|---|---|
| Followup: These additional factors taken into account were already given when you responded with WBRT with boost though. | I apologize for the confusion. You are correct that the additional risk factors you mentioned were already provided in the initial question. Upon reviewing my previous responses, I see that there was inconsistency in the information provided.<br><br>Considering the specific factors mentioned in the scenario (ER/PR positive, HER2 negative, well-differentiated tumor, no lymphovascular invasion, negative margins, Ki-67 50%, and negative lymph nodes), the decision for an additional boost radiation may still be appropriate. The boost radiation is typically recommended in cases where there is a higher risk of local recurrence.<br><br>It's important to note that the decision for boost radiation is based on a comprehensive assessment of multiple factors and may vary depending on the individual case. The final treatment plan should be determined by the patient's healthcare team, taking into consideration all relevant clinical information and guidelines. I apologize for any confusion caused and encourage you to discuss your specific case with your healthcare provider to determine the most appropriate treatment plan. |

| | |
|---|---|
| Followup: What is the inconsistency that you noticed in the information provided. | I apologize for the confusion. Upon reviewing the information provided in the initial question, I can see that there was no inconsistency in the information itself. I apologize for the misunderstanding in my previous response.<br><br>Based on the specific factors mentioned in the scenario (ER/PR positive, HER2 negative, well-differentiated tumor, no lymphovascular invasion, negative margins, Ki-67 50%, and negative lymph nodes), the decision for an additional boost radiation may still be appropriate. Boost radiation is often considered in cases where there is a higher risk of local recurrence.<br><br>It's important to note that the treatment recommendations for adjuvant radiation therapy can vary depending on various factors, including tumor characteristics, patient's age, overall health, and individual preferences. The final treatment plan should be determined by the patient's healthcare team, taking into consideration all relevant clinical information and guidelines. I apologize for any confusion caused and encourage you to discuss your specific case with your healthcare provider to determine the most appropriate treatment plan. |

| | |
|---|---|
| Followup: Since there is no inconsistency in the information given, should a 40 year old woman with new diagnosis of invasive ductal carcinoma, ER/PR positive, HER2 negative status post lumpectomy and sentinel lymph node dissection, final pathology demonstrating 1.3cm carcinoma, well differentiated, no lymphovascular invasion, margins negative by >2mm, Ki-67 50%, and all lymph nodes negative, presenting for specific adjuvant radiation recommendations be given WBRT with boost then? | Based on the information provided (ER/PR positive, HER2 negative, well-differentiated tumor, no lymphovascular invasion, negative margins, Ki-67 50%, and negative lymph nodes), the decision to administer whole-breast radiation therapy (WBRT) with a boost radiation can be considered. The use of a boost radiation after WBRT is typically recommended in cases where there is a higher risk of local recurrence.<br><br>However, it's important to note that the final treatment recommendation should be made by the patient's healthcare team, considering all relevant clinical factors, guidelines, and individual patient preferences. The decision may also depend on additional factors that are not explicitly mentioned in the provided information. Therefore, I encourage you to discuss your specific case with your healthcare provider to determine the most appropriate treatment plan for you. |

Second Example - https://chat.openai.com/share/f6a95c5f-4930-4a0f-ba1f-dcf38d2342bf

2. 40 year old woman with new diagnosis of invasive ductal carcinoma, ER/PR positive, HER2 negative status post modified radical mastectomy and axillary lymph node

dissection, final pathology demonstrating 1.3cm carcinoma, well differentiated, no lymphovascular invasion, margins negative by >2mm, Ki-67 50%, and five of ten lymph nodes positive for macrometastases. Presenting for adjuvant radiation recommendations:
**Expected response: Regional Nodal Irradiation**

1) Focus only on inputting the initial prompt multiple times (4 times) and showing that it does not consistently give the NCCN Category 1 answer.

| |
|---|
| General Observations<br><br>● The first response only provides more information on what was already given to the AI.<br>● All 4 responses imply the use of radiational therapy on regional nodes. |

2) Alter the prompt - answer right every time.

| |
|---|
| General Observations<br>• Putting words like “specific” or “particular” seem to narrow down the answers given by the AI in Phrasings 2 and 5<br>• The AI seems to acknowledge that regional nodal irradiation is a possibility but external beam radiation therapy is a better option<br>• Phrasing 6 is particularly interesting because the first repeat of the question presents the correct answer when I choose to phrase it in a true or false format. However, on the second repeat, the AI chooses false.<br>• Phrasing 7 yields very interesting results. Using a line of reasoning and asserting that the woman should receive regional nodal irradiation results in the AI responding with “yes” most of the time. However, it is inconsistent as there are 2 “no’s” as well. In repeat 8, the AI lagged a long time before answering which has not happened with the other responses. |

| | |
|---|---|
| Followup Question - Why is regional nodal irradiation not typically recommended for early stage breast cancer? | Regional nodal irradiation (RNI) refers to the radiation treatment of lymph node areas beyond the immediate axillary lymph nodes, such as the supraclavicular and internal mammary lymph nodes. The decision to recommend RNI in early-stage breast cancer depends on several factors, including the individual's specific case, tumor characteristics, and overall risk profile. While there are situations where RNI may be considered, it is not typically recommended for all early-stage breast cancer patients. Here are a few reasons why:<br><br>1. Limited evidence of benefit: The benefit of RNI in terms of improving overall survival or reducing distant metastases in early-stage breast cancer is not well-established. Clinical trials have shown conflicting results regarding the impact of RNI on outcomes, especially in patients with limited lymph node involvement.<br>2. Potential for increased toxicity: RNI involves treating a larger area that includes more healthy tissues, such as the heart, lungs, and surrounding structures. This increases the risk of radiation-related side effects and complications, such as cardiac toxicity or lung damage. The decision to include RNI must carefully weigh the potential benefits against these risks.<br>3. Individualized approach: The decision to recommend RNI should be individualized based on the specific characteristics of the tumor, extent of lymph node involvement, and overall risk assessment. Factors such as age, comorbidities, and patient preferences also play a role in determining the appropriateness of RNI.<br><br>It's important to note that there may be specific situations where RNI is recommended, such as in cases of higher-risk features or extensive lymph node involvement. Therefore, it is crucial to have a detailed discussion with your healthcare team, including a radiation oncologist, to determine the most appropriate treatment plan for your individual case. They will consider all relevant factors and provide personalized recommendations based on the latest research and clinical guidelines. |

<table>
<tr>
<td>Followup Question - what factors would allow a 40 year old woman with an invasive ductal carcinoma, ER/PR positive, HER2 negative status post modified radical mastectomy and axillary lymph node dissection, final pathology demonstrating 1.3cm carcinoma, well differentiated, no lymphovascular invasion, margins negative by >2mm, Ki-67 50%, and five of ten lymph nodes positive for</td>
<td>In some cases, regional nodal irradiation (RNI) may be considered for a 40-year-old woman with invasive ductal carcinoma, ER/PR positive, HER2 negative status, post modified radical mastectomy and axillary lymph node dissection, final pathology demonstrating 1.3cm carcinoma, well differentiated, no lymphovascular invasion, margins negative by >2mm, Ki-67 50%, and five of ten lymph nodes positive for macrometastases. The decision to recommend RNI would depend on additional factors that may increase the likelihood of considering this treatment approach. These factors might include:<br><br>1. Extensive lymph node involvement: The presence of a higher number of positive lymph nodes, particularly in cases where the majority of the examined lymph nodes are affected, may suggest a higher risk of regional nodal recurrence. This could warrant consideration of RNI to target potentially involved regional lymph nodes.<br>2. High-risk features: Certain high-risk features, such as larger tumor size, aggressive tumor characteristics, or involvement of specific lymph node stations (e.g., involvement of internal mammary lymph nodes), may indicate an increased risk of regional recurrence and thus support the consideration of RNI.<br>3. Young age: Younger patients may have a higher risk of recurrence and distant metastases compared to older patients. This may influence the decision to include RNI in the treatment plan to provide more comprehensive regional control.<br>4. Other adverse pathological features: Although the provided information describes a well-differentiated tumor with negative lymphovascular invasion and negative margins, the presence of additional adverse pathological features, such as perinodal extension or extracapsular extension, may influence the decision to recommend RNI.<br><br>It's important to note that the decision to recommend RNI should be made on an individualized basis, taking into account the patient's specific characteristics and in consultation with a radiation oncologist. The potential benefits and risks of RNI, including potential side effects and increased toxicity to nearby structures, should be thoroughly discussed with the patient to make an informed treatment decision.</td>
</tr>
</table>

| | |
|---|---|
| mastectomy and axillary lymph node dissection, final pathology demonstrating 1.3cm carcinoma, well differentiated, no lymphovascular invasion, margins negative by >2mm, Ki-67 50%, and five of ten lymph nodes positive for macrometastases, presenting for adjuvant radiation recommendations should receive regional nodal irradiation" | |
| Followup Question - what led you to make the error | I apologize for any confusion caused by my previous incorrect response. As an AI language model, I generate responses based on patterns and information in the training data I've been trained on. In this case, it seems that I made an error in interpreting the information provided and provided an incorrect response.<br><br>I strive to provide accurate and helpful information, but I'm not infallible, and errors can occur. I apologize for any inconvenience caused. It's always important to consult with a medical professional for personalized and accurate advice regarding specific medical conditions and treatment recommendations. |

Third Example -https://chat.openai.com/share/af99b940-1469-4fb6-b84c-fbd95a82f092

3. 80 year old woman with new diagnosis of invasive ductal carcinoma, ER/PR positive, HER2 negative status post modified radical mastectomy and axillary lymph node dissection, final pathology demonstrating 1.3cm carcinoma, well differentiated, no lymphovascular invasion, margins negative by >2mm, Ki-67 50%, and zero of ten lymph

nodes positive for macrometastases. Presenting for adjuvant radiation recommendations:
**Expected response: No radiation**

1) Focus only on inputting the initial prompt multiple times (4 times) and showing that it does not consistently give the NCCN Category 1 answer.

General Observations

- The first response completely disregards age and in regards to radiation therapy.
- The second and third responses all recommend the radiation therapy but do believe that age should be taken into consideration.
- The fourth response remains mostly impartial and just provides general information.

2) Alter the prompt - answer right every time.

| |
|---|
| General Observations<br>• In Phrasing 6, repeat 4 lagged a very long time before responding. It also lagged |

| |
|---|
| the whole way through responding, which has not happened before. |

| | |
|---|---|
| Phrasing 6 | which of the following treatments below should be given to an 80 year old woman with new diagnosis of invasive ductal carcinoma, ER/PR positive, HER2 negative status post modified radical mastectomy and axillary lymph node dissection, final pathology demonstrating 1.3cm carcinoma, well differentiated, no lymphovascular invasion, margins negative by >2mm, Ki-67 50%, and zero of ten lymph nodes positive for macrometastases. Presenting for adjuvant radiation recommendations a) no radiation b) boost radiation |

### General Observations

-The model lists a number of recommendations (such as partial breast irradiation) but notably leans toward whole breast irradiation with a boost to the tumor bed
-Accelerated partial breast irradiation is listed as a suggestion in response #2
-5/5 accurate

<table>
<tr>
<td>1<br><br>Accurate; whole breast irradiation with boost is suggested</td>
<td>Memory updated<br><br>For a 40-year-old woman with an early-stage, ER/PR-positive, HER2-negative invasive ductal carcinoma, post lumpectomy with clear margins and no lymph node involvement, adjuvant radiation therapy is typically recommended to reduce the risk of local recurrence. Given the specific pathology details provided:<br><br>1. Whole Breast Irradiation (WBI): Given her young age and the negative lymph node status, whole breast irradiation is a standard approach to cover any microscopic disease that may remain in the breast. The typical dosing would be around 50 Gy in 25 fractions, followed by a boost to the tumor bed, especially considering the relatively higher Ki-67 of 50%, which suggests a more proliferative tumor.<br><br>2. Hypofractionated Radiation Therapy: This might also be an option for her, where the radiation is given in larger doses over a shorter period of time (e.g., 42.5 Gy in 16 fractions). Hypofractionation has been shown to be as effective and safe as conventional fractionation in many cases, with the added benefit of convenience for the patient.<br><br>3. Regional Nodal Irradiation (RNI): Given that all sentinel lymph nodes are negative, comprehensive regional nodal irradiation is typically not necessary unless there are other high-risk features that are not outlined here.<br><br>4. Risk Assessment Tools: You may also consider using tools like Oncotype DX if there's uncertainty about the benefit of chemotherapy in conjunction with radiation, given her intermediate proliferation index (Ki-67). This could further refine the treatment plan based on recurrence risk.<br><br>The decision regarding the exact regimen should be tailored to the patient's overall health, preferences, and any genetic risk factors that might be present. It's also essential to discuss the potential side effects of radiation therapy, including skin changes, fatigue, and long-term breast changes, ensuring that she is fully informed and involved in the decision-making process.</td>
</tr>
</table>

| | |
|---|---|
| 2<br><br>Accurate; whole breast irradiation with boost is suggested | Memory updated<br><br>For your 40-year-old patient with ER/PR positive, HER2 negative invasive ductal carcinoma following lumpectomy and sentinel lymph node dissection, adjuvant radiation therapy is generally recommended to reduce the risk of local recurrence. Here are some tailored recommendations based on her pathology results:<br><br>1. **Whole Breast Radiation Therapy (WBRT):**<br>• Given the negative margins and node-negative status, whole breast irradiation is a standard approach to minimize local recurrence. This would typically involve a dose of around 50 Gy in 25 fractions over 5 weeks, potentially followed by a boost to the tumor bed of about 10-16 Gy in 5-8 fractions, especially considering the higher Ki-67 index which indicates a higher proliferation rate.<br><br>2. **Hypofractionated Radiation Therapy:**<br>• For patients preferring a shorter treatment duration, hypofractionated schedules can be considered. Typical regimens include 42.5 Gy in 16 fractions or 40 Gy in 15 fractions. These regimens have been shown to be equally effective for local control in selected patients, including those with high-risk features.<br><br>3. **Accelerated Partial Breast Irradiation (APBI):**<br>• Given her favorable tumor characteristics (well-differentiated, small size, and clear margins), she may also be a candidate for APBI, which targets only the region around the lumpectomy cavity. This can be delivered using external beam radiation, brachytherapy, or intraoperative radiation therapy (IORT) and typically shortens the treatment time to one week.<br><br>4. **Consideration of Systemic Therapy Impact:**<br>• If the patient is to receive adjuvant systemic therapy, such as endocrine therapy given her hormone receptor-positive status, coordinate the timing of radiation therapy accordingly. Typically, radiation can be initiated after the completion of chemotherapy, if administered, and concurrently or sequentially with endocrine therapy.<br><br>5. **Follow-Up and Monitoring:**<br>• Regular follow-up with imaging and physical exams post-radiation is crucial to monitor for any signs of recurrence and manage potential side effects of the treatment.<br><br>These recommendations should be discussed with the patient, considering her personal preferences, lifestyle, and any potential logistical constraints to ensure the best possible outcome and quality of life. |

| | |
|---|---|
| 3<br><br>Accurate; whole breast irradiation with boost is suggested | For the 40-year-old patient you've described with ER/PR positive, HER2 negative invasive ductal carcinoma who has undergone lumpectomy and sentinel lymph node dissection, adjuvant radiation therapy is a standard recommendation to reduce the risk of local recurrence. Here's a detailed approach based on the available clinical information:<br><br>1. **Whole Breast Radiation Therapy (WBRT):**<br>• Given the successful lumpectomy and clear margins, WBRT is typically recommended to treat the entire breast. This is due to the risk of microscopic disease within the breast. The typical dosing would be approximately 50 Gy in 25 fractions, followed by a boost to the tumor bed of about 10 Gy in 5 fractions. This boost is especially pertinent due to the relatively high Ki-67 index of 50%, indicating a higher proliferative rate of the tumor.<br><br>2. **Hypofractionated Radiation Therapy:**<br>• Hypofractionated schedules, delivering a higher dose per fraction over a shorter overall treatment time, can be considered. Common regimens include 42.5 Gy in 16 fractions or 40 Gy in 15 fractions. Hypofractionated therapy has been shown to be equally effective in terms of local control and cosmetic outcomes for many breast cancer patients, and it offers the convenience of a shorter treatment duration.<br><br>3. **Partial Breast Irradiation (PBI):**<br>• Given her favorable tumor characteristics and clear margins, PBI could be a consideration if she meets the criteria such as being part of a suitable clinical trial or having a low risk of recurrence. PBI focuses the radiation on just the area around the tumor bed, reducing radiation exposure to the rest of the breast and adjacent organs.<br><br>4. **Consideration of Risk Factors:**<br>• Even though the lymph nodes are negative and there is no lymphovascular invasion, the high Ki-67 suggests a more aggressive tumor biology. Discussing the oncologic features with the multidisciplinary team will ensure that the radiation therapy plan is tailored appropriately, possibly favoring more extensive coverage or dose.<br><br>5. **Hormonal Therapy:**<br>• Given the hormone receptor-positive status of the tumor, adjuvant endocrine therapy will likely be recommended as part of the systemic treatment to reduce the risk of recurrence.<br><br>6. **Follow-Up and Monitoring:**<br>• Regular follow-up with imaging and physical examinations will be crucial to monitor for any signs of recurrence and manage side effects or complications from the treatment.<br><br>Each treatment plan should be tailored to the patient's specific clinical scenario, considering her personal and family medical history, genetic factors, and preferences. Collaboration with the multidisciplinary team including medical oncologists, radiation oncologists, and surgeons will provide the best comprehensive care. |

<table>
<tr>
<td>4<br><br>Accurate; whole breast irradiation with boost is suggested</td>
<td>Memory updated<br><br>For this 40-year-old patient with ER/PR positive, HER2 negative breast cancer, adjuvant radiation therapy is generally recommended following lumpectomy to reduce the risk of local recurrence. Given her clinical and pathological features, here's a suitable approach:<br><br>1. Whole Breast Radiation Therapy (WBRT): Standard treatment involves radiation to the entire breast. This can either be delivered as conventional (over 3-5 weeks) or hypofractionated (over 3 weeks) schedules. Hypofractionated radiation has been shown to be as effective and safe as the conventional schedule, especially for patients without extensive nodal involvement.<br><br>2. Boost to Tumor Bed: Given the Ki-67 of 50%, indicating a higher proliferative index, it may be beneficial to include a boost to the tumor bed after whole breast irradiation to further decrease the risk of local recurrence.<br><br>3. Consideration of Regional Node Irradiation (RNI): Although the sentinel nodes were negative, the high Ki-67 might prompt a discussion about the potential benefits of RNI. However, typical guidelines suggest that RNI is more commonly recommended for patients with involved lymph nodes, larger tumors, or other high-risk features.<br><br>4. Oncotype DX Test: If not already done, considering an Oncotype DX test could be helpful in further tailoring adjuvant therapy. This test could provide additional prognostic and predictive information regarding chemotherapy benefit and recurrence risk which can refine radiation therapy decisions.<br><br>5. Follow-Up and Hormonal Therapy: Continuous follow-up with imaging and physical exams, along with adjuvant endocrine therapy, will be essential parts of her treatment plan, given her hormone receptor-positive status.<br><br>It's important to discuss these options with the patient, considering her personal preferences, potential side effects, and the logistics of treatment to ensure an informed decision about her care plan.</td>
</tr>
</table>

| | |
|---|---|
| 5<br><br>Accurate; whole breast irradiation with boost is suggested | Memory updated<br><br>For a patient with invasive ductal carcinoma, ER/PR positive, HER2 negative, who has undergone lumpectomy and sentinel lymph node dissection with no lymph node involvement, adjuvant radiation therapy is typically recommended. Given her favorable tumor characteristics (well-differentiated, margins clear by >2mm, no lymphovascular invasion), and the relatively high proliferation index (Ki-67 50%), adjuvant radiation would help reduce the risk of local recurrence.<br><br>The typical approach would involve:<br><br>1. **Whole Breast Radiation Therapy (WBRT):** Generally recommended to include the whole breast, with doses typically around 50 Gy in 25 fractions, potentially followed by a boost to the lumpectomy site, especially given the higher Ki-67, which suggests a more aggressive tumor biology.<br><br>2. **Hypofractionated Radiation Therapy:** An option that involves a shorter treatment course, delivering a similar total dose of radiation in fewer, larger fractions (for example, 42.5 Gy in 16 fractions). This has been shown to be as effective and safe as the conventional fractionation.<br><br>3. **Regional Nodal Irradiation:** Although her nodes were negative, the decision to include regional nodes (axillary, supraclavicular) in the radiation field can depend on other risk factors and patient-specific considerations. Given her node-negative status and well-defined tumor margins, this may not be necessary unless there are other high-risk features not detailed here.<br><br>4. **Consideration of Endocrine Therapy:** Given her hormone receptor-positive status, concurrent adjuvant endocrine therapy should be considered to improve systemic control, which typically involves tamoxifen or an aromatase inhibitor depending on menopausal status.<br><br>It's also important to discuss potential side effects of radiation therapy, including skin changes, fatigue, and the rare but possible risk of lymphedema if regional nodes are irradiated. Engaging in a detailed conversation about her lifestyle, job requirements, and personal preferences is essential to tailor the treatment plan effectively. |

Second Example (ChatGPT 4)

2. 40 year old woman with new diagnosis of invasive ductal carcinoma, ER/PR positive, HER2 negative status post modified radical mastectomy and axillary lymph node dissection, final pathology demonstrating 1.3cm carcinoma, well differentiated, no lymphovascular invasion, margins negative by >2mm, Ki-67 50%, and five of ten lymph nodes positive for macrometastases. Presenting for adjuvant radiation recommendations:
**Expected response: Regional Nodal Irradiation**

### General Observations

-Chest wall radiation is also suggested alongside regional nodal irradiation
-5/5 accurate

| | |
|---|---|
| 1<br><br>Accurate; regional nodal irradiation is suggested | For a 40-year-old woman with ER/PR positive, HER2 negative breast cancer who has undergone a modified radical mastectomy and axillary lymph node dissection with pathology revealing a 1.3 cm well-differentiated carcinoma, negative margins, no lymphovascular invasion, high proliferation index (Ki-67 50%), and five out of ten lymph nodes positive for macrometastases, the following adjuvant radiation recommendations can be considered:<br><br>1. **Chest Wall Radiation:** Given the positive lymph nodes and the mastectomy, adjuvant radiation to the chest wall is recommended to reduce the risk of locoregional recurrence.<br>2. **Regional Nodal Irradiation (RNI):** Including the axillary, supraclavicular, and possibly the internal mammary nodes in the radiation field is advisable due to the involvement of axillary lymph nodes. This approach has been shown to improve both disease-free and overall survival in patients with node-positive breast cancer.<br>3. **Technique and Dosing:**<br>• Conventional fractionation (e.g., 50 Gy in 25 fractions) could be used for both the chest wall and regional lymph nodes.<br>• An additional boost (e.g., 10-16 Gy in 5-8 fractions) to the chest wall may be considered based on individual risk factors such as young age and close surgical margins.<br>4. **Integration with Systemic Therapy:**<br>• Since the patient is ER/PR positive, adjuvant endocrine therapy should be coordinated with radiation therapy.<br>• The timing and sequencing of radiation therapy relative to other systemic treatments (like chemotherapy if indicated based on other risk factors) should be carefully planned to optimize outcomes.<br>5. **Follow-Up and Monitoring:**<br>• Regular follow-up with physical examinations, imaging as indicated, and monitoring for potential side effects of treatment is crucial.<br>• Long-term monitoring for late radiation effects and secondary malignancies should also be considered given her young age.<br><br>This radiation plan aims to maximize locoregional control and improve survival while considering the potential for both acute and long-term side effects in a relatively young patient. Collaborative discussions with the multidisciplinary team, including medical oncologists and surgical oncologists, are essential to tailor the treatment plan to the patient's specific needs and circumstances. |

<table>
<tr>
<td>2<br><br>Accurate; regional nodal irradiation is suggested</td>
<td>
Memory updated<br><br>
For a 40-year-old patient with invasive ductal carcinoma who has undergone a modified radical mastectomy and axillary lymph node dissection, with five of ten lymph nodes positive, adjuvant radiation therapy is strongly recommended. Here's a tailored approach based on her pathology:<br><br>
1. Breast/Chest Wall Radiation: Given the mastectomy, chest wall radiation would be indicated to reduce the risk of locoregional recurrence. This would typically involve a course of radiation over 5-6 weeks, targeting the chest wall and including any surgical scar with adequate margins.<br><br>
2. Regional Lymph Node Radiation: The involvement of axillary lymph nodes (5/10 positive nodes) suggests a significant risk for regional recurrence. Radiation should encompass the supraclavicular fossa and may also include the axillary and infraclavicular nodes. Consideration for the inclusion of the internal mammary nodes should be based on the location of the tumor and patterns of lymphatic drainage.<br><br>
3. Dose and Fractionation: Standard fractionation (e.g., 50 Gy in 25 fractions) can be used. However, hypofractionated schedules might be suitable and should be considered based on the patient's overall treatment plan and specific risk factors.<br><br>
4. Consideration of Boost Doses: While boost doses to the chest wall are less commonly employed compared to breast-conserving therapy, they may be considered if there are high-risk features such as close margins or extensive lymphovascular invasion, though the latter is not present in this case.<br><br>
5. Integration with Systemic Therapy: Coordination with medical oncology is crucial to optimize the timing of radiation therapy relative to any adjuvant systemic therapies that might be indicated (e.g., hormonal therapy given the hormone receptor-positive status).<br><br>
6. Follow-Up and Monitoring: Regular follow-up with imaging and clinical examination is necessary to monitor for signs of recurrence and manage any side effects of treatment.<br><br>
This plan should be tailored further based on the patient's overall health, preferences, and any other [illegible]
</td>
</tr>
</table>

| | |
|---|---|
| 3<br><br>Accurate; regional nodal irradiation is suggested | Memory updated<br><br>For a 40-year-old woman with the described breast cancer pathology and surgical history, adjuvant radiation therapy is a recommended component of treatment to reduce the risk of local recurrence, particularly given the involvement of lymph nodes.<br><br>Here are the considerations for radiation treatment:<br><br>1. **Chest Wall Radiation:** Given the mastectomy, radiation to the chest wall is advisable due to the number of positive lymph nodes. This will help manage any residual microscopic disease.<br><br>2. **Regional Lymph Node Irradiation (RLNI):** With five out of ten nodes involved, irradiation of the regional lymph nodes, including the axillary, supraclavicular, and possibly the internal mammary nodes, is strongly recommended. This targets potential residual cancer cells in the lymphatic pathways.<br><br>3. **Technique and Dosing:**<br>• Typically, a dose of about 50 Gy in 25 fractions over five weeks is used for the chest wall and regional nodes.<br>• Techniques such as intensity-modulated radiation therapy (IMRT) or volumetric modulated arc therapy (VMAT) can be employed to optimize the dose distribution and minimize exposure to adjacent structures like the heart and lungs, especially important on the left side if applicable.<br><br>4. **Consideration of Boost:** Depending on surgical findings and oncologist's assessment, a boost to any areas of particular concern, such as close or positive margins (if they had been present), might be considered.<br><br>5. **Systemic Therapy Integration:** Coordination with medical oncology to discuss the timing of radiation relative to any adjuvant systemic therapy, such as endocrine therapy given the hormone receptor-positive status, is crucial.<br><br>6. **Follow-Up and Monitoring:** Regular follow-ups will be necessary to monitor for side effects of radiation and signs of recurrence, along with ongoing systemic therapy management.<br><br>Each element of this radiation treatment plan would be tailored to the patient's specific case details, overall health, and preferences, ensuring a comprehensive approach to her post-surgical cancer care. |

<table>
<tr>
<td>4<br><br>Accurate; regional nodal irradiation is suggested</td>
<td>Memory updated<br><br>
For your patient with invasive ductal carcinoma, ER/PR positive, HER2 negative, the presence of five out of ten lymph nodes positive for macrometastases suggests a high risk of locoregional recurrence, which supports the use of adjuvant radiation therapy. Here's a guideline-based approach for adjuvant radiation recommendations:<br><br>
1. Chest Wall Radiation: Given the mastectomy and node-positive disease, radiation to the chest wall is indicated to reduce the risk of local recurrence. This typically includes treatment of the chest wall and any remaining axillary tissue, especially if there was extensive nodal involvement.<br><br>
2. Regional Lymph Node Radiation: In cases with multiple positive lymph nodes, radiation should extend to include the regional lymph nodes. This includes the supraclavicular (SCV), axillary, and internal mammary nodes (IMNs), which are commonly included in the radiation field to address potential microscopic disease.<br><br>
3. Technique and Dosing:<br>
• Conventional Fractionation: Typically, a dose of 50 Gy in 25 fractions is used for the chest wall and regional lymph nodes.<br>
• Hypofractionation: For patients who can be treated with a shorter course of radiation, hypofractionated regimens may be considered, especially if the patient prefers a shorter treatment duration for convenience or other reasons.<br><br>
4. Considerations for Boost: If there are high-risk features such as close margins or very aggressive tumor biology (suggested by the high Ki-67), a boost to the mastectomy scar might be considered, typically an additional 10-16 Gy in fractions of 2 Gy.<br><br>
5. Ongoing Evaluation: It's important to monitor for any signs of lymphedema, especially with comprehensive nodal irradiation, and manage them proactively.<br><br>
6. Multidisciplinary Discussion: Review the patient's overall health status and any potential comorbidities that might affect her suitability for radiation or necessitate adjustments in the radiation plan.<br><br>
Given the patient's young age and the high Ki-67 indicating a more proliferative tumor, these recommendations aim to minimize the risk of recurrence while considering the long-term side effects of treatment. Ensure to tailor the approach based on further inputs from the surgical and medical oncology teams.</td>
</tr>
</table>

<table>
<tr>
<td>5<br><br>Accurate; regional nodal irradiation is suggested</td>
<td>Memory updated<br><br>For a 40-year-old woman with this clinical profile following a modified radical mastectomy, adjuvant radiation therapy is strongly indicated given the positive nodal status. Here's a tailored recommendation:<br><br>1. Whole Breast/Chest Wall Radiation: If any breast tissue remains or if she has undergone reconstruction, radiation should be considered to the chest wall and any residual breast tissue to reduce locoregional recurrence. For the chest wall, typical doses would be 50 Gy in 25 fractions.<br><br>2. Regional Nodal Irradiation: Given that five out of ten nodes were positive, regional nodal irradiation (RNI) is recommended. This should include the axillary, supraclavicular, and internal mammary nodes. RNI has been shown to improve both disease-free and overall survival in patients with involved axillary lymph nodes.<br><br>3. Dose Considerations: An additional boost to the surgical scar area could be considered, particularly if there are any high-risk features not clearly addressed by the standard radiation dose.<br><br>4. Discussion Points:<br>• The potential benefits of radiation in reducing local and regional recurrence.<br>• Possible side effects, including lymphedema, especially given the axillary node dissection, skin changes, and potential impacts on cardiac and pulmonary tissues depending on the technique and area irradiated.<br>• The importance of multidisciplinary care, including ongoing systemic therapy considerations based on hormone receptor status and high Ki-67.<br><br>Ensure a thorough discussion with the patient regarding the balance between the benefits and risks of radiation therapy in her specific case. Additionally, consider genetic counseling given her age and cancer characteristics, as this may influence her long-term management plan.</td>
</tr>
</table>

Third Example (ChatGPT 4)

3. 80 year old woman with new diagnosis of invasive ductal carcinoma, ER/PR positive, HER2 negative status post modified radical mastectomy and axillary lymph node dissection, final pathology demonstrating 1.3cm carcinoma, well differentiated, no lymphovascular invasion, margins negative by >2mm, Ki-67 50%, and zero of ten lymph nodes positive for macrometastases. Presenting for adjuvant radiation recommendations:
**Expected response: No radiation**

### General Observations

Original Prompt:
-The model displays notable and appropriate hesitancy when considering adjuvant radiation therapy, but often simply emphasizes the importance of making an

personalized decision considering the patient's preferences, values, and other factors that may affect her health
-In all responses, the model erroneously implies or states that adjuvant radiation may be a consideration for the patient
-0/5 accurate
-1/5 partially accurate

-Accuracy improved notably after alternative prompting

Phrasing 1:
-1/5 accurate
-4/5 partially accurate

Phrasing 2:
-5/5 accurate

Phrasing 3:
-0/5 accurate
-5/5 inaccurate

Chain of Reasoning Prompting:
-5/5 accurate
-In two chats, the model correctly answered the original prompt without alternative wording

<table>
<tr>
<td>1<br><br>Inaccurate; omitting adjuvant radiation is not suggested as the only viable option</td>
<td>For an 80-year-old woman with a new diagnosis of ER/PR positive, HER2 negative invasive ductal carcinoma who has undergone a modified radical mastectomy and axillary lymph node dissection, with final pathology showing a 1.3 cm well-differentiated tumor, no lymphovascular invasion, clear margins, a high Ki-67 of 50%, and no lymph node involvement, the decision on adjuvant radiation therapy needs careful consideration of her overall health, life expectancy, and the balance between potential benefits and side effects of radiation.<br><br>Here are some considerations for adjuvant radiation recommendations:<br><br>1. Life Expectancy and Comorbidities: At 80 years old, it's crucial to evaluate her overall health and any comorbid conditions. If she has a good life expectancy and is in relatively good health, adjuvant radiation may be considered to decrease local recurrence risks, particularly given the high proliferation index (Ki-67) which suggests a more aggressive tumor behavior.<br><br>2. Tumor Characteristics and Surgical Outcomes: The tumor size is relatively small, and the surgical margins are negative, which are favorable factors. The lack of lymphovascular invasion and negative lymph nodes are also positive indicators that diminish the necessity for radiation in terms of systemic spread.<br><br>3. Radiation Side Effects: Consideration of the potential side effects of radiation is essential, especially in older adults who may be more susceptible to the skin and soft tissue effects, fatigue, and possibly impacts on cardiac and pulmonary functions, depending on the radiation field.<br><br>4. Guidelines and Recommendations: According to various oncology guidelines, adjuvant radiation therapy is often recommended for breast cancer patients with higher risk factors such as positive margins or lymph node involvement, which are not present in this case. However, the high Ki-67 might tilt the balance towards considering radiation due to its association with higher tumor aggressiveness.<br><br>5. Patient Preference: It is also vital to discuss the potential benefits and drawbacks of radiation therapy with the patient, considering her personal values and preferences regarding quality of life and treatment burden.<br><br>Given these factors, a thorough discussion with a multidisciplinary team, including medical oncologists, radiation oncologists, and possibly geriatric specialists, would be appropriate to tailor the best treatment plan for her. Additionally, considering her age and the specifics of the tumor, the decision to forego radiation could be reasonable if the treatment's potential risks and impacts outweigh the benefits in her particular situation.</td>
</tr>
</table>

| | |
|---|---|
| 2<br><br>Inaccurate; omitting adjuvant radiation is not suggested as the only viable option | For an 80-year-old woman with the described pathology after a modified radical mastectomy and axillary lymph node dissection, the radiation recommendation can be individualized based on several factors, including her overall health, life expectancy, and preferences.<br><br>Given the pathology:<br>• Invasive ductal carcinoma, 1.3 cm<br>• Well-differentiated<br>• No lymphovascular invasion<br>• Margins clear by more than 2mm<br>• High Ki-67 of 50% (suggesting a higher proliferation rate)<br>• No lymph node involvement<br><br>Here are some considerations for adjuvant radiation therapy:<br><br>1. **Breast Radiation**: Generally, for older patients with small, node-negative, and well-differentiated tumors, the benefit of whole breast irradiation might be marginal, especially if systemic therapy (like hormonal therapy) is planned. The high Ki-67, however, does suggest a more aggressive tumor behavior, which could tip the balance in favor of recommending radiation to reduce local recurrence risk.<br>2. **Chest Wall Radiation**: Since she has undergone a mastectomy, radiation to the chest wall may be considered if there were higher risk features (like close margins or lymphovascular invasion), which she does not have. Given her age and the favorable pathology, omitting chest wall radiation could be reasonable unless there are specific patient or tumor factors not mentioned that increase risk.<br>3. **Regional Nodal Irradiation**: As there is no lymph node involvement, regional nodal irradiation would typically not be indicated in this scenario.<br>4. **Hormonal Therapy**: Given the ER/PR positive status, hormonal therapy will likely be the cornerstone of her adjuvant treatment. The decision to add radiation could also depend on her response and tolerance to hormonal therapy.<br>5. **Patient Preference and Life Expectancy**: Consideration of her quality of life and life expectancy is crucial. Discussions about the potential benefits versus the side effects of radiation are important, particularly in the context of her age and any comorbid conditions.<br><br>In conclusion, if she is in good health with a reasonable life expectancy, discussing the potential benefits of chest wall radiation to reduce the risk of local recurrence might be warranted due to the high Ki-67. However, the decision should be made after thorough discussion with the patient about the potential benefits and drawbacks, considering her personal values and goals of care. |

| | |
|---|---|
| 3<br><br>Partially accurate; whilst omitting adjuvant radiation is recommended, the model implies that radiotherapy may be a consideration | Memory updated<br><br>For this 80-year-old patient with ER/PR positive, HER2 negative invasive ductal carcinoma, having undergone a modified radical mastectomy and axillary lymph node dissection with favorable pathology, the decision on adjuvant radiation primarily considers the balance between the potential benefits and the risks associated with radiation therapy, particularly in the context of her age and the specifics of the tumor.<br><br>**Key Considerations:**<br><br>1. **Age and Comorbidities:** Given her age, it's important to consider her overall health status, potential life expectancy, and how comorbidities might affect her tolerance to radiation therapy.<br>2. **Pathology Outcomes:** The absence of lymph node involvement, well-differentiated histology, and clear margins are favorable factors that reduce the recurrence risk. The Ki-67 index at 50% indicates a higher proliferative rate, suggesting a more aggressive tumor behavior which could somewhat elevate the recurrence risk.<br>3. **Risk vs. Benefit:** The main benefit of radiation therapy in this context would be reducing the risk of local recurrence. However, the absence of nodal involvement and clean surgical margins significantly lower this risk. The side effects of radiation, including potential cardiac toxicity, especially relevant in left-sided breast cancers, and the risk of lymphedema, need to be weighed against its potential benefits.<br><br>**Recommendations:**<br><br>• **Omit Radiation:** Considering her age, the favorable pathology, and the potential side effects of radiation, she might be a candidate to safely omit adjuvant radiation, especially if her life expectancy is considered limited by other health factors.<br>• **Endocrine Therapy:** Since the tumor is ER/PR positive, endocrine therapy would likely be beneficial and less burdensome in terms of side effects compared to radiation, improving systemic control and possibly providing a survival benefit.<br>• **Patient Preferences and Quality of Life:** Discuss the potential benefits and drawbacks of radiation therapy with the patient, taking into account her values and preferences regarding quality of life and treatment burden.<br><br>It would be prudent to have a detailed discussion with the patient about the expected benefits of radiation in her specific case, considering her personal health goals and potential for side effects. This decision should ideally be made in a multidisciplinary team setting, involving medical oncology, radiation oncology, and possibly geriatric oncology, to ensure a decision that best aligns with the patient's overall health status and treatment preferences. |

<table>
<tr>
<td>4<br><br>Inaccurate; omitting adjuvant radiation is not suggested as the only viable option</td>
<td>

Given the clinical scenario you've described for an 80-year-old woman with invasive ductal carcinoma, ER/PR positive, HER2 negative, the decision regarding adjuvant radiation therapy should be individualized, especially considering her age and the favorable pathology of her tumor.

#### Key Points to Consider:

1. Tumor Characteristics:
   - **Size and Differentiation:** The tumor is relatively small (1.3 cm) and well-differentiated.
   - **Ki-67 Index:** The Ki-67 index is 50%, which suggests a higher proliferative rate and might typically argue for more aggressive treatment. However, this needs to be balanced with her age and overall health.
   - **Lymph Node Status:** No lymph node involvement is a favorable factor that reduces the risk of locoregional recurrence.
   - **Margins:** Negative surgical margins greater than 2mm also favor a lower risk of local recurrence.
2. Patient Age and Comorbidities:
   - Consideration of the patient's overall health, life expectancy, and potential to tolerate the side effects of radiation therapy is crucial. At 80 years old, the patient may have other comorbid conditions that could impact the decision-making process.
3. Radiation Therapy Benefits and Risks:
   - **Benefits:** Adjuvant radiation can decrease the risk of local recurrence, which might be particularly important given the high Ki-67 index.
   - **Risks:** Potential side effects include skin irritation, fatigue, and, rarely, more severe effects such as rib fractures or cardiac issues, especially relevant in an elderly population.

#### Recommendations:

- **Discuss with the Patient:** Engage in a thorough discussion with the patient about the potential benefits and risks of radiation therapy, considering her personal values and goals of care.
- **Multidisciplinary Team Consultation:** Consultation with a multidisciplinary team including medical oncology, geriatrics, and possibly a radiation oncology specialist could provide a balanced perspective.
- **Decision Aids:** Use decision aids that can incorporate patient-specific factors and preferences to help in making an informed choice.

#### Alternative Considerations:

- If the patient opts against radiation therapy, careful follow-up with regular clinical and possibly imaging evaluations could be a reasonable approach given the favorable pathology, though the high Ki-67 might necessitate a more vigilant follow-up schedule.

Ultimately, the decision should align with the patient's preferences and overall health status, with a clear communication of the trade-offs involved in choosing or foregoing adjuvant radiation therapy.

</td>
</tr>
</table>

| | |
|---|---|
| 5<br><br>Inaccurate; omitting adjuvant radiation is not suggested as the only viable option | For an 80-year-old woman with the described pathology following a modified radical mastectomy and axillary lymph node dissection, the decision to recommend adjuvant radiation therapy depends on various factors including her overall health, life expectancy, and specific characteristics of the cancer.<br><br>1. **Tumor Characteristics and Risk Factors:**<br>• **Tumor Size and Differentiation:** The tumor size is relatively small (1.3 cm) and well-differentiated, suggesting a lower grade cancer.<br>• **Ki-67 Index:** The Ki-67 of 50% indicates a high proliferative rate, which is generally associated with a higher risk of recurrence.<br>• **Lymph Node Status:** No lymph node involvement is a favorable prognostic factor reducing the likelihood of regional recurrence.<br>• **Margins:** Negative surgical margins greater than 2mm also lower the risk of locoregional recurrence.<br><br>2. **Role of Adjuvant Radiation:**<br>• **Life Expectancy and Comorbidities:** Given her age, the decision should also weigh her life expectancy and any comorbid conditions which might impact the benefit from radiation therapy.<br>• **Risk of Recurrence:** Despite the high Ki-67, the absence of lymph node involvement and clear margins might suggest a lower overall risk of recurrence. However, the high Ki-67 could still tip the balance towards considering radiation due to its association with aggressive tumor behavior.<br><br>3. **Patient Preferences and Quality of Life:**<br>• **Potential Benefits vs. Side Effects:** It's crucial to discuss the potential benefits of reducing the risk of local recurrence against the possible side effects of radiation, which may include fatigue, skin changes, and rare but serious effects such as cardiac or pulmonary complications.<br>• **Patient Values and Goals of Care:** Understanding her values and goals of care is essential. Some patients may prioritize maximal therapy aimed at reducing recurrence risk, while others might prioritize quality of life without undergoing radiation.<br><br>4. **Guideline Considerations:**<br>• Guidelines often suggest considering radiation therapy in patients with high-risk factors despite having clear margins and no lymph node involvement, particularly with a high Ki-67 index. However, they also emphasize individualized decision-making based on patient-specific factors and preferences.<br><br>In conclusion, a discussion with the patient about the benefits and risks of adjuvant radiation therapy, taking into account her personal health goals and potential for improved outcomes, is recommended. Additionally, consulting multidisciplinary team opinions including medical oncologists could provide further personalized guidance. |

| | |
|---|---|
| Phrasing 1 | What specific kind of treatment should be provided for an 80 year old woman with new diagnosis of invasive ductal carcinoma, ER/PR positive, HER2 negative status post modified radical mastectomy and axillary lymph node dissection, final pathology demonstrating 1.3cm carcinoma, well differentiated, no lymphovascular invasion, margins negative by >2mm, Ki-67 50%, and zero of ten lymph nodes |

| | |
|---|---|
| | positive for macrometastases presenting for adjuvant radiation recommendations? |
| 1<br><br>Accurate; adjuvant radiation therapy is not recommended | For an 80-year-old woman with a diagnosis of ER/PR positive, HER2 negative invasive ductal carcinoma who has undergone a modified radical mastectomy and axillary lymph node dissection, the recommendation for adjuvant radiation therapy can be influenced by several factors from her pathology report and overall health status.<br><br>Given the details you provided:<br>• Tumor size is 1.3 cm, well-differentiated.<br>• No lymphovascular invasion is present.<br>• Margins are clear with more than 2mm.<br>• Ki-67 is 50%, indicating a higher proliferation rate.<br>• No lymph nodes involved out of ten sampled.<br><br>**Recommendations:**<br>1. **Consideration of Radiation Therapy:**<br>• **Breast Radiation:** Typically, adjuvant whole breast radiation is not indicated after mastectomy unless there are high-risk features like close/positive margins, lymphovascular invasion, or lymph node involvement, which are not present in this case.<br>• **Chest Wall Radiation:** Given the patient's age, the small size of the tumor, and the lack of high-risk features, routine radiation to the chest wall is generally not indicated. However, the high Ki-67 might prompt a discussion about the benefits versus the risks of radiation given its association with more aggressive tumor behavior.<br>2. **Systemic Therapy:**<br>• **Hormonal Therapy:** Since the tumor is ER/PR positive, adjuvant hormonal therapy with an aromatase inhibitor or tamoxifen should be strongly considered, as it can significantly reduce the risk of recurrence, especially in hormone receptor-positive breast cancers.<br>3. **Patient's Overall Health and Preferences:**<br>• The decision for additional treatments such as radiation should also take into account the patient's comorbidities, life expectancy, and quality of life considerations. Given her age, it is crucial to balance the potential benefits of radiation with the risks and her personal values and preferences.<br>4. **Multidisciplinary Consultation:**<br>• It's important to have a multidisciplinary team discussion, including medical oncology, radiation oncology, and surgical oncology, to tailor the treatment plan based on the patient's overall health status and treatment preferences.<br><br>**Conclusion:**<br>For this specific patient, unless there are additional risk factors not mentioned (such as genetic predispositions or other pathological high-risk features), routine adjuvant radiation to the chest wall may not be necessary based on the current guidelines and evidence. Hormonal therapy should be a key component of her adjuvant treatment. Each case should be considered individually, and patient preferences should always be integrated into the final treatment decision. |

| | |
|---|---|
| 2<br><br>Partially Accurate; omitting adjuvant radiation is suggested, but no decisive recommendation was made | In an 80-year-old woman with invasive ductal carcinoma, ER/PR positive, HER2 negative, who has undergone a modified radical mastectomy with clear margins, and no lymphovascular invasion or lymph node involvement, the decision to proceed with adjuvant radiation therapy should be individualized based on her overall health status, life expectancy, and personal preferences.<br><br>Given her age and the favorable pathological features of her tumor (well-differentiated, no lymphovascular invasion, negative margins, and no nodal involvement), she might have a relatively low risk of local recurrence. The high Ki-67 of 50%, however, suggests a higher proliferation rate, which is typically associated with a more aggressive tumor behavior and could argue for more aggressive treatment including adjuvant radiation therapy to reduce the risk of local recurrence.<br><br>The benefits of adjuvant radiation therapy in elderly patients must be weighed against the potential side effects and the impact on quality of life. In breast cancer patients over the age of 70, especially those with ER-positive tumors treated with adjuvant hormonal therapy, the omission of radiation therapy after mastectomy can be considered if they are at low risk for recurrence based on the pathology.<br><br>Here are considerations and possible recommendations:<br>1. **Discuss Goals and Preferences**: It is important to have a detailed discussion with the patient about her goals of care and her preferences regarding quality of life and potential treatment-related side effects.<br>2. **Consider Comorbidities:** Assess her overall health and any comorbid conditions that might affect her tolerance of additional therapy.<br>3. **Adjuvant Systemic Therapy**: Given the hormone receptor-positive status, adjuvant endocrine therapy (such as tamoxifen or an aromatase inhibitor) should be strongly considered to reduce the risk of distant recurrence.<br>4. **Radiation Therapy Decision**: If she is in excellent health and is motivated to minimize the risk of local recurrence, discussing the potential benefits and risks of radiation therapy is appropriate. However, if her life expectancy is limited due to age or comorbidities, the addition of radiation therapy might not provide a substantial benefit.<br><br>Ultimately, the decision should be made collaboratively, considering the patient's values and the clinical judgment of the healthcare team. |

<table>
<tr>
<td>3<br><br>Partially accurate; omitting adjuvant radiation is suggested, but no decisive recommendation was made</td>
<td>
For an 80-year-old woman with a new diagnosis of invasive ductal carcinoma, ER/PR positive, HER2 negative, who has undergone a modified radical mastectomy and axillary lymph node dissection with favorable pathology outcomes (no lymphovascular invasion, negative margins, no macrometastases in lymph nodes), the decision to pursue adjuvant radiation therapy needs to be carefully considered based on her overall health, life expectancy, and personal preferences.
Here's a breakdown of treatment considerations:
<ol>
<li>Radiation Therapy: Generally, adjuvant radiation therapy after mastectomy in breast cancer is considered if there are high-risk features such as positive lymph nodes, close or positive margins, or large tumor size. In this case, where the tumor is relatively small (1.3 cm), well-differentiated, and all other prognostic markers are favorable except for a high Ki-67 (50%), the benefit of radiation therapy might be limited, particularly given her advanced age and the absence of lymph node involvement.</li>
<li>Ki-67 Index: A high Ki-67 index indicates a higher proliferation rate, suggesting a more aggressive tumor biology. This might lean towards a more aggressive adjuvant treatment strategy. However, in the absence of other high-risk features and considering the patient's age, the impact of this single factor might be mitigated.</li>
<li>Systemic Therapy: Given the hormone receptor-positive status of the tumor, adjuvant hormonal therapy (e.g., aromatase inhibitors or tamoxifen) would likely be beneficial in reducing the risk of recurrence. This treatment is generally well-tolerated, especially important in older patients, and can be an effective adjuvant treatment with a favorable side effect profile compared to chemotherapy or radiation.</li>
<li>Patient's Health and Preferences: It's crucial to consider the patient's overall health status, comorbidities, and personal values and preferences. Discussions about the risks and benefits of additional radiation therapy, potential side effects, and the impact on quality of life are essential.</li>
</ol>
In conclusion, considering her age and the pathology report's favorable findings, a strong consideration could be given to systemic hormonal therapy while potentially omitting adjuvant radiation, unless other individual factors weigh towards a more aggressive approach. Each decision should be personalized, ideally discussed in a multidisciplinary team setting and with the patient's active involvement.
</td>
</tr>
<tr>
<td>4<br><br>Partially accurate; the LLM leans towards omitting adjuvant radiation, but is indecisive until further prompting</td>
<td>
In the scenario you've described—an 80-year-old woman with invasive ductal carcinoma that is ER/PR positive, HER2 negative, with a favorable pathological report post-surgery—the decision for adjuvant radiation would be based on a balance of potential benefits against the risks and the patient's overall health status and preferences.
Here are key considerations for adjuvant therapy in this case:
<ol>
<li>Patient Age and Comorbidities: At 80 years old, it's crucial to consider the patient's life expectancy and any comorbid conditions that might impact the risks and benefits of further treatment, such as radiation therapy.</li>
<li>Pathological Features:
<ul>
<li>Tumor Size and Differentiation: The tumor is relatively small (1.3 cm) and well-differentiated, suggesting a lower risk of recurrence.</li>
<li>Lymph Node Status: No lymph node involvement is a significant positive prognostic factor.</li>
<li>Margins: Clear margins (>2 mm) reduce the likelihood of local recurrence.</li>
<li>Ki-67 Index: A higher Ki-67 (50%) indicates a higher proliferation rate, suggesting a more aggressive tumor biology. This could tilt considerations towards offering adjuvant therapy to reduce the risk of recurrence despite other favorable factors.</li>
</ul>
</li>
<li>ER/PR Positive, HER2 Negative Status: Hormone receptor-positive status would typically prompt endocrine therapy, which could be sufficient as the primary adjuvant treatment, given the overall lower aggressiveness implied by the other factors but countered somewhat by the high Ki-67.</li>
</ol>
</td>
</tr>
</table>

| | |
|---|---|
| | 4. **Radiation Therapy Considerations**:<br>• In older patients, especially those over 70, radiation therapy after breast-conserving surgery is sometimes omitted due to the lower absolute benefit in non-life-threatening recurrence risk reduction and the quality of life considerations.<br>• Given that she had a mastectomy, the indication for radiation would depend more on factors like a larger tumor size, close/positive margins, or lymph node involvement, most of which are not present in this case.<br>5. **Guidelines and Recommendations**:<br>• National and international guidelines (like those from the NCCN or ASCO) could provide further tailored recommendations based on the combination of risk factors.<br>• Decision aids or predictive models like Adjuvant! Online might help quantify the benefits and risks of adjuvant radiation in her specific scenario.<br>6. **Patient Preferences**: Understanding the patient's values and preferences regarding quality of life versus the potential for slightly extended survival or reduced recurrence risk is paramount.<br><br>Given these factors, the inclination might lean towards recommending against adjuvant radiation therapy, particularly if the patient values quality of life over a marginal increase in recurrence prevention, focusing instead on systemic therapy like endocrine treatment. However, a thorough discussion with the patient about the potential benefits and side effects of radiation, along with a multidisciplinary team consultation, would be essential to make a tailored decision.<br><br>Follow-up question: If you had to choose, what adjuvant radiation recommendation would you go with?<br><br>Given the details you've provided about the case—an 80-year-old woman with a relatively small, well-differentiated invasive ductal carcinoma, no lymph node involvement, clear surgical margins, and a high Ki-67 index—the decision on adjuvant radiation would pivot primarily on the patient's overall health status, life expectancy, and the high proliferative index of the tumor.<br><br>Considering that:<br>• The patient's age and likely comorbidities may impact her tolerance and the benefits she could derive from radiation therapy.<br>• She has favorable factors like negative lymph nodes and clear margins which generally lessen the necessity for adjuvant radiation.<br>• The high Ki-67 index indicates a more aggressive tumor biology, which is a factor leaning towards recommending adjuvant therapies to reduce the risk of recurrence.<br><br>**Recommendation**: I would lean towards not recommending routine adjuvant radiation in this scenario, primarily due to her age, the surgical success with clear margins and no lymph node involvement, and considering the balance of improving quality of life against the modest potential benefits of radiation in this context. Instead, focusing on systemic adjuvant therapies like endocrine therapy would be more beneficial and have a significant impact on reducing recurrence risks with fewer side effects compared to radiation. This approach aligns with the principles of geriatric oncology, which emphasize tailoring cancer treatment to individual health status and patient preferences to maximize benefits while minimizing harms.<br><br>However, this recommendation would ideally be preceded by a thorough discussion with the patient about her preferences, values, and understanding of the trade-offs involved. Consulting with a multidisciplinary team would also ensure that all perspectives are considered before finalizing the treatment plan. |

<table>
<tr>
<td>5<br><br>Partially accurate; omitting adjuvant radiation is suggested (“Hormone Therapy Alone”) but no decisive recommendation was made</td>
<td>
For an 80-year-old woman with the described pathology from invasive ductal carcinoma, the decision to recommend adjuvant radiation therapy must be personalized based on her overall health status, life expectancy, and preferences, considering the risks and benefits of the treatment.<br><br>
Here's a breakdown of the factors and considerations:<br>
1. Tumor Characteristics and Surgery Outcomes:<br>
• Size and Differentiation: The tumor size (1.3 cm) is relatively small, and it is well differentiated, suggesting a slower-growing cancer.<br>
• Lymph Node Involvement: No lymph nodes are involved, which typically indicates a lower risk of regional recurrence.<br>
• Margins: Negative margins (>2mm) suggest that the surgical resection was successful in removing the visible tumor.<br>
• Ki-67 Index: A high Ki-67 index (50%) indicates a higher proliferative rate of the cancer cells, which generally correlates with a higher risk of recurrence. This factor might argue for more aggressive adjuvant therapies.<br>
2. Hormonal Status:<br>
• ER/PR Positive: The positive hormone receptor status suggests that the tumor will respond to hormonal therapies, which can be a significant part of adjuvant treatment to reduce the risk of recurrence.<br>
3. Adjuvant Radiation Therapy Considerations:<br>
• Age and Comorbidities: At 80 years old, consideration of the patient's overall health, potential side effects, and life expectancy is crucial. Adjuvant radiation therapy can lead to improved local control, but the benefits must be weighed against the potential for acute and chronic side effects, which might impact her quality of life.<br>
• Benefits of Radiation: For some older patients, particularly those who are healthy and have a good life expectancy, radiation can offer significant benefits in terms of reducing local recurrence. However, in a case with favorable pathology and successful surgical outcomes, the absolute benefit might be small.<br>
4. Alternative Strategies:<br>
• Hormonal Therapy Alone: Given her ER/PR positive status, hormonal therapy could be the primary adjuvant treatment. This approach would help manage systemic risks of recurrence without the local risks associated with radiation.<br>
5. Patient Preference: It's essential to engage the patient in discussions about the pros and cons of additional treatments like radiation. Her values and preferences regarding quality of life and treatment burden should guide the decision-making process.<br><br>
Recommendations:<br>
• Discuss with the Patient: Review the potential benefits and drawbacks of radiation therapy in the context of her overall health status and personal preferences.<br>
• Consider Hormonal Therapy: As she is ER/PR positive, hormonal therapy (like tamoxifen or an aromatase inhibitor, depending on her menopausal status) should be strongly considered as it could provide significant benefit in reducing the risk of recurrence.<br>
• Multidisciplinary Team Consultation: It would be beneficial to discuss her case in a tumor board setting or with a multidisciplinary team to integrate various perspectives, including medical oncology and geriatrics, if available.<br><br>
This approach ensures that all aspects of her health, pathology results, and personal preferences are considered in the treatment planning.
</td>
</tr>
</table>

| | |
|---|---|
| Phrasing 2 | What would NCCN category 1 recommend for an 80 year old woman with new diagnosis of invasive ductal carcinoma, ER/PR positive, HER2 negative status post modified radical |

| | |
|---|---|
| | mastectomy and axillary lymph node dissection, final pathology demonstrating 1.3cm carcinoma, well differentiated, no lymphovascular invasion, margins negative by >2mm, Ki-67 50%, and zero of ten lymph nodes positive for macrometastases presenting for adjuvant radiation recommendations? |
| 1<br><br>Accurate; omitting adjuvant radiation is recommended | For an 80-year-old woman with a diagnosis of invasive ductal carcinoma that is ER/PR positive and HER2 negative, with the characteristics described in the pathology report, the NCCN (National Comprehensive Cancer Network) guidelines would consider several factors to make a recommendation regarding adjuvant radiation therapy.<br><br>Given the pathology specifics:<br>• Tumor size is 1.3 cm (T1)<br>• Well-differentiated<br>• No lymphovascular invasion<br>• Margins are negative with a clearance of >2 mm<br>• Ki-67 is 50%, suggesting a higher proliferation rate<br>• Zero of ten lymph nodes positive for metastases<br><br>For a patient who has undergone a modified radical mastectomy, the NCCN guidelines generally consider post-mastectomy radiation therapy (PMRT) based on a combination of factors including the size of the tumor, lymph node involvement, and other pathological features that might increase the risk of recurrence.<br><br>Here are key considerations based on the current scenario:<br>1. **Age and General Health:** At 80 years old, the overall health status and life expectancy of the patient play crucial roles in the decision-making process. If the patient has a good performance status and a life expectancy that justifies adjuvant treatments, recommendations might lean more aggressively. However, for many older patients, the side effects and potential morbidity associated with radiation might outweigh the benefits, particularly when the tumor characteristics suggest a lower risk of local recurrence.<br>2. **Tumor Characteristics and Lymph Node Status:** The small tumor size (1.3 cm), absence of lymph node involvement, and clear margins generally suggest a lower risk of local recurrence. This might argue against the need for PMRT. However, the relatively high Ki-67 suggests a higher biological aggressiveness, which complicates the decision.<br>3. **Guideline-Specific Recommendations:** NCCN guidelines for a patient with these specific characteristics (especially being node-negative and having had a mastectomy with clear margins) typically do not recommend routine PMRT unless there are additional high-risk features. In this case, the only high-risk feature noted is the high Ki-67, which might not be sufficient alone to warrant PMRT.<br>4. **Patient Preference:** Discussing the potential benefits and drawbacks of radiation therapy, considering the patient's personal values and preferences regarding quality of life and potential treatment-related side effects, is essential.<br><br>Therefore, based on the NCCN Category 1 recommendations and the specifics of the case, adjuvant radiation therapy might not be routinely recommended unless there are other individual factors or preferences that tilt the balance in favor of radiation. It's always important to personalize cancer care to the specific patient, taking into account all medical, personal, and social factors involved. |

<table>
<tr>
<td>2<br><br>Accurate; omitting adjuvant radiation is recommended</td>
<td>
For an 80-year-old woman with the described pathology from a modified radical mastectomy for invasive ductal carcinoma that is ER/PR positive and HER2 negative, the National Comprehensive Cancer Network (NCCN) Guidelines provide tailored recommendations based on several factors, including tumor size, grade, lymph node involvement, and other pathological features.<br><br>
Given the specific details you provided:<br>
• 1.3 cm carcinoma, well-differentiated<br>
• No lymphovascular invasion<br>
• Margins negative by >2 mm<br>
• Ki-67 at 50%<br>
• 0/10 lymph nodes positive for macrometastases<br><br>
Here are the NCCN guidelines considerations for adjuvant radiation therapy:<br>
1. Radiation after Mastectomy: Typically, post-mastectomy radiation therapy (PMRT) is considered for patients with tumors larger than 5 cm or with positive lymph node involvement. Since this patient has a smaller tumor size (1.3 cm) and no nodal involvement, PMRT would generally not be recommended based on size and nodal status alone.<br>
2. Age Considerations: For elderly patients, especially those aged 80 and above, the benefits of radiation need to be balanced with the potential risks and the patient's overall health status and life expectancy. Given her age and the favorable pathological features of the tumor (e.g., no lymphovascular invasion, negative margins, well-differentiated), the risks of radiation may outweigh the benefits, leading to a decision against recommending PMRT.<br>
3. Ki-67 High Proliferation Index: A Ki-67 of 50% indicates a high proliferative rate, which is typically associated with a higher risk of recurrence. However, in the context of other favorable factors and the absence of nodal involvement, this alone might not justify the use of PMRT but would be a consideration for systemic therapy.<br>
4. Systemic Therapy: Hormonal therapy would be recommended for this patient given the ER/PR positive status of the tumor. The high Ki-67 might also influence considerations regarding the addition of chemotherapy, which should be discussed based on her overall health and ability to tolerate treatment.<br>
5. Patient Preferences and Comorbidities: It's crucial to consider the patient's comorbidities, functional status, and preferences when deciding on adjuvant treatments, including radiation.<br><br>
In summary, based on the NCCN guidelines and the specifics of this case, routine adjuvant radiation therapy would not typically be recommended for this patient, focusing instead on systemic therapy options and supportive care tailored to her overall health status and treatment preferences. Always consult the most current NCCN guidelines or discuss with a multidisciplinary team for personalized treatment planning.
</td>
</tr>
</table>

<table>
<tr>
<td>3<br><br>Accurate; although indecisive, the LLM correctly interprets NCCN recommendations</td>
<td>

For an 80-year-old woman with the specified pathology of invasive ductal carcinoma, ER/PR positive, HER2 negative, post-modified radical mastectomy, and axillary lymph node dissection with the given characteristics, the NCCN (National Comprehensive Cancer Network) Guidelines provide recommendations based on the overall health status, life expectancy, and patient preferences, along with the specifics of the tumor pathology and surgical outcomes.

Given the details:

- Tumor size of 1.3 cm
- Well-differentiated
- No lymphovascular invasion
- Margins negative by >2mm
- Ki-67 of 50%, indicating a higher proliferation rate
- Zero of ten lymph nodes positive for macrometastases

For this patient, the decision for adjuvant radiation would be influenced by several factors:

1. **Age and Comorbidities:** At 80 years old, consideration of her overall health and potential life expectancy is crucial. If she is in good health with a life expectancy of more than 10 years, more aggressive treatments might be justified.
2. **Ki-67 Index:** A higher Ki-67 index (50%) suggests a more proliferative tumor, which typically correlates with a higher risk of recurrence. This could tilt the balance in favor of adjuvant therapies.
3. **Tumor Characteristics and Surgery Outcomes:** The small tumor size, well-differentiated nature, lack of lymphovascular invasion, and clear margins are all favorable factors that generally decrease the likelihood of recurrence.

**NCCN Guidelines:**

- For patients with T1-2, N0, M0 breast cancer who have undergone mastectomy, post-mastectomy radiotherapy (PMRT) is generally not recommended unless there are factors that increase the risk of locoregional recurrence.
- High-risk features that might prompt consideration of PMRT include close (<1 mm) or positive margins, extensive lymph node involvement (not present in this case), lymphovascular invasion (also not present here), and possibly a high Ki-67.

Given the lack of traditional high-risk features except for a high Ki-67, and considering the patient's age and presumably favorable life expectancy, the NCCN might categorize this scenario as one where adjuvant radiation could be considered but is not automatically recommended. The decision would ideally be made in a multidisciplinary setting, taking into account the patient's values and preferences regarding potential treatment benefits and side effects.

It would be beneficial to have a thorough discussion with the patient about the potential benefits and risks of adjuvant radiation in her specific case, including the potential for improvement in local control and the possible side effects of radiation.

</td>
</tr>
</table>

<table>
<tr>
<td>4<br><br>Accurate; omitting adjuvant radiation is recommended</td>
<td>For an 80-year-old woman with a new diagnosis of invasive ductal carcinoma, ER/PR positive, HER2 negative, post modified radical mastectomy and axillary lymph node dissection, with the specific pathological features you described, the NCCN guidelines provide tailored recommendations based on various factors, including the patient's age, overall health, and specific pathology details.<br><br>Based on the information provided:<br>• **Tumor size** is 1.3 cm, well-differentiated.<br>• **Ki-67 index** is high at 50%, indicating a higher proliferative rate.<br>• **Margins** are negative, greater than 2 mm.<br>• **Lymph node status** is negative, with zero of ten nodes positive for macrometastases.<br>• **No lymphovascular invasion** is noted.<br><br>**NCCN Recommendations:**<br>1. **Adjuvant Radiation Therapy:**<br>• Generally, adjuvant radiation therapy to the chest wall and regional lymph nodes might not be indicated in patients over 70 years if they have small (<5 cm), node-negative tumors, especially if surgical margins are clear and the tumor is well-differentiated.<br>• Given her age and the favorable pathological features (small tumor size, negative margins, no lymph node involvement), the recommendation could lean towards **not routinely offering adjuvant radiation therapy**, unless there are specific clinical concerns that might tilt the balance (e.g., high Ki-67 indicating higher tumor aggressiveness). However, the high Ki-67 could prompt a more thorough discussion about the potential benefits versus risks.<br>2. **Systemic Therapy:**<br>• **Endocrine Therapy:** Given the hormone receptor-positive status, adjuvant endocrine therapy (e.g., tamoxifen or an aromatase inhibitor) would be recommended to reduce the risk of recurrence.<br>3. **Follow-Up and Monitoring:**<br>• Regular follow-up with clinical exams and mammography is advised to monitor for recurrence.<br><br>**Clinical Judgment:**<br>In practice, decision-making for adjuvant treatment in elderly patients often incorporates a comprehensive discussion about the patient's life expectancy, comorbid conditions, preferences, and values regarding quality of life versus potential treatment-related side effects.<br><br>**Conclusion:**<br>No adjuvant radiation might be recommended based on the favorable features and her age, focusing instead on systemic therapy and regular monitoring. However, a detailed discussion involving the patient and her caregivers about the risks and benefits of all treatment options is crucial.</td>
</tr>
</table>

| | |
|---|---|
| 5<br><br>Accurate; omitting adjuvant radiation is recommended | For an 80-year-old woman with the described case of invasive ductal carcinoma (ER/PR positive, HER2 negative, status post modified radical mastectomy with no lymph node involvement, and well-differentiated tumor), the NCCN (National Comprehensive Cancer Network) guidelines would recommend as follows:<br><br>**Adjuvant Radiation Recommendation (NCCN Category 1)**<br><br>• **Post-mastectomy radiation therapy (PMRT):** According to NCCN guidelines, post-mastectomy radiation therapy (PMRT) is generally considered for patients with certain higher-risk factors, including larger tumors, positive lymph nodes, or close/positive margins.<br>• **In this case:**<br> • The tumor size is relatively small (1.3 cm).<br> • The tumor is well differentiated.<br> • There is no lymphovascular invasion.<br> • The margins are negative by more than 2 mm.<br> • No lymph node involvement (0/10 nodes positive).<br><br>Given these findings, **NCCN Category 1 guidelines would likely not recommend post-mastectomy radiation therapy (PMRT)** in this case, as there are no high-risk features like positive lymph nodes, large tumor size, or close margins.<br><br>However, other factors such as the high Ki-67 index (50%) might raise concerns about the tumor's aggressiveness, but this alone does not typically drive a Category 1 recommendation for radiation. The decision for radiation in such cases may still be personalized, considering the patient's overall health, life expectancy, and preferences.<br><br>In summary, for this particular case, **PMRT would not be recommended as per NCCN Category 1**, but consideration of other factors like Ki-67 should be discussed in a multidisciplinary setting. |

| | |
|---|---|
| Phrasing 3 | True or false: an 80 year old woman with new diagnosis of invasive ductal carcinoma, ER/PR positive, HER2 negative status post modified radical mastectomy and axillary lymph node dissection, final pathology demonstrating 1.3cm carcinoma, well differentiated, no lymphovascular invasion, margins negative by >2mm, Ki-67 50%, and zero of ten lymph nodes positive for macrometastases presenting for adjuvant radiation recommendations should receive no radiation. |
| 1<br><br>Inaccurate; omitting adjuvant radiation is not recommended | False. Although older age and certain favorable pathologic features (like small tumor size, no lymphovascular invasion, negative margins, and no lymph node involvement) might argue against the need for adjuvant radiation in some cases, the high Ki-67 index (50%) suggests a more aggressive tumor biology, which could potentially benefit from adjuvant radiation to reduce the risk of local recurrence.<br><br>In the scenario you described, decisions on adjuvant radiation therapy would typically consider patient's overall health, life expectancy, the specifics of tumor biology (like the high Ki-67), and personal preferences. The indication for radiation therapy in this context is nuanced and can vary based on these factors. It would be important to discuss the potential benefits and risks of radiation therapy in this particular patient with the care team and the patient herself to make a well-informed decision. |

| | |
|---|---|
| 2<br><br>Inaccurate; omitting adjuvant radiation is not recommended | False. While the decision to offer adjuvant radiation therapy in breast cancer involves multiple factors, there are key elements in this case that suggest considering radiation therapy.<br><br>For an 80-year-old woman with the described pathology, the decision to offer radiation therapy would typically depend on the balance between her overall health, life expectancy, and the potential benefits and risks of radiation. The tumor characteristics such as size, margin status, and lack of lymphovascular invasion are indeed favorable. However, the relatively high Ki-67 index of 50% indicates a higher proliferative activity, which can be associated with a higher risk of recurrence.<br><br>Current guidelines suggest that adjuvant radiation therapy is generally considered in patients who have undergone mastectomy if the tumor is greater than 5 cm in size or if there are positive lymph nodes. However, in cases where there are high-risk features such as a high Ki-67, even with no lymph node involvement, a discussion about the benefits and risks of radiation therapy would be prudent.<br><br>It is also important to assess the patient's comorbidities, functional status, and preferences when making a treatment decision. A multidisciplinary team approach and a thorough discussion with the patient about the potential benefits of reducing local recurrence versus the risks and side effects of radiation therapy would guide the best individualized treatment plan. |
| 3<br><br>Inaccurate; omitting adjuvant radiation is not recommended | False. The decision to recommend adjuvant radiation therapy in an 80-year-old woman with the described pathology after a modified radical mastectomy involves considering several factors. Here are some pertinent considerations:<br>1. **Age and General Health**: At 80 years old, the patient's overall health and life expectancy play significant roles in determining the aggressiveness of treatment. If she is in good health with a reasonable life expectancy, she might benefit from further adjuvant therapy.<br>2. **Tumor Characteristics**: The tumor is relatively small (1.3 cm), well-differentiated, and with no lymphovascular invasion, which are all favorable prognostic factors. However, a Ki-67 of 50% indicates a high proliferative rate, suggesting a more aggressive tumor biology.<br>3. **Hormone Receptor Status**: ER/PR positive status is generally favorable and indicates that the tumor may respond well to hormonal therapy, which should be considered as part of the adjuvant treatment.<br>4. **Lymph Node Status**: No lymph nodes involved out of ten examined is a good prognostic sign; however, it does not automatically rule out the benefit of radiation, especially considering other tumor characteristics and patient-specific factors.<br>5. **Radiation Therapy Considerations**: While the traditional factors might suggest a lower risk category where radiation might be omitted (particularly considering her age), the high Ki-67 could sway the decision towards offering radiation to mitigate the risk of locoregional recurrence, especially in light of the aggressive tumor biology indicated by this marker.<br><br>Thus, it might be appropriate to discuss the potential benefits and risks of adjuvant radiation therapy with the patient, taking into account her personal values, potential for improved local control, and the side effects associated with treatment. A nuanced approach involving a multidisciplinary team discussion would be most beneficial in making a tailored decision for her care. |

| | |
|---|---|
| 4<br><br>Inaccurate; omitting adjuvant radiation is not recommended | False.<br><br>In this case, the decision to recommend adjuvant radiation therapy, despite the successful surgical outcomes and the absence of lymph node involvement, could be influenced by several factors:<br>1. **Age and Comorbidities**: Although she is 80 years old, if her overall health status is good and she is expected to have a reasonable life expectancy, the benefit of adjuvant radiation in reducing local recurrence might outweigh the risks.<br>2. **High Ki-67 Index**: A Ki-67 index of 50% indicates a high proliferative rate of the tumor cells, which is associated with a higher risk of recurrence. This factor might tilt the decision towards recommending radiation despite the favorable other pathological findings.<br>3. **Tumor Characteristics and Guidelines**: Guidelines suggest considering radiation therapy in cases where there are higher risk features even if traditional nodal and margin status indicators are favorable. High Ki-67 is often considered a higher risk feature.<br>4. **Patient Preferences**: Depending on the patient's preferences and tolerance for potential side effects, the decision could vary.<br><br>Therefore, it is important to weigh all these factors, possibly discuss in a multidisciplinary team, and tailor the treatment recommendation to the individual patient's clinical context and preferences. |
| 5<br><br>Inaccurate; omitting adjuvant radiation is not recommended | False. While traditionally, some patients with early-stage breast cancer with no lymph node involvement might not receive adjuvant radiation therapy after mastectomy, several factors in this case suggest considering radiation:<br>1. **Age**: While older age might sometimes lessen the inclination for aggressive treatments, each case is assessed on its own merits based on overall health and life expectancy.<br>2. **Tumor Biology**: The Ki-67 index of 50% indicates a high proliferative rate, suggesting a more aggressive tumor behavior which might benefit from additional local control measures such as radiation.<br>3. **Surgical Outcome**: Although the surgical margins are clear and there is no lymphovascular invasion, the presence of a relatively high Ki-67 in the context of breast cancer often prompts a more cautious approach.<br><br>In this scenario, the decision to recommend adjuvant radiation would be influenced by a multidisciplinary discussion, taking into account the patient's overall health, preferences, life expectancy, and specific tumor characteristics. Radiation therapy could be considered to reduce the risk of locoregional recurrence, especially given the high Ki-67 index. |

| | |
|---|---|
| Chain of Reasoning Prompting (Respectively): Question Sequence Ends When Accurate Answer is Given | 80 year old woman with new diagnosis of invasive ductal carcinoma, ER/PR positive, HER2 negative status post modified radical mastectomy and axillary lymph node dissection, final pathology demonstrating 1.3cm carcinoma, well differentiated, no lymphovascular invasion, margins negative by >2mm, Ki-67 50%, and zero of ten lymph nodes positive for macrometastases. Presenting for adjuvant radiation recommendations:<br>1.Considering this woman's age, what would be your adjuvant radiation recommendation?<br>2.Why might you consider omitting adjuvant radiation for this specific woman? |

| | 3.Based on these considerations, would you still recommend adjuvant radiation for this individual? |
|---|---|
| 1<br><br>Before CoR: partially accurate<br><br>Prompt 1: partially accurate<br><br>Prompt 2: partially accurate<br><br>Prompt 3: accurate<br><br>Accurate; omitting adjuvant radiation is recommended at CoR Prompt 3 | For the 80-year-old woman you described with invasive ductal carcinoma (ER/PR positive, HER2 negative) who has undergone a modified radical mastectomy and axillary lymph node dissection, the recommendations for adjuvant radiation therapy should be carefully considered given her age and the characteristics of her tumor. Here's a breakdown of factors to consider:<br><br>1. **Tumor Characteristics and Surgery Outcomes:**<br>• **Size and Differentiation:** The tumor is relatively small at 1.3 cm and well-differentiated.<br>• **Margins:** Negative margins greater than 2 mm suggest good local control achieved surgically.<br>• **Lymph Node Status:** No lymph node involvement (0/10 nodes positive).<br>• **Ki-67 Index:** Elevated at 50%, indicating a higher proliferative rate, which can be a marker for more aggressive behavior.<br>2. **Adjuvant Radiation Therapy Considerations:**<br>• **Chest Wall Radiation:** Typically considered if there are high-risk features such as close/positive margins, large tumor size, lymph node involvement, or unfavorable biology. In this case, the only high-risk feature is the high Ki-67, which might tilt considerations slightly towards offering radiation, but it's not definitive.<br>• **Regional Lymph Node Radiation:** Given the absence of lymph node involvement and the effective surgical clearance, radiation to the regional lymph nodes is generally not indicated.<br>• **Patient Age and Comorbidities:** At 80 years old, the overall health status and life expectancy of the patient must be considered. The potential benefits of radiation must be weighed against the risks of radiation-induced side effects and the impact on her quality of life.<br>3. **Oncotype DX or Other Genomic Testing:**<br>• If not already performed, genomic testing like Oncotype DX might be helpful in this scenario, particularly because of the high Ki-67. This test could provide additional prognostic information and help in assessing the benefit of additional systemic therapy, potentially impacting the decision regarding adjuvant radiation.<br>4. **Endocrine Therapy:**<br>• Since the tumor is ER/PR positive, endocrine therapy will likely play a critical role in reducing the risk of recurrence. The decision regarding adjuvant radiation may be influenced by the planned endocrine treatment regimen.<br><br>Given these considerations, the decision to recommend adjuvant radiation should be personalized based on a detailed discussion with the patient regarding the potential benefits and risks, considering her overall health status, comorbidities, and personal preferences. If her health is good and she is motivated to pursue aggressive measures to minimize recurrence risk, discussing the option of radiation, perhaps with a focus on the chest wall given the high Ki-67, might be reasonable. Otherwise, a more conservative approach focusing on systemic therapy might be more appropriate.<br><br>1. |

| | |
|---|---|
| | Given the specific details of this patient's case and her age, a nuanced approach would be appropriate for considering adjuvant radiation therapy:<br>1. **Chest Wall Radiation:**<br>• The primary factor favoring consideration of radiation in this scenario is the Ki-67 index of 50%, which indicates a high proliferative rate. However, the other favorable factors—small tumor size, clear margins, and absence of lymph node involvement—tend to reduce the necessity for radiation.<br>• At 80 years old, the benefit of chest wall radiation should be carefully weighed against the potential for side effects, such as skin irritation, fatigue, and the rarer risks of cardiac or pulmonary complications, especially if there are existing comorbidities.<br>2. **Quality of Life Considerations:**<br>• The decision should also take into account the patient's quality of life, her functional status, and any other comorbid conditions that might affect her tolerance for radiation therapy.<br>• Discussing the patient's values and preferences regarding the potential side effects of radiation and its impact on her daily life is crucial.<br>3. **Alternative Strategies:**<br>• Strong consideration should be given to systemic therapies, particularly endocrine therapy, due to the hormone receptor-positive status of the tumor. This approach has a significant impact on reducing recurrence with a potentially more favorable side effect profile compared to radiation.<br>• Monitoring closely with regular follow-ups might be more suitable if opting out of radiation, considering the effective surgical management already achieved.<br>**Recommendation:**<br>• **Conservative Approach:** If the patient has significant comorbidities or values a treatment plan with fewer side effects, focusing on systemic therapy alone (endocrine therapy) and regular follow-ups might be preferable.<br>• **Aggressive Approach:** If the patient is in good health, understands the potential benefits and risks, and prefers a more aggressive approach to minimize recurrence risk, discussing the potential benefit of chest wall radiation could be considered, emphasizing that the decision leans more on the Ki-67 proliferation index rather than other high-risk features.<br>In either scenario, a personalized discussion is essential to align the treatment plan with the patient's health status, life expectancy, and personal preferences regarding care and quality of life.<br>2. |

Omitting adjuvant radiation in the case of this 80-year-old woman with invasive ductal carcinoma, who has undergone a modified radical mastectomy with favorable surgical outcomes, can be justified based on several considerations:

1. **Favorable Tumor Characteristics and Surgical Outcome:**
   - **Small Tumor Size:** The tumor is relatively small (1.3 cm), which typically correlates with a lower risk of local recurrence.
   - **Negative Margins:** The surgical margins were negative and greater than 2 mm, indicating effective local control achieved through surgery.
   - **Absence of Lymph Node Involvement:** No lymph nodes were found to be positive, suggesting a lower risk of systemic spread and local recurrence.
   - **Well-differentiated Histology:** This often correlates with less aggressive tumor behavior.
2. **Patient Age and Life Expectancy:**
   - **Age-Related Risks:** At 80 years old, the balance between the benefits and risks of adjuvant radiation shifts, particularly because the side effects and complications from radiation might significantly impact her quality of life and overall health.
   - **Comorbidities:** Elderly patients often have other health issues that could make the potential side effects of radiation (like skin toxicity, pneumonitis, or cardiac effects) more problematic and harder to manage.
3. **High Ki-67 Index Consideration:**
   - While the Ki-67 index is elevated, suggesting a higher proliferation rate, the other factors (small size, no node involvement, clear margins) mitigate this risk somewhat. Genomic testing such as Oncotype DX could further clarify the recurrence risk and influence the decision about adjuvant treatments, potentially obviating the need for radiation.
4. **Potential Side Effects and Impact on Quality of Life:**
   - **Immediate Side Effects:** These can include skin changes, fatigue, and discomfort, which can be significant in older patients.
   - **Long-term Risks:** More serious complications like rib fractures, cardiac damage, or secondary cancers, although rare, might be more concerning in someone of advanced age where the benefits might not outweigh these risks over her expected lifetime.
5. **Efficacy of Alternative Therapies:**
   - **Endocrine Therapy:** Given the hormone receptor-positive status, endocrine therapy could provide significant protection against recurrence with a potentially more favorable side effect profile, which is particularly advantageous in an elderly patient.

Based on these factors, the decision to omit adjuvant radiation could be a prudent choice aimed at maximizing her quality of life while still maintaining a robust defense against cancer recurrence through less invasive means.

3.

| | |
|---|---|
| | Given the overall clinical scenario and the factors laid out, I would lean towards not recommending adjuvant radiation for this 80-year-old woman. The key points influencing this recommendation include:<br>1. **Low-Risk Tumor Features**: The small size of the tumor, well-differentiated histology, absence of lymph node involvement, and negative surgical margins are all factors that generally predict a lower risk of local recurrence.<br>2. **High Ki-67 Index**: Although the Ki-67 index is elevated, suggesting a higher cell proliferation rate, the absence of other high-risk features somewhat mitigates this concern. The impact of this index could be better understood in the context of genomic profiling, if available, which might influence the decision regarding the necessity of radiation.<br>3. **Patient's Age and Comorbidities**: At the age of 80, the patient's overall health, potential comorbidities, and life expectancy are crucial considerations. The side effects and potential complications of radiation therapy might outweigh the benefits, especially when her life expectancy and quality of life are taken into account.<br>4. **Alternatives to Radiation**: The hormone receptor-positive nature of the tumor strongly supports the use of endocrine therapy as an effective adjuvant treatment. This approach could provide a significant benefit in reducing recurrence risk with a more tolerable side effect profile for an elderly patient.<br>5. **Quality of Life Considerations**: Minimizing treatment-related side effects is particularly important in elderly patients. The potential acute and chronic side effects of radiation could significantly impact her quality of life, which is a critical consideration given her age.<br><br>In conclusion, focusing on systemic therapy such as endocrine treatment, coupled with vigilant follow-up, would likely be a more appropriate management strategy for this patient, avoiding the risks associated with radiation therapy while effectively managing the risk of recurrence. This approach should be discussed thoroughly with the patient, considering her values, expectations, and the potential impact on her quality of life. |
| 2<br><br>Before CoR: accurate<br><br>Accurate; omitting adjuvant radiation is recommended before CoR prompts | For the 80-year-old woman with early-stage, well-differentiated invasive ductal carcinoma, ER/PR positive, HER2 negative, the decision to recommend adjuvant radiation therapy needs careful consideration of her overall health, life expectancy, and preferences. The key aspects of her pathology report are:<br>1. **Tumor Size and Differentiation**: The tumor is relatively small at 1.3 cm and well-differentiated, which generally suggests a more favorable prognosis.<br>2. **Lymph Node Status**: No lymph node involvement (0/10) is a very favorable factor and significantly reduces the risk of local and regional recurrence.<br>3. **Margins**: Negative surgical margins (>2mm) also suggest a lower risk of local recurrence.<br>4. **Ki-67 Index**: A Ki-67 index of 50% indicates a higher proliferative rate, which is associated with a more aggressive tumor behavior.<br><br>Given these factors, the major points for consideration include:<br>• **Age and Comorbidities**: At 80 years old, the patient's life expectancy and any comorbid conditions are crucial in decision-making. If she has a good life expectancy and minimal comorbidities, adjuvant therapies might improve her overall prognosis.<br>• **Radiation Therapy Risks vs. Benefits**: Radiation can decrease local recurrence, but in the context of negative nodes and margins, the absolute benefit may be small. For an elderly patient, it's essential to weigh the potential side effects of radiation, such as skin changes, fatigue, and rarely, more severe complications like rib fractures or cardiac issues. |

<table>
<tr>
<td></td>
<td>

#### Current Guidelines and Recommendations:

- **Post-Mastectomy Radiation Therapy (PMRT):** Typically recommended for patients with high-risk features such as larger tumor size (>5 cm), lymphovascular invasion, or positive lymph nodes. In this patient, the absence of these high-risk features may argue against the need for PMRT.
- **Endocrine Therapy:** Given the ER/PR positive status, endocrine therapy will likely be a key component of her adjuvant treatment. This therapy can significantly reduce the risk of recurrence and improve survival with a favorable side effect profile, particularly important in older patients.

#### Personalized Approach:

For this patient, a thorough discussion about the risks and benefits of radiation therapy is essential. Consideration of her values and preferences regarding quality of life and treatment side effects is important. If she has significant comorbidities or values quality of life over aggressive cancer treatment, it might be reasonable to omit adjuvant radiation, focusing instead on systemic endocrine therapy.

#### Conclusion:

The decision for adjuvant radiation in this scenario would typically lean towards omission unless there are additional patient or tumor characteristics that suggest a higher-than-average risk of local recurrence. The management plan should be personalized, and involve a multidisciplinary discussion along with the patient's input to ensure that her treatment aligns with her overall health goals and preferences.

</td>
</tr>
<tr>
<td>

3

Before CoR: accurate

Accurate; omitting adjuvant radiation is the only pursued suggestion before CoR prompts

</td>
<td>

For an 80-year-old woman with the described pathology following a modified radical mastectomy and axillary lymph node dissection for invasive ductal carcinoma that is ER/PR positive and HER2 negative, the decision to recommend adjuvant radiation therapy needs to be carefully balanced against the patient's age, overall health, life expectancy, and the specifics of the tumor characteristics and surgical outcomes.

Given the pathology report:

- **Tumor Size and Differentiation:** The tumor is relatively small (1.3 cm) and well-differentiated.
- **Margins:** The surgical margins are clear by >2mm, which is favorable.
- **Lymph Node Involvement:** There are no lymph nodes positive for macrometastases out of the ten examined, which significantly reduces the risk of regional recurrence.
- **Ki-67 Index:** A Ki-67 of 50% indicates a high proliferative rate, suggesting a more aggressive tumor biology despite other favorable factors.

#### Adjuvant Radiation Considerations:

1. **Age and Comorbidities:** At 80 years old, the patient's life expectancy and any comorbid conditions play a critical role in deciding whether to proceed with adjuvant radiation. The potential benefits must outweigh the risks and impacts on quality of life.
2. **Radiation Benefits:**
   - Radiation therapy could potentially reduce the risk of local recurrence, particularly considering the high Ki-67 index.
   - However, in the absence of lymphovascular invasion, extensive nodal involvement, and with clear margins, the absolute benefit might be limited.

</td>
</tr>
</table>

| | |
|---|---|
| | 3. **Treatment Guidelines**:<br>• According to guidelines such as those from the National Comprehensive Cancer Network (NCCN), the decision to recommend radiation in a patient of this age and stage might lean towards omission, especially if she is considered frail or has significant comorbidities.<br>4. **Patient Preference**:<br>• Discussion with the patient about the benefits versus risks, such as the potential side effects of radiation (e.g., skin changes, fatigue, rare but serious effects like rib fractures or cardiac issues), is crucial.<br>• Understanding her values and preferences regarding quality of life versus potential extension of disease-free survival is important.<br>**Recommendation:**<br>• **Potential Omission of Radiation**: Given the overall favorable pathology and the patient's age, the omission of radiation could be considered, especially if her performance status is not optimal.<br>• **Hormonal Therapy**: Since the tumor is ER/PR positive, adjuvant hormonal therapy with an aromatase inhibitor or tamoxifen, depending on her menopausal status and medical fitness, would likely be beneficial and less physically burdensome than radiation.<br>This patient's case should be discussed in a multidisciplinary team meeting, and her preferences should be heavily weighted in the final decision. Further genetic profiling could also be considered if there are ambiguities regarding the aggressiveness of the tumor that might impact treatment decisions. |
| 4<br><br>Before CoR prompting: partially accurate<br><br>Prompt 1: partially accurate<br><br>Prompt 2: accurate<br><br>Accurate; omitting adjuvant radiation is recommended after CoR Prompt 2 | For an 80-year-old woman with the described pathology, the decision for adjuvant radiation therapy should be guided by her overall health status, life expectancy, and the specifics of her tumor characteristics and surgical outcomes. Here's how these factors inform the decision:<br>1. **Tumor Characteristics and Surgical Outcome**:<br>• **Size and Differentiation**: The tumor is relatively small at 1.3 cm and well-differentiated.<br>• **Margins**: Negative by >2mm, which is favorable.<br>• **Lymph Node Involvement**: No lymph node involvement (0/10) decreases the likelihood of local recurrence.<br>• **Ki-67**: The high Ki-67 index of 50% suggests a higher proliferation rate, indicating a more aggressive tumor biology despite other favorable factors.<br>2. **Guideline Considerations**:<br>• Guidelines suggest that adjuvant radiation therapy is typically considered if there are factors that increase the risk of local recurrence, such as close/positive margins or lymph node involvement, neither of which is present in this case.<br>• However, the high Ki-67 may raise concerns about the aggressive nature of the tumor, warranting a discussion about the benefits and risks of radiation.<br>3. **Patient Age and Comorbidities**:<br>• At 80 years old, it's crucial to consider her functional status, comorbidities, and life expectancy. These factors influence the potential benefit she might derive from adjuvant radiation, balancing it against the treatment burden and potential side effects. |

4. **Radiation Therapy Considerations:**
   - If you opt for radiation, considering the patient's age and likely goal of minimizing treatment burden and maximizing quality of life, a hypofractionated schedule could be considered. Hypofractionated radiation therapy has been shown to be effective and may reduce the overall treatment time and potentially the side effects.
5. **Shared Decision-Making:**
   - It's essential to engage in a detailed discussion with the patient (and her caregivers, if applicable) regarding the potential benefits of radiation in reducing local recurrence risk, weighed against the risks and her personal preferences and values.

Given these points, the decision to proceed with or without adjuvant radiation therapy should be made collaboratively, considering all medical and personal factors. If her general health is good and she is motivated to pursue all measures to prevent recurrence, discussing the potential use of adjuvant radiation could be appropriate, with a clear discussion about what to expect and how it aligns with her goals of care.

1.

Given the patient's age of 80 years, and the characteristics of the tumor and surgical outcomes, my recommendation would lean towards discussing the option of omitting adjuvant radiation, unless there are specific factors that suggest a higher benefit from the treatment. Here's the rationale:

1. **Favorable Pathologic Features:**
   - The small size of the tumor (1.3 cm), well-differentiated histology, absence of lymph node involvement, and clear margins are all favorable factors that suggest a lower risk of local recurrence.
2. **High Ki-67 Index:**
   - The high Ki-67 index indicates a more aggressive tumor, which is a point of concern. However, in the context of no other high-risk features (like positive margins or lymph node involvement), it might not alone justify the use of adjuvant radiation, especially considering the potential side effects and the patient's advanced age.
3. **Age and Quality of Life Considerations:**
   - At 80 years, the primary goal is often to maximize quality of life and minimize treatment-related morbidity. Radiation therapy, while generally well-tolerated, can still pose acute and potentially long-term side effects which might significantly impact her quality of life.
   - Additionally, the life expectancy and any comorbid conditions should be taken into account to assess whether the potential extension in disease-free survival justifies the immediate side effects of radiation therapy.
4. **Treatment Burden:**
   - Radiation therapy, even in a hypofractionated schedule, requires regular visits to a radiation facility, which can be a significant burden for elderly patients, especially those with mobility issues or other health problems.
5. **Shared Decision-Making:**
   - Engaging in a detailed discussion with the patient about the potential benefits and drawbacks of radiation therapy is crucial. This should include a discussion about her values and preferences regarding treatment intensity and quality of life.

Based on these considerations, if the patient is generally healthy, understands the potential slight increase in recurrence risk without radiation, and prefers to avoid additional treatment, it would be reasonable to consider omitting radiation. However, if she is highly motivated to minimize the risk of recurrence and is willing to undergo the treatment despite the potential side effects, then discussing a tailored, possibly hypofractionated radiation plan would be appropriate.

2.

| | |
|---|---|
| | Omitting adjuvant radiation for this 80-year-old woman with early-stage, well-differentiated breast cancer, no lymph node involvement, and clear surgical margins can be considered for several reasons:<br><br>1. **Low Recurrence Risk:**<br>• The pathology report shows a small tumor size (1.3 cm), no lymphovascular invasion, and no lymph node involvement, all of which contribute to a lower risk of local recurrence. The negative margins further reduce the likelihood of residual disease.<br>2. **Patient Age and Life Expectancy:**<br>• At 80 years old, her life expectancy must be considered. The benefits of radiation in terms of prolonging survival might be limited when compared to a younger patient, given that the natural life expectancy might be shorter due to age and possibly other comorbidities.<br>3. **Quality of Life Considerations:**<br>• Radiation therapy can lead to side effects such as fatigue, skin irritation, and, less commonly, more severe effects such as rib fractures or cardiac issues, particularly pertinent in older patients who may have pre-existing conditions. The treatment regimen, even if hypofractionated, requires frequent visits to a radiation facility, which can be burdensome and may impact her overall quality of life.<br>4. **High Ki-67 Index:**<br>• While the Ki-67 index is high, suggesting a more aggressive tumor behavior, the absence of other high-risk features might still make the omission of radiation a reasonable option, especially when balancing the pros and cons of radiation in the context of her overall health status and prognosis.<br>5. **Current Guidelines and Studies:**<br>• Current guidelines and studies often suggest that in cases like hers, where traditional high-risk factors (like positive margins or nodal involvement) are absent, the added benefit of adjuvant radiation may be minimal. Studies such as the CALGB 9343 trial have shown that older women with early-stage, ER-positive breast cancer and no lymph node involvement have relatively low rates of local recurrence, and the absolute benefit of radiation in such cases is small.<br>6. **Patient Preferences:**<br>• Her preferences regarding treatment intensity, potential side effects, and how she values quality of life over marginal gains in survival or recurrence prevention should significantly influence the decision-making process.<br><br>In summary, considering her age, the favorable tumor characteristics, and the potential impacts on her quality of life, omitting radiation could be a reasonable choice that aligns with a goal of care focused on maintaining quality of life while managing the cancer effectively. |

| | |
|---|---|
| 5<br><br>Before CoR: inaccurate<br><br>Prompt 1: accurate<br><br>Accurate; omitting adjuvant radiation is recommended after CoR Prompt 1 | For an 80-year-old woman with an early-stage invasive ductal carcinoma that is ER/PR positive and HER2 negative, who has undergone a modified radical mastectomy and axillary lymph node dissection with favorable pathology findings, the decision for adjuvant radiation therapy would be nuanced. Here are several considerations:<br><br>1. **Age and Comorbidities:** Given her age, it's important to consider her overall health, life expectancy, and potential comorbidities which could impact her tolerance to additional treatment.<br>2. **Tumor Characteristics and Pathology:**<br>• **Size and Differentiation:** The tumor is relatively small (1.3 cm) and well-differentiated, which are favorable prognostic factors.<br>• **Lymph Node Status:** With no lymph nodes involved, her risk of regional recurrence is lower.<br>• **Margins:** Negative surgical margins (>2mm) further reduce the risk of local recurrence.<br>• **Ki-67 Index:** A Ki-67 of 50% indicates a higher proliferative index, which could suggest a more aggressive tumor biology despite other favorable factors.<br>3. **Radiation Therapy Considerations:**<br>• **Chest Wall Radiation:** In older patients with no lymph node involvement and clear margins, the benefit of chest wall radiation is less clear and could be considered overtreatment, especially given the potential side effects in the context of limited life expectancy.<br>• **Regional Nodal Radiation:** Since the axillary nodes were negative, regional nodal irradiation would typically not be indicated.<br>4. **Guidelines and Predictive Tools:**<br>• **Clinical Guidelines:** Most guidelines might suggest that adjuvant radiotherapy can be omitted in older patients with favorable risk profiles to avoid unnecessary side effects.<br>• **Decision Aids:** Tools like the Oncotype DX might be used in some scenarios to further stratify risk and guide the use of adjuvant therapies, although its primary utility is in guiding the use of systemic therapy rather than radiation.<br>5. **Patient Preferences:** Discussing the potential benefits and drawbacks of adjuvant radiation, including the impact on quality of life, is crucial. Many elderly patients prioritize quality of life over small potential increases in disease control.<br><br>Given these factors, it may be reasonable to consider omitting adjuvant radiation in this patient, focusing instead on hormonal therapy given her hormone receptor-positive status. This approach would aim to minimize treatment-related side effects while managing her cancer effectively. Always, the final decision should be made after a detailed discussion with the patient about her values and preferences.<br><br>1. |

| | |
|---|---|
| | Given the patient's age, tumor characteristics, and post-surgical pathology findings, my recommendation would lean towards omitting adjuvant radiation therapy. Here's the rationale:<br><br>1. **Tumor and Surgical Factors**: The patient has a small, well-differentiated tumor with no lymph node involvement and clear margins. These factors all suggest a lower risk of local and regional recurrence.<br>2. **Age and Quality of Life**: At 80 years old, the primary goal should be to maximize her quality of life while managing her breast cancer effectively. Radiation therapy, particularly in the chest wall and axillary regions, carries risks of side effects such as skin irritation, fatigue, and potentially more severe complications like pneumonitis or rib fractures, which could significantly impact her quality of life.<br>3. **Lack of Clear Benefit**: The absence of lymph node involvement and the presence of negative margins reduce the potential benefit of radiation therapy in significantly improving her survival or decreasing recurrence rates, given the already low risk based on her pathology.<br>4. **Guideline Support**: Clinical guidelines often support the omission of adjuvant radiation in older patients with early-stage, low-risk breast cancer who have undergone adequate surgery, especially when life expectancy and the balance between treatment benefits and harms are considered.<br>5. **Hormonal Therapy**: Since her tumor is ER/PR positive, hormonal therapy would be an appropriate and less physically burdensome approach to reduce the risk of recurrence.<br><br>In conclusion, omitting adjuvant radiation and focusing on hormonal therapy would likely provide an optimal balance of effective cancer control and maintaining a good quality of life for this patient. This decision should be discussed in detail with the patient, taking into account her personal values, preferences, and any other health considerations. |

First Example (ChatGPT 4o)

1. 40 year old woman with new diagnosis of invasive ductal carcinoma, ER/PR positive, HER2 negative status post lumpectomy and sentinel lymph node dissection, final pathology demonstrating 1.3cm carcinoma, well differentiated, no lymphovascular invasion, margins negative by >2mm, Ki-67 50%, and all lymph nodes negative, presenting for adjuvant radiation recommendations: **Expected response: Whole breast with boost**

### General Observations

-In contrast with the older model (ChatGPT 4), ChatGPT 4o further details its consideration of the patient's age, not just the Ki-67 index, when suggesting a boost to the tumor bed
-Unlike ChatGPT 4, this model does not list accelerated partial breast irradiation as an

adjuvant radiation recommendation
-5/5 accurate

<table>
<tr>
<td>1<br><br>Accurate; whole breast irradiation with boost is suggested</td>
<td>
For a 40-year-old woman with a new diagnosis of invasive ductal carcinoma, ER/PR positive, HER2 negative, who has undergone a lumpectomy and sentinel lymph node dissection, the final pathology shows:
<ul>
<li>Tumor size: 1.3 cm</li>
<li>Grade: Well differentiated</li>
<li>Lymphovascular invasion: None</li>
<li>Margins: Negative by >2mm</li>
<li>Ki-67: 50%</li>
<li>Lymph nodes: All negative</li>
</ul>
The patient is presenting for adjuvant radiation recommendations.<br><br>
Given these details, the standard adjuvant radiation therapy would typically include whole breast irradiation. Here are some key points to consider for her adjuvant radiation therapy:<br><br>
Radiation Therapy Recommendations
<ol>
<li>Whole Breast Irradiation (WBI):
<ul>
<li>Dose: Typically, 45-50 Gy in 25 fractions or a hypofractionated regimen of 40-42.5 Gy in 15-16 fractions.</li>
<li>Boost to Tumor Bed: Consider a boost of 10-16 Gy in 4-8 fractions to the tumor bed, especially given her young age and the Ki-67 index of 50%, which indicates a higher proliferation rate.</li>
</ul></li>
<li>Hypofractionated Radiation Therapy:
<ul>
<li>Hypofractionated regimens have become more common and are equally effective, with fewer side effects and more convenience for patients. This would involve 40-42.5 Gy in 15-16 fractions, followed by a tumor bed boost.</li>
</ul></li>
</ol>
</td>
</tr>
</table>

| | |
|---|---|
| | 3. **Consideration of Oncoplastic Surgery:**<br>• If the patient had an oncoplastic lumpectomy, ensure that the surgical bed is accurately marked for the boost.<br>**Factors Supporting the Use of Boost**<br>• **Age:** Younger patients (less than 50 years) have a higher risk of local recurrence, justifying the use of a boost.<br>• **Tumor Characteristics:** High Ki-67 (50%) indicates higher proliferation, which may be associated with increased risk of recurrence.<br>• **Margin Status:** Although margins are negative by >2mm, the boost provides additional assurance of local control.<br>**Discussion with the Patient**<br>• Discuss the potential benefits and risks of radiation therapy, including acute side effects like skin changes and fatigue, and long-term risks such as fibrosis and secondary malignancies.<br>• Consider the patient's preferences, lifestyle, and ability to attend daily treatments for several weeks.<br>**Follow-Up and Monitoring**<br>• Regular follow-up visits for physical examinations and imaging as per standard post-treatment guidelines.<br>• Monitoring for potential late effects of radiation and managing any complications that arise.<br>**Multidisciplinary Discussion**<br>• Ensure coordination with the patient's medical oncologist for potential adjuvant systemic therapy, considering her ER/PR positive status, which may include hormonal therapy.<br>By providing these comprehensive recommendations, the patient can be well-informed and involved in the decision-making process regarding her adjuvant radiation therapy. |

| | |
|---|---|
| 2<br><br>Accurate; whole breast irradiation with boost is suggested | For a 40-year-old woman with invasive ductal carcinoma, ER/PR positive, HER2 negative, who has undergone lumpectomy and sentinel lymph node dissection with the final pathology showing the following:<br><br>• Tumor size: 1.3 cm<br>• Grade: Well-differentiated<br>• Lymphovascular invasion: None<br>• Margins: Negative by >2 mm<br>• Ki-67: 50%<br>• Lymph nodes: All negative<br><br>The standard adjuvant radiation recommendations for such a patient would generally include:<br><br>1. **Whole Breast Irradiation (WBI):** This is the typical approach following breast-conserving surgery (lumpectomy) to reduce the risk of local recurrence. The common dose and fractionation schedules include:<br>• Conventional fractionation: 50 Gy in 25 fractions (2 Gy per fraction) over 5 weeks.<br>• Hypofractionation: 40-42.5 Gy in 15-16 fractions (2.66-2.67 Gy per fraction) over 3-4 weeks, which is now often preferred due to similar efficacy and lower toxicity.<br>2. **Boost to the Tumor Bed:** Given the patient's age and the Ki-67 of 50%, a tumor bed boost could be considered to further reduce the risk of local recurrence. The boost dose is typically:<br>• 10-16 Gy in 4-8 fractions (2-2.5 Gy per fraction) following the completion of WBI.<br>3. **Regional Nodal Irradiation (RNI):** Given that all lymph nodes are negative and there is no lymphovascular invasion, RNI is not typically indicated in this scenario. However, the Ki-67 of 50% is relatively high and might influence the discussion depending on other individual patient factors and multidisciplinary team input.<br><br>**Considerations:**<br><br>• **Age and Aggressiveness:** The patient's young age and the high Ki-67 index (50%) suggest a more aggressive tumor biology, which can influence the decision for the inclusion of a boost and the consideration of systemic therapies if not already planned.<br>• **Systemic Therapy:** Ensure the patient is also considered for appropriate systemic therapies (e.g., hormonal therapy) based on ER/PR positivity and overall risk profile.<br><br>**Discussion with the Patient:**<br><br>• Explain the rationale for radiation therapy, including the benefits of reducing local recurrence.<br>• Discuss potential side effects, such as skin changes, fatigue, and rare risks of more serious complications.<br>• Involve the patient in shared decision-making regarding the treatment schedule, especially if choosing between conventional and hypofractionated schedules.<br>• Consider patient preferences, lifestyle, and any logistical issues related to treatment duration and frequency.<br><br>Overall, the recommended approach is whole breast irradiation with consideration for a tumor bed boost, factoring in the patient's specific pathology and clinical context. |

<table>
<tr>
<td>3<br><br>Accurate; whole breast irradiation with boost is suggested</td>
<td>

For a 40-year-old woman with a new diagnosis of invasive ductal carcinoma, ER/PR positive, HER2 negative, who has undergone lumpectomy and sentinel lymph node dissection with final pathology showing a 1.3 cm well-differentiated tumor, no lymphovascular invasion, negative margins (>2mm), Ki-67 50%, and all lymph nodes negative, the following adjuvant radiation recommendations can be considered:

#### Adjuvant Radiation Recommendations:

1. **Whole Breast Radiation Therapy (WBRT):**
    - **Dose:** Standard WBRT involves delivering 50 Gy in 25 fractions over 5 weeks or a hypofractionated schedule of 40 Gy in 15 fractions over 3 weeks.
    - **Boost to Tumor Bed:** Consider a boost to the tumor bed given her age and the Ki-67 index. A typical boost dose is 10-16 Gy in 4-8 fractions.
    - **Technique:** 3D conformal radiotherapy (3D-CRT) or intensity-modulated radiotherapy (IMRT) to ensure adequate coverage of the breast while minimizing dose to the heart and lungs.
2. **Partial Breast Irradiation (PBI):**
    - **Indication:** PBI can be considered for patients with favorable features such as her small, well-differentiated tumor and negative lymph nodes. However, given her young age, whole breast radiation is generally preferred.
    - **Techniques:** PBI can be delivered via external beam radiation, brachytherapy, or intraoperative radiation therapy (IORT).

#### Considerations:

- **Age and Ki-67 Index:** Her young age and high Ki-67 index suggest a more aggressive tumor biology, supporting the use of adjuvant radiation to reduce the risk of local recurrence.
- **Cosmetic Outcomes:** Discuss with the patient the potential cosmetic outcomes of radiation therapy, especially if a boost is delivered to the tumor bed.
- **Follow-Up:** Regular follow-up with clinical exams and imaging as appropriate to monitor for recurrence and manage any long-term effects of radiation.

#### Multidisciplinary Discussion:

- Engage in a multidisciplinary tumor board discussion involving surgical oncology, medical oncology, and radiation oncology to ensure a comprehensive treatment plan.
- Consider genetic counseling and testing if not already performed, given her young age and the implications for systemic therapy.

#### Patient Education:

- Educate the patient about the potential side effects of radiation therapy, including skin changes, fatigue, and rare long-term risks such as cardiotoxicity or secondary malignancies.
- Provide support resources such as psychosocial support, nutritional counseling, and survivorship care planning.

This plan should be tailored to the patient's preferences and overall health status, and it should be based on a thorough discussion of the risks and benefits of adjuvant radiation therapy.

</td>
</tr>
</table>

4

Accurate; whole breast irradiation with boost is suggested

For a 40-year-old woman with a new diagnosis of invasive ductal carcinoma (IDC), ER/PR positive, HER2 negative, status post-lumpectomy and sentinel lymph node dissection, the adjuvant radiation therapy recommendations would typically be as follows:

#### Adjuvant Radiation Therapy Recommendations:

1. **Whole Breast Irradiation (WBI):**
   - **Dose and Fractionation:**
     - Standard fractionation: 45-50 Gy in 25-28 fractions over 5-6 weeks.
     - Hypofractionation: 40-42.5 Gy in 15-16 fractions over 3-4 weeks (preferred for many patients, including those with smaller tumors and no lymph node involvement).
2. **Boost to the Tumor Bed:**
   - Given the patient's age and high Ki-67, a boost to the tumor bed might be considered to reduce the risk of local recurrence.
   - **Dose:** Typically 10-16 Gy in 4-8 fractions.
3. **Techniques and Considerations:**
   - **3D Conformal Radiotherapy (3D-CRT):** Commonly used to deliver WBI.
   - **Intensity-Modulated Radiotherapy (IMRT):** May be considered to improve dose homogeneity and reduce dose to surrounding normal tissues.
   - **Deep Inspiration Breath Hold (DIBH):** Particularly if the tumor is on the left side, to minimize radiation exposure to the heart.
   - **Image-Guided Radiotherapy (IGRT):** For accurate daily positioning.
4. **Regional Nodal Irradiation (RNI):**
   - Not indicated since the sentinel lymph nodes are negative.
5. **Considerations for Young Age and High Proliferation Index (Ki-67 50%):**
   - Close follow-up due to the higher risk of recurrence associated with younger age and higher proliferation index.

#### Additional Considerations:

- **Multidisciplinary Discussion:**
  - Ensure a coordinated approach involving the medical oncologist, surgeon, and radiation oncologist.
  - Discuss potential need for adjuvant systemic therapy (e.g., endocrine therapy) considering the patient's hormone receptor status.
- **Patient Counseling:**
  - Discuss the benefits and potential side effects of radiation therapy.
  - Address fertility preservation if applicable, considering the patient's age.

These recommendations are based on current guidelines and the specifics of the patient's pathology. Individual patient factors and preferences should always be considered in finalizing the treatment plan.

| | |
|---|---|
| 5<br><br>Accurate; whole breast irradiation with boost is suggested | For a 40-year-old woman with invasive ductal carcinoma, ER/PR positive, HER2 negative, who has undergone a lumpectomy and sentinel lymph node dissection, the final pathology showing a 1.3 cm well-differentiated carcinoma with no lymphovascular invasion, negative margins (>2 mm), Ki-67 50%, and negative lymph nodes, the following adjuvant radiation recommendations are generally considered:<br><br>1. **Whole Breast Radiation Therapy (WBRT):**<br>• **Standard Fractionation:** 45-50.4 Gy in 25-28 fractions.<br>• **Hypofractionation:** 40-42.5 Gy in 15-16 fractions, which is increasingly being used due to its convenience and comparable efficacy and safety profile.<br>2. **Boost Radiation:**<br>• Given her young age (40 years) and the high Ki-67 (50%), a tumor bed boost is often recommended.<br>• Typical boost dose: 10-16 Gy in 4-8 fractions.<br>3. **Regional Nodal Irradiation (RNI):**<br>• In this case, given the absence of lymph node involvement and other high-risk features, RNI is generally not indicated.<br>4. **Considerations:**<br>• The decision for adjuvant radiation therapy should be discussed in a multidisciplinary setting, considering the patient's overall health, preferences, and any potential contraindications.<br>• Genetic counseling might be recommended due to her age at diagnosis to assess for hereditary breast cancer syndromes, which could impact her long-term surveillance and management.<br>5. **Hormonal Therapy:**<br>• As the patient is ER/PR positive, she should be counseled regarding adjuvant endocrine therapy, typically with tamoxifen or an aromatase inhibitor, depending on her menopausal status and other factors.<br><br>These recommendations aim to minimize the risk of local recurrence while balancing the potential side effects of treatment. It's essential to tailor the approach to the individual patient's clinical scenario and personal preferences. |

Second Example (ChatGPT 4o)

2. 40 year old woman with new diagnosis of invasive ductal carcinoma, ER/PR positive, HER2 negative status post modified radical mastectomy and axillary lymph node dissection, final pathology demonstrating 1.3cm carcinoma, well differentiated, no lymphovascular invasion, margins negative by >2mm, Ki-67 50%, and five of ten lymph nodes positive for macrometastases. Presenting for adjuvant radiation recommendations:
**Expected response: Regional Nodal Irradiation**

### General Observations

-5/5 accurate

| | |
|---|---|
| 1<br><br>Accurate; regional nodal irradiation is suggested | For the 40-year-old woman with invasive ductal carcinoma (IDC), ER/PR positive, HER2 negative, who has undergone a modified radical mastectomy and axillary lymph node dissection, the final pathology indicating a 1.3 cm carcinoma, well-differentiated, no lymphovascular invasion, margins negative by >2mm, Ki-67 at 50%, and five of ten lymph nodes positive for macrometastases, the adjuvant radiation therapy recommendations would typically include:<br><br>**Rationale for Adjuvant Radiation Therapy**<br><br>Given the presence of five positive lymph nodes, adjuvant radiation therapy is recommended to reduce the risk of local recurrence and potentially improve overall survival. The significant number of positive lymph nodes suggests a higher risk of locoregional recurrence, which supports the benefit of post-mastectomy radiation therapy (PMRT).<br><br>**Recommended Radiation Therapy Approach**<br><br>1. **Chest Wall Irradiation:**<br>• **Dose:** Typically 50 Gy in 25 fractions or an equivalent fractionation schedule.<br>• **Rationale:** To target any residual microscopic disease in the chest wall area.<br>2. **Regional Nodal Irradiation (RNI):**<br>• **Target Areas:** Supraclavicular, infraclavicular, axillary, and internal mammary lymph nodes.<br>• **Dose:** Typically 50 Gy in 25 fractions or an equivalent fractionation schedule.<br>• **Rationale:** Given the five positive lymph nodes, irradiating the regional nodal basins can help address any microscopic residual disease in these areas and further reduce the risk of recurrence.<br><br>**Specific Considerations**<br><br>• **Treatment Planning:** Utilize CT-based planning to ensure precise targeting of the chest wall and nodal regions while minimizing exposure to adjacent normal tissues, such as the heart and lungs.<br>• **Techniques:** Advanced techniques like intensity-modulated radiation therapy (IMRT) or volumetric modulated arc therapy (VMAT) may be considered to optimize dose distribution and minimize toxicity.<br>• **Patient Positioning:** Consider the use of prone positioning or deep inspiration breath-hold (DIBH) techniques to reduce heart and lung dose, especially in left-sided breast cancers. |

| | |
|---|---|
| | ### Multidisciplinary Discussion<br>• **Systemic Therapy:** Coordinate with the medical oncologist to integrate radiation therapy with ongoing or planned systemic treatments, such as chemotherapy, hormonal therapy, or targeted therapies.<br>• **Follow-up and Monitoring:** Establish a schedule for follow-up visits to monitor for acute and late toxicities, assess response to treatment, and detect any signs of recurrence early.<br>### Conclusion<br>Adjuvant radiation therapy encompassing the chest wall and regional lymph nodes is recommended for this patient to reduce the risk of local and regional recurrence. The treatment plan should be individualized based on the patient's anatomy, overall health, and preferences, while integrating multidisciplinary input to optimize outcomes. |

| | |
|---|---|
| 2<br><br>Accurate; regional nodal irradiation is suggested | Based on the provided clinical information for the 40-year-old woman with invasive ductal carcinoma, ER/PR positive, HER2 negative, status post modified radical mastectomy, and axillary lymph node dissection, the final pathology reveals a 1.3 cm carcinoma, well-differentiated, no lymphovascular invasion, negative margins by >2mm, a Ki-67 of 50%, and five out of ten lymph nodes positive for macrometastases. Here are the adjuvant radiation therapy recommendations:<br><br>**Adjuvant Radiation Therapy Recommendations:**<br><br>1. Chest Wall Radiation:<br>• Given the presence of positive lymph nodes (5/10), adjuvant radiation to the chest wall is recommended.<br>• This is to reduce the risk of local recurrence in the chest wall and surrounding tissues.<br><br>2. Regional Nodal Irradiation (RNI):<br>• Since there are multiple positive lymph nodes, regional nodal irradiation (RNI) should be considered.<br>• Target areas typically include the supraclavicular lymph nodes, the internal mammary nodes, and the remaining axillary nodes.<br>• This is crucial for controlling any microscopic disease in these regions and improving overall survival.<br><br>3. Radiation Dose and Fractionation:<br>• Typical radiation dose to the chest wall is around 50 Gy in 25 fractions or 45-50 Gy in 25 fractions, depending on the treatment center's protocol.<br>• An additional boost to the mastectomy scar or areas of close margins might be considered, typically around 10-16 Gy in 4-8 fractions.<br>• For regional nodal irradiation, a similar dose of 50 Gy in 25 fractions is generally used.<br><br>4. Timing of Radiation:<br>• Adjuvant radiation therapy should ideally start within 4-6 weeks post-surgery, once healing has occurred and chemotherapy, if indicated, is completed.<br><br>5. Multidisciplinary Approach:<br>• A multidisciplinary discussion involving the surgical oncologist, medical oncologist, and radiation oncologist is essential to tailor the treatment plan.<br>• Integration of systemic therapy (e.g., adjuvant chemotherapy and/or endocrine therapy) should be coordinated with radiation therapy to optimize patient outcomes.<br><br>6. Patient Counseling and Follow-up:<br>• Discuss potential side effects of radiation therapy, including skin changes, fatigue, lymphedema, and potential long-term risks.<br>• Provide detailed instructions on skin care during radiation therapy.<br>• Plan for regular follow-up to monitor for treatment response and manage any side effects.<br><br>**Additional Considerations:**<br>• Oncotype DX Testing: If not already done, genomic testing like Oncotype DX may help guide the decision for adjuvant chemotherapy.<br>• Endocrine Therapy: Given the ER/PR positive status, adjuvant endocrine therapy (e.g., tamoxifen or aromatase inhibitors) should be considered as part of the overall treatment plan.<br><br>It is essential to personalize the radiation therapy plan based on the patient's specific clinical scenario and preferences, involving her in the decision-making process. |

| | |
|---|---|
| 3<br><br>Accurate; regional nodal irradiation is suggested | For this 40-year-old woman with invasive ductal carcinoma, ER/PR positive, HER2 negative, post-modified radical mastectomy and axillary lymph node dissection, with the following pathology characteristics: 1.3 cm well-differentiated carcinoma, no lymphovascular invasion, negative margins by >2 mm, Ki-67 of 50%, and five of ten lymph nodes positive for macrometastases, the following adjuvant radiation recommendations are considered:<br><br>1. **Indications for Radiation Therapy:**<br>• **Positive Lymph Nodes:** The presence of macrometastases in five out of ten lymph nodes indicates a higher risk of locoregional recurrence, making adjuvant radiation therapy necessary.<br>• **Tumor Size and Other Factors:** While the primary tumor size is relatively small (1.3 cm), the high Ki-67 index (50%) suggests a more aggressive tumor biology, further supporting the need for adjuvant radiation.<br><br>2. **Radiation Therapy Recommendations:**<br>• **Chest Wall Radiation:** Given the modified radical mastectomy, radiation to the chest wall is recommended to reduce the risk of local recurrence.<br>• **Regional Nodal Irradiation (RNI):** Due to the positive lymph nodes, radiation to the regional lymph nodes (including the supraclavicular, infraclavicular, and internal mammary nodes) is recommended to address potential microscopic disease.<br><br>3. **Radiation Dose and Fractionation:**<br>• **Chest Wall:** A typical regimen would be 50 Gy in 25 fractions over 5 weeks, followed by a boost to the mastectomy scar area if indicated, based on surgical findings and margin status.<br>• **Regional Nodes:** Similar dosing of 50 Gy in 25 fractions over 5 weeks can be applied to the regional nodes.<br><br>4. **Potential Considerations:**<br>• **Patient Age and Risk Factors:** Given the patient's younger age and the aggressive nature suggested by the Ki-67 index, comprehensive treatment to the chest wall and regional nodes is crucial.<br>• **Concurrent Systemic Therapy:** Coordination with medical oncology to integrate radiation with any ongoing or planned systemic therapies, such as hormone therapy or chemotherapy, is essential.<br><br>5. **Follow-Up and Monitoring:**<br>• Regular follow-up to monitor for radiation-related side effects and assess for any signs of recurrence.<br>• Multidisciplinary approach involving medical oncology, surgical oncology, and radiation oncology to ensure comprehensive care.<br><br>In summary, adjuvant radiation therapy to both the chest wall and regional lymph nodes is recommended for this patient, considering the significant nodal involvement and high Ki-67 index. This approach aims to reduce the risk of locoregional recurrence and improve overall survival outcomes. |

<table>
<tr>
<td>4<br><br>Accurate; regional nodal irradiation is suggested</td>
<td>
For a 40-year-old woman with invasive ductal carcinoma (IDC), ER/PR positive, HER2 negative, who has undergone a modified radical mastectomy and axillary lymph node dissection, the pathology findings suggest a relatively small primary tumor (1.3 cm), well-differentiated, without lymphovascular invasion, but with significant nodal involvement (5 out of 10 nodes positive for macrometastases). Given these findings, adjuvant radiation therapy is indicated to reduce the risk of local recurrence and improve overall survival. Here are the recommendations for adjuvant radiation therapy:<br><br>
Adjuvant Radiation Recommendations:<br><br>
1. Chest Wall Radiation:<br>
• Indication: Given the positive nodal involvement and relatively high Ki-67 index (50%), chest wall radiation is recommended.<br>
• Dose: Typically, 50 Gy in 25 fractions over 5 weeks, followed by a boost to the mastectomy scar if indicated based on tumor location and margin status.<br><br>
2. Regional Nodal Irradiation (RNI):<br>
• Indication: Due to the presence of positive axillary lymph nodes, regional nodal irradiation is recommended to include the supraclavicular, infraclavicular, and internal mammary nodes.<br>
• Dose: Generally, 50 Gy in 25 fractions over 5 weeks, which can be delivered concurrently with chest wall radiation.<br><br>
Factors to Consider:<br>
• Age and Risk of Recurrence: Younger patients have a higher risk of recurrence, justifying a more aggressive adjuvant treatment approach.<br>
• Tumor Characteristics: The absence of lymphovascular invasion and well-differentiated status are favorable, but the significant nodal involvement increases the recurrence risk.<br>
• Systemic Therapy: Ensure coordination with medical oncology to optimize systemic therapy, including hormone therapy and possibly chemotherapy if not already initiated.<br><br>
Potential Side Effects:<br>
• Acute: Skin reactions (erythema, desquamation), fatigue, esophagitis (if internal mammary nodes are irradiated).<br>
• Chronic: Risk of lymphedema, rib fracture, pneumonitis, brachial plexopathy, and cardiac toxicity (particularly if left-sided breast cancer).<br><br>
Follow-Up:<br>
• Close follow-up is essential to monitor for potential side effects, response to therapy, and signs of recurrence. Regular imaging and physical examinations will be part of the follow-up plan.<br><br>
Multidisciplinary Discussion:<br>
• Discuss this plan in a multidisciplinary tumor board meeting to ensure comprehensive care and consideration of all treatment aspects.<br><br>
Implementing these recommendations should provide optimal local control and contribute to the patient's overall treatment strategy.
</td>
</tr>
</table>

| | |
|---|---|
| 5<br><br>Accurate; regional nodal irradiation is suggested | For a 40-year-old woman with invasive ductal carcinoma, ER/PR positive, HER2 negative, status post modified radical mastectomy and axillary lymph node dissection, with the given pathological findings, adjuvant radiation therapy is an important consideration. Here's a detailed recommendation plan:<br><br>**Indications for Adjuvant Radiation Therapy:**<br>1. **Positive Lymph Nodes:** Presence of macrometastases in 5 out of 10 lymph nodes.<br>2. **Tumor Characteristics:** 1.3 cm well-differentiated tumor, although the size is relatively small, the positive lymph nodes necessitate further treatment.<br>3. **High Proliferative Index:** Ki-67 is 50%, indicating a high proliferation rate.<br><br>**Goals of Adjuvant Radiation Therapy:**<br>• Reduce the risk of local and regional recurrence.<br>• Improve overall survival.<br><br>**Recommended Radiation Therapy Plan:**<br>1. **Chest Wall Irradiation:**<br>• **Dose:** Typically 50 Gy in 25 fractions over 5 weeks.<br>• **Technique:** 3D conformal radiation therapy (3D-CRT) or intensity-modulated radiation therapy (IMRT) to spare surrounding tissues.<br>2. **Regional Nodal Irradiation:**<br>• **Inclusion of Supraclavicular and Axillary Nodes:** Due to the involvement of multiple lymph nodes.<br>• **Dose:** 50 Gy in 25 fractions.<br>• **Technique:** 3D-CRT or IMRT.<br>3. **Boost to Tumor Bed:**<br>• Generally not required as margins are negative by >2mm, but clinical judgment can be used based on additional risk factors and patient specifics.<br><br>**Considerations and Special Instructions:**<br>• **Patient Positioning:** Supine position with arms above head or a prone position if the patient's anatomy requires it.<br>• **Heart and Lung Sparing Techniques:** Especially important if the tumor is left-sided to minimize the dose to the heart and lungs. Techniques such as deep inspiration breath hold (DIBH) can be utilized.<br>• **Skin Care Management:** Educate the patient on skin care during radiation therapy.<br>• **Follow-Up and Monitoring:** Regular follow-up visits to monitor for acute and late toxicities, as well as to assess for any signs of recurrence.<br><br>**Multidisciplinary Approach:**<br>• **Coordination with Medical Oncology:** Ensure the patient continues on appropriate systemic therapies, such as endocrine therapy given her ER/PR positive status.<br>• **Patient Counseling:** Discuss the benefits, potential side effects, and the importance of adherence to the radiation schedule.<br><br>This comprehensive approach aims to maximize local control and minimize toxicity, contributing to improved overall outcomes for the patient. |

Third Example (ChatGPT 4o)

3. 80 year old woman with new diagnosis of invasive ductal carcinoma, ER/PR positive, HER2 negative status post modified radical mastectomy and axillary lymph node dissection, final pathology demonstrating 1.3cm carcinoma, well differentiated, no lymphovascular invasion, margins negative by >2mm, Ki-67 50%, and zero of ten lymph nodes positive for macrometastases. Presenting for adjuvant radiation recommendations:
**Expected response: No radiation**

### General Observations

Original Prompt:
-In comparison with ChatGPT-4, the model is slightly more direct when suggesting no adjuvant radiation therapy
-Again, the model erroneously implies or states that adjuvant radiation may be a consideration for the patient
Original Prompting:
-0/5 accurate

-Accuracy improved notably after alternative prompting

Phrasing 1:

Phrasing 2:

Phrasing 3:

Chain of Reasoning Prompting:

<table>
<tr>
<td>1<br><br>Inaccurate; omitting adjuvant radiation is not suggested as the only viable option</td>
<td>
For an 80-year-old woman with a new diagnosis of invasive ductal carcinoma, ER/PR positive, HER2 negative, status post modified radical mastectomy and axillary lymph node dissection, and given the final pathology:<br>
<ul>
<li>1.3 cm well-differentiated carcinoma</li>
<li>No lymphovascular invasion</li>
<li>Margins negative by >2 mm</li>
<li>Ki-67 50%</li>
<li>Zero of ten lymph nodes positive for macrometastases</li>
</ul>
Here are the considerations for adjuvant radiation therapy recommendations:<br>
<h3>Clinical Considerations:</h3>
<ol>
<li>Patient Age and Comorbidities:
<ul><li>Given her age (80 years), it is essential to consider her overall health, life expectancy, and any comorbidities that might affect her ability to tolerate radiation therapy.</li></ul></li>
<li>Tumor Characteristics:
<ul><li>The tumor is small (1.3 cm) and well-differentiated with no lymphovascular invasion and clear margins, indicating a favorable prognosis.</li>
<li>A high Ki-67 (50%) suggests a higher proliferative rate, which might indicate a more aggressive tumor biology.</li></ul></li>
<li>Node-Negative Status:
<ul><li>No lymph node involvement (zero of ten nodes positive) generally lowers the risk of recurrence.</li></ul></li>
</ol>
<h3>Adjuvant Radiation Therapy Options:</h3>
<ol>
<li>Omission of Radiation Therapy:
<ul><li>Given the patient's age, comorbidities, and favorable tumor characteristics, one option is to omit radiation therapy, especially if the patient has a limited life expectancy or significant comorbidities.</li>
<li>Some studies suggest that for older women with small, node-negative, hormone receptor-positive tumors, the benefit of radiation therapy might be minimal and can be safely omitted in selected cases.</li></ul></li>
</ol>
</td>
</tr>
</table>

2. **Partial Breast Irradiation (PBI):**
    - If radiation therapy is considered, PBI can be an option. It targets only the tumor bed and surrounding tissue, minimizing exposure to the rest of the breast and reducing treatment time.
    - Techniques for PBI include brachytherapy, intraoperative radiotherapy, or external beam radiotherapy.
3. **Whole Breast Irradiation (WBI):**
    - Although typically recommended for higher-risk patients, WBI can be considered depending on patient preference and specific clinical factors. However, it involves a longer treatment course and more significant exposure to surrounding tissues.

#### Decision-Making Process:

- **Patient Preferences:**
    - Discuss the risks and benefits of radiation therapy with the patient, considering her preferences and quality of life.
- **Multidisciplinary Team Discussion:**
    - Involve a multidisciplinary team, including medical oncologists and geriatricians, to tailor the treatment plan to the patient's overall health and preferences.

#### Summary:

Given the favorable pathology and the patient's age, the omission of radiation therapy could be considered reasonable after discussing with the patient and evaluating her overall health status. If radiation therapy is pursued, PBI may be a suitable option to minimize treatment burden and side effects.

It's important to have a thorough discussion with the patient about the potential benefits and risks to make an informed, patient-centered decision.

| | |
|---|---|
| 2<br><br>Inaccurate; omitting radiation therapy is not suggested as the only viable option | For an 80-year-old woman with a new diagnosis of invasive ductal carcinoma, ER/PR positive, HER2 negative, who has undergone a modified radical mastectomy and axillary lymph node dissection, and with final pathology showing a 1.3 cm well-differentiated carcinoma, no lymphovascular invasion, negative margins by >2mm, Ki-67 at 50%, and zero of ten lymph nodes positive, the following considerations for adjuvant radiation therapy are relevant:<br><br>1. **Age and Overall Health:** Given her age, it is important to assess her overall health, comorbidities, and life expectancy to determine if the potential benefits of radiation therapy outweigh the risks and potential side effects.<br><br>2. **Tumor Characteristics:** The tumor is relatively small (1.3 cm), well-differentiated, and has no lymphovascular invasion, which generally indicates a lower risk of recurrence. However, the Ki-67 index of 50% suggests a higher proliferative rate, which may increase the risk of recurrence.<br><br>3. **Lymph Node Status:** With zero of ten lymph nodes positive, the risk of regional recurrence is low. This would generally suggest that radiation therapy might not be necessary, especially given her age and the absence of nodal involvement.<br><br>4. **Radiation Options:** If radiation therapy is considered, it might be limited to the chest wall (post-mastectomy radiation therapy) to reduce the risk of local recurrence. However, given the negative lymph node status and clear margins, omitting radiation might be a reasonable option.<br><br>5. **Patient Preferences:** Discuss the risks and benefits of radiation therapy with the patient, considering her values, preferences, and quality of life.<br><br>**Recommendation:**<br><br>Considering the patient's age, the favorable tumor characteristics (small size, well-differentiated, no lymphovascular invasion, negative margins), and the absence of lymph node involvement, **omission of adjuvant radiation therapy** might be a reasonable option. However, the higher Ki-67 index suggests a need for a careful discussion regarding the potential benefits and risks of radiation therapy.<br><br>In summary, based on current guidelines and the patient's specific case, the recommendation could be to forgo radiation therapy, but this decision should be made in consultation with the patient and her multidisciplinary care team, considering her overall health, preferences, and potential for increased follow-up and surveillance. |

<table>
<tr>
<td>3<br><br>Inaccurate; omitting adjuvant radiation is not suggested as the only viable option</td>
<td>
For an 80-year-old woman with a new diagnosis of invasive ductal carcinoma (IDC), ER/PR positive, HER2 negative, who has undergone a modified radical mastectomy and axillary lymph node dissection with the following final pathology:
<ul>
<li>Tumor size: 1.3 cm</li>
<li>Grade: Well-differentiated</li>
<li>Lymphovascular invasion: None</li>
<li>Margins: Negative by >2mm</li>
<li>Ki-67: 50%</li>
<li>Lymph nodes: 0/10 positive for macrometastases</li>
</ul>
The decision for adjuvant radiation therapy (RT) in this patient would need to consider several factors including the overall health and life expectancy of the patient, the risk of local recurrence, and the potential benefits and risks of RT.<br><br>
Clinical Considerations:
<ol>
<li>Age and Life Expectancy: At 80 years old, the patient's life expectancy and comorbidities should be considered. If the patient is otherwise healthy and has a longer life expectancy, the potential benefits of RT might be more favorable.</li>
<li>Tumor Characteristics:
<ul>
<li>Size and Grade: The tumor size is relatively small (1.3 cm), and it is well-differentiated, which generally suggests a lower risk of recurrence.</li>
<li>Lymph Node Status: No lymph node involvement is a favorable prognostic factor.</li>
<li>Margins: Negative margins by >2mm decrease the likelihood of local recurrence.</li>
<li>Ki-67: A higher Ki-67 (50%) indicates a higher proliferation rate, which might suggest a more aggressive tumor behavior.</li>
</ul>
</li>
</ol>
</td>
</tr>
</table>

| | |
|---|---|
| | 3. **Benefit of Radiation:** Adjuvant radiation therapy can reduce the risk of local recurrence. However, in this case, with small tumor size, no lymphovascular invasion, and no lymph node involvement, the absolute benefit may be relatively small.<br><br>4. **Risks of Radiation:** Consider the risks and potential side effects of radiation, especially in an older patient. These can include skin changes, fatigue, and potential impacts on nearby organs (e.g., lung and heart).<br><br>**Recommendation:**<br><br>Given the patient's favorable tumor characteristics and no lymph node involvement, the decision for adjuvant radiation therapy should be individualized.<br><br>**Options include:**<br><br>1. **Observation without Radiation:** Given the small tumor size, well-differentiated histology, clear margins, and no lymph node involvement, the patient may opt for observation without adjuvant radiation. This option should be considered if the patient has significant comorbidities or prefers to avoid the potential side effects of radiation.<br><br>2. **Adjuvant Radiation Therapy:** For patients who are fit and wish to minimize any risk of local recurrence, adjuvant radiation therapy can be considered, particularly if there is concern about the high Ki-67 indicating a more aggressive tumor.<br><br>**Conclusion:**<br><br>Given the clinical presentation, a discussion with the patient about the risks and benefits of radiation therapy is essential. This conversation should take into account her personal values, preferences, and overall health status. In many cases, given the favorable pathology, observation may be a reasonable approach, but the patient's input is crucial in making this decision.<br><br>Would you like to discuss any specific considerations or details further? |

| | |
|---|---|
| 4<br><br>Inaccurate; omitting radiation therapy is not suggested as the only viable option | For an 80-year-old woman with a new diagnosis of invasive ductal carcinoma, ER/PR positive, HER2 negative, who has undergone a modified radical mastectomy and axillary lymph node dissection, with final pathology showing a 1.3 cm well-differentiated carcinoma, no lymphovascular invasion, margins negative by >2mm, Ki-67 50%, and zero of ten lymph nodes positive for macrometastases, the following considerations and recommendations for adjuvant radiation therapy (RT) are suggested:<br><br>**Clinical Considerations:**<br><br>1. **Age and Comorbidities:**<br>• Given the patient's age, it's important to consider her overall health, life expectancy, and presence of comorbidities. This can impact the decision to proceed with radiation and the potential benefits versus risks.<br>2. **Pathologic Features:**<br>• The tumor size (1.3 cm), well-differentiated nature, absence of lymphovascular invasion, and negative margins are favorable prognostic factors.<br>• The high Ki-67 index (50%) indicates a more aggressive tumor biology.<br>3. **Nodal Status:**<br>• Zero of ten lymph nodes positive for macrometastases is a favorable factor indicating no nodal involvement.<br><br>**Radiation Therapy Recommendations:**<br><br>1. **Chest Wall Radiation:**<br>• Given the small tumor size, favorable histology, and negative nodal status, chest wall radiation may be considered optional, particularly in the context of the patient's age and comorbidities. |

- If the decision is made to treat, standard dose and fractionation schedules (e.g., 50 Gy in 25 fractions or a hypofractionated regimen) can be used.

2. **Regional Nodal Irradiation:**
    - Since there is no lymph node involvement, regional nodal irradiation is not typically indicated.

#### Decision-Making Factors:

- **Shared Decision-Making:**
    - Discuss the potential benefits of adjuvant radiation, including local control and potential survival benefit, balanced against the risks and side effects of radiation therapy.
    - Engage the patient in a thorough discussion, considering her preferences, values, and quality of life.
- **Alternative Approaches:**
    - Consider close surveillance if radiation therapy is deferred, especially given the favorable pathology and lack of nodal involvement.
    - Hormonal therapy may be considered as part of the systemic management for her ER/PR positive disease.

#### Conclusion:

In this case, the decision to proceed with adjuvant radiation therapy should be individualized based on the patient's overall health, preferences, and specific tumor characteristics. Given the favorable pathology, radiation therapy could be considered optional, particularly in light of the patient's advanced age and the potential for comorbid conditions. Close follow-up and consideration of systemic hormonal therapy are also important components of her overall treatment plan.

| | |
|---|---|
| 5<br><br>Inaccurate; omitting adjuvant radiation is not suggested as the only viable option | For an 80-year-old woman with a new diagnosis of invasive ductal carcinoma (IDC), ER/PR positive, HER2 negative, post-modified radical mastectomy and axillary lymph node dissection with the described pathology, here are the considerations for adjuvant radiation therapy:<br><br>**Pathology Review:**<br>• Tumor Size: 1.3 cm<br>• Histology: Well-differentiated IDC<br>• Lymphovascular Invasion: None<br>• Margins: Negative by >2 mm<br>• Ki-67: 50%<br>• Lymph Node Involvement: 0 of 10 nodes positive<br><br>**Key Factors:**<br>1. Age and Comorbidities: The patient is 80 years old. Consider her overall health, comorbidities, and life expectancy.<br>2. Tumor Characteristics: Small, well-differentiated tumor with no lymphovascular invasion and negative margins, indicating a lower risk of local recurrence.<br>3. Lymph Node Status: No lymph node involvement, which also suggests a lower risk of recurrence.<br><br>**Adjuvant Radiation Therapy Considerations:**<br>1. Standard Care: Post-mastectomy radiation therapy (PMRT) is typically recommended for patients with larger tumors or positive lymph nodes. Given the patient's small tumor size and negative lymph nodes, the benefit of PMRT is less clear.<br>2. Individualized Risk Assessment: Given the patient's age, overall health, and pathology report, it may be reasonable to consider omitting PMRT, especially if she has significant comorbidities or limited life expectancy.<br>3. Potential Benefits: While PMRT can reduce the risk of local recurrence, the absolute benefit in this patient might be small given the favorable pathology.<br>4. Potential Risks: Radiation therapy carries risks, including skin toxicity, fatigue, and potential cardiac and pulmonary side effects, which may be more pronounced in older patients.<br><br>**Recommendations:**<br>• Discuss Risks and Benefits: A thorough discussion with the patient and her family about the potential benefits and risks of adjuvant radiation therapy is crucial.<br>• Consider Omission of Radiation: Given the favorable pathology and the patient's age, it may be reasonable to consider omitting radiation therapy.<br>• Alternative Surveillance: If radiation therapy is omitted, ensure a robust follow-up plan with regular physical exams and imaging as appropriate.<br><br>**Conclusion:**<br>In this specific case, the decision to recommend adjuvant radiation therapy should be individualized. Given the patient's age and favorable tumor characteristics, omitting radiation therapy could be a reasonable approach. However, this decision should be made in conjunction with the patient, taking into account her values, preferences, and overall health status. |

| | |
|---|---|
| Phrasing 1 | What specific kind of treatment should be provided for an 80 year old woman with new diagnosis of invasive ductal carcinoma, ER/PR positive, HER2 negative status post modified radical mastectomy and axillary lymph node |

| | |
|---|---|
| | dissection, final pathology demonstrating 1.3cm carcinoma, well differentiated, no lymphovascular invasion, margins negative by >2mm, Ki-67 50%, and zero of ten lymph nodes positive for macrometastases presenting for adjuvant radiation recommendations? |

| | |
|---|---|
| Phrasing 2 | What would NCCN category 1 recommend for an 80 year old woman with new diagnosis of invasive ductal carcinoma, ER/PR positive, HER2 negative status post modified radical mastectomy and axillary lymph node dissection, final pathology demonstrating 1.3cm carcinoma, well differentiated, no lymphovascular invasion, margins negative by >2mm, Ki-67 50%, and zero of ten lymph nodes positive for macrometastases presenting for adjuvant radiation recommendations? |

| | |
|---|---|
| Phrasing 3 | True or false: an 80 year old woman with new diagnosis of invasive ductal carcinoma, ER/PR positive, HER2 negative status post modified radical mastectomy and axillary lymph node dissection, final pathology demonstrating 1.3cm carcinoma, well differentiated, no lymphovascular invasion, margins negative by >2mm, Ki-67 50%, and zero of ten lymph nodes positive for macrometastases presenting for adjuvant radiation recommendations should receive no radiation. |

| | |
|---|---|
| Chain of Reasoning Prompting (Respectively): Question Sequence Ends When Accurate Answer is Given | 1.Considering this woman's age, what would be your adjuvant radiation recommendation?<br>2.Why might you consider omitting adjuvant radiation for this specific woman?<br>3.Based on these considerations, would you still recommend adjuvant radiation for this individual? |

First Example (Anthropic Claude 3.5 Sonnet)

1. 40 year old woman with new diagnosis of invasive ductal carcinoma, ER/PR positive, HER2 negative status post lumpectomy and sentinel lymph node dissection, final pathology demonstrating 1.3cm carcinoma, well differentiated, no lymphovascular invasion, margins negative by >2mm, Ki-67 50%, and all lymph nodes negative, presenting for adjuvant radiation recommendations: **Expected response: Whole breast with boost**

### General Observations

-Similarly to ChatGPT 4, accelerated partial breast irradiation is suggested in one of the five responses; however, when asked if this is a viable option in a separate chat box (provided below), the model displays notable hesitancy due to its considerations of the patient's young age and whole breast irradiation as a better alternative

Accelerated Partial Breast Irradiation (APBI) is an interesting option to consider for this patient, but it comes with some caveats. Let's analyze the pros and cons:

Pros of APBI for this patient:

1. Shorter treatment course: APBI typically takes 1-2 weeks, which is significantly shorter than conventional whole breast irradiation.
2. Targeted treatment: It focuses radiation on the tumor bed, potentially reducing radiation exposure to surrounding healthy tissue.
3. Pathological factors: The tumor size (1.3 cm), negative margins, and negative lymph nodes fit within the typical criteria for APBI candidacy.
4. Convenience: Less time commitment and potentially fewer side effects could be appealing to the patient.

Cons and considerations:

1. Age concern: At 40 years old, this patient is younger than the typically recommended age for APBI. Many guidelines suggest APBI for women 50 or older, as younger women have shown higher rates of local recurrence in some studies.
2. Limited long-term data: While there's growing evidence supporting APBI, long-term follow-up data is still more limited compared to whole breast irradiation, especially in younger patients.
3. Ki-67 of 50%: This high proliferation index suggests a more aggressive tumor biology, which might benefit from whole breast treatment.
4. Potential for microscopic disease: Although margins are negative, there's always a possibility of microscopic disease beyond the immediate tumor bed, which whole breast irradiation would address more comprehensively.
5. Technical considerations: The specific APBI technique (interstitial brachytherapy, balloon brachytherapy, external beam) would need to be carefully chosen based on the patient's anatomy and tumor bed location.

Conclusion:
While APBI could be considered for this patient, her young age (40) and the high Ki-67 index are significant factors that raise caution. The current standard of care for a patient with these characteristics would typically lean towards whole breast irradiation with a boost to the tumor bed.

-5/5 accurate

1

Accurate; whole breast irradiation with boost is suggested

Based on the information provided, I'll outline the key points and provide recommendations for adjuvant radiation therapy for this patient:

Key points:

1. 40-year-old woman
2. Newly diagnosed invasive ductal carcinoma
3. ER/PR positive, HER2 negative
4. Status post lumpectomy and sentinel lymph node dissection
5. Pathology:
   - 1.3 cm carcinoma
   - Well-differentiated
   - No lymphovascular invasion
   - Margins negative by >2mm
   - Ki-67 50%
   - All lymph nodes negative

Adjuvant radiation recommendations:

1. Whole Breast Irradiation (WBI):
   - Standard recommendation for breast-conserving therapy
   - Typically 45-50 Gy in 25 fractions over 5 weeks, or
   - Hypofractionated regimen: 40-42.5 Gy in 15-16 fractions over 3 weeks
2. Boost to the tumor bed:
   - Consider a boost of 10-16 Gy in 4-8 fractions
   - Rationale: Young age (40 years) is associated with higher risk of local recurrence
3. Regional nodal irradiation:
   - Not routinely recommended in this case due to negative lymph nodes and small tumor size
4. Accelerated Partial Breast Irradiation (APBI):
   - May be considered as an alternative to WBI
   - Suitable candidate based on ASTRO criteria (age ≥40, tumor size ≤3cm, negative margins, no LVI)
   - Options include interstitial brachytherapy, balloon brachytherapy, or external beam APBI
5. Timing:
   - Ideally, radiation should start within 4-8 weeks after surgery, if no chemotherapy is planned
   - If chemotherapy is recommended, radiation typically follows completion of chemotherapy

Additional considerations:

- Discuss potential side effects, including skin reactions, fatigue, and long-term effects
- Coordinate with medical oncology regarding sequencing of treatments (e.g., radiation and endocrine therapy)
- Consider genetic testing given the young age at diagnosis
- Encourage participation in clinical trials, if available

These recommendations are based on current guidelines, but the final decision should be made in conjunction with the patient, considering her preferences and any additional factors not mentioned in the provided information.

| | |
|---|---|
| 2<br><br>Accurate; whole breast irradiation with boost is suggested | Based on the information provided, I'll outline the key factors and recommendations for adjuvant radiation therapy for this patient:<br><br>Patient Profile:<br>• 40-year-old woman<br>• New diagnosis of invasive ductal carcinoma<br>• ER/PR positive, HER2 negative<br><br>Surgical Management:<br>• Lumpectomy performed<br>• Sentinel lymph node dissection completed<br><br>Pathology Findings:<br>• 1.3 cm carcinoma<br>• Well-differentiated<br>• No lymphovascular invasion<br>• Margins negative by >2mm<br>• Ki-67 50%<br>• All lymph nodes negative<br><br>Recommendations for Adjuvant Radiation Therapy:<br>1. Whole Breast Irradiation (WBI):<br> • Standard of care following breast-conserving surgery<br> • Typically delivered over 3-6 weeks, depending on the fractionation schedule chosen<br>2. Boost to the Tumor Bed:<br> • Consider a boost to the lumpectomy cavity<br> • Especially important given the patient's young age (40 years old)<br>3. Hypofractionation:<br> • Given the patient's age and tumor characteristics, a hypofractionated regimen could be considered<br> • This involves delivering a higher daily dose over a shorter overall treatment time (typically 3-4 weeks) |

4. Regional Nodal Irradiation:
   - Not routinely recommended in this case, as all lymph nodes were negative
5. Partial Breast Irradiation (PBI):
   - While this patient might be a candidate for PBI based on tumor size and nodal status, her young age (40) may make her less suitable for this approach
   - Discuss the pros and cons if the patient expresses interest in a shorter treatment course
6. Timing:
   - Radiation therapy should typically start within 4-8 weeks after surgery, assuming wound healing is adequate
7. Systemic Therapy Considerations:
   - Coordinate with medical oncology regarding the sequencing of radiation and any planned systemic therapies (e.g., endocrine therapy, chemotherapy)

Additional Considerations:

- Discuss the rationale, benefits, and potential side effects of radiation therapy with the patient
- Address any fertility concerns, given the patient's age
- Encourage the patient to quit smoking if applicable, as it can impact treatment efficacy and side effects

These recommendations are based on current standard practices, but the final decision should be made in conjunction with the patient, considering her preferences and any additional factors not mentioned in the provided information.

Would you like me to elaborate on any specific aspect of these recommendations?

Copy Retry

| | |
|---|---|
| 3<br><br>Accurate; whole breast irradiation with boost is suggested | Based on the information provided, I'll outline the key points and provide recommendations for adjuvant radiation therapy for this patient:<br><br>Key points:<br>1. 40-year-old woman<br>2. Newly diagnosed invasive ductal carcinoma<br>3. ER/PR positive, HER2 negative<br>4. Status post lumpectomy and sentinel lymph node dissection<br>5. Final pathology:<br>• 1.3cm carcinoma<br>• Well differentiated<br>• No lymphovascular invasion<br>• Margins negative by >2mm<br>• Ki-67 50%<br>• All lymph nodes negative<br><br>Adjuvant radiation recommendations:<br>1. Whole Breast Irradiation (WBI): The standard of care for this patient would be adjuvant whole breast irradiation. This is recommended for most patients who have undergone breast-conserving surgery (lumpectomy) to reduce the risk of local recurrence.<br>2. Radiation schedule options: a) Conventional fractionation: 45-50 Gy in 25-28 fractions over 5-6 weeks b) Hypofractionation: 40-42.5 Gy in 15-16 fractions over 3-4 weeks Given the patient's age and favorable tumor characteristics, hypofractionation could be considered as it offers similar efficacy with improved convenience and potentially fewer side effects.<br>3. Boost to the tumor bed: A boost of 10-16 Gy in 4-8 fractions to the tumor bed should be considered, especially given the patient's young age (40 years old). This can further reduce the risk of local recurrence.<br>4. Regional nodal irradiation: Given that all lymph nodes were negative on sentinel lymph node dissection, regional nodal irradiation is not routinely recommended in this case.<br>5. Timing: Radiation therapy should typically start within 4-6 weeks after surgery, assuming wound healing is adequate.<br>6. Systemic therapy considerations: The sequencing of radiation therapy with systemic treatments (e.g., chemotherapy, endocrine therapy) should be discussed with the medical oncologist. If chemotherapy is recommended, it usually precedes radiation therapy.<br>7. Side effect management: Discuss potential side effects and management strategies with the patient, including skin care during treatment.<br>8. Follow-up: Regular follow-up should be scheduled to monitor for any radiation-related side effects and to assess for recurrence.<br><br>These recommendations are based on current standard practices, but the final decision should be made in conjunction with the patient, considering her preferences and any additional factors not mentioned in the provided information. A multidisciplinary tumor board discussion may also be beneficial to ensure comprehensive care.<br><br>Would you like me to elaborate on any specific aspect of these recommendations?<br>Copy Retry |

| | |
|---|---|
| 4<br><br>Accurate; whole breast irradiation with boost is suggested | Based on the information provided, I can offer some insights and recommendations regarding adjuvant radiation therapy for this patient. Here's an analysis of the case and potential treatment approach:<br><br>Patient Profile:<br>• 40-year-old woman<br>• Newly diagnosed invasive ductal carcinoma<br>• ER/PR positive, HER2 negative<br><br>Pathological Findings:<br>• 1.3 cm carcinoma<br>• Well-differentiated<br>• No lymphovascular invasion<br>• Margins negative by >2mm<br>• Ki-67 50%<br>• All lymph nodes negative<br>• Status post lumpectomy and sentinel lymph node dissection<br><br>Recommendations for Adjuvant Radiation Therapy:<br>1. Whole Breast Irradiation (WBI): The standard recommendation for this patient would be whole breast irradiation. This is based on multiple randomized trials showing that breast-conserving surgery followed by radiation therapy offers equivalent survival outcomes to mastectomy for early-stage breast cancer.<br>2. Hypofractionated vs. Conventional Fractionation: Given the patient's age and tumor characteristics, hypofractionated whole breast irradiation could be considered. This involves higher daily doses over a shorter overall treatment time (typically 3-4 weeks) compared to conventional fractionation (5-6 weeks). Studies have shown equivalent outcomes and cosmesis with hypofractionation in appropriate patients.<br>3. Boost to the Tumor Bed: A boost to the tumor bed should be considered, especially given the patient's young age (40 years). The EORTC boost vs. no boost trial demonstrated a reduction in local recurrence rates with the addition of a boost, particularly in younger patients.<br>4. Regional Nodal Irradiation: Given that all lymph nodes were negative, regional nodal irradiation is not routinely recommended in this case.<br>5. Partial Breast Irradiation: While partial breast irradiation might be an option for some early-stage breast cancer patients, this patient's young age and high Ki-67 (50%) may make her a less ideal candidate for this approach.<br>6. Timing: Radiation therapy typically starts within 4-6 weeks after surgery, assuming wound healing is adequate.<br><br>Additional Considerations:<br>• The high Ki-67 (50%) suggests a more proliferative tumor, which might influence systemic therapy decisions.<br>• Given the ER/PR positive status, endocrine therapy would likely be recommended in addition to radiation.<br>• The patient's age and tumor characteristics might also warrant a discussion about chemotherapy with medical oncology.<br><br>It's important to note that these recommendations should be discussed in a multidisciplinary tumor board and tailored to the patient's individual circumstances, preferences, and any additional factors not mentioned in this brief case description.<br><br>Would you like me to elaborate on any specific aspect of these recommendations?<br><br>Copy Retry |

| | |
|---|---|
| 5<br><br>Accurate; whole breast irradiation with boost is suggested | Based on the information provided, I'll offer some thoughts on adjuvant radiation recommendations for this patient. However, please note that final treatment decisions should be made in consultation with the patient's oncology team, taking into account her full medical history and preferences.<br><br>For this 40-year-old woman with early-stage breast cancer, adjuvant radiation therapy is generally recommended. Here's a breakdown of the key factors and recommendations:<br><br>1. Tumor characteristics:<br>• Invasive ductal carcinoma<br>• ER/PR positive, HER2 negative<br>• 1.3 cm tumor size<br>• Well-differentiated<br>• No lymphovascular invasion<br>• Ki-67 50% (indicating high proliferation rate)<br>2. Surgical outcome:<br>• Lumpectomy performed (breast-conserving surgery)<br>• Negative margins (>2mm)<br>• Sentinel lymph node dissection with all nodes negative<br>3. Radiation therapy recommendations: a) Whole Breast Irradiation (WBI):<br>• Standard recommendation after lumpectomy<br>• Typically delivered over 3-6 weeks, depending on the fractionation schedule chosen<br>b) Boost to the tumor bed:<br>• Often recommended, especially for younger patients (age <50)<br>• Can help further reduce local recurrence risk<br>c) Regional nodal irradiation:<br>• Not typically indicated in this case, given the negative sentinel lymph nodes<br>d) Accelerated Partial Breast Irradiation (APBI):<br>• Might be considered as an alternative to WBI in select low-risk patients<br>• However, given the patient's young age and high Ki-67, standard WBI would likely be preferred<br>4. Additional considerations:<br>• The patient's young age (40) is a factor that generally favors more aggressive treatment, as younger patients have a higher risk of recurrence<br>• The high Ki-67 (50%) indicates a more rapidly proliferating tumor, which might influence the decision towards standard WBI over APBI<br>5. Potential radiation schedule options:<br>• Conventional fractionation: ~50 Gy in 25 fractions over 5 weeks, followed by a boost<br>• Hypofractionation: ~40-42.5 Gy in 15-16 fractions over 3 weeks, followed by a boost<br>• The choice between these schedules would depend on institutional preferences and patient factors<br><br>In summary, for this patient, whole breast irradiation with a boost to the tumor bed would be the most commonly recommended approach. The specific schedule and any additional considerations should be discussed with the patient, taking into account her preferences, lifestyle, and any relevant medical history not provided in this scenario.<br><br>Would you like me to elaborate on any specific aspect of these recommendations?<br><br>Copy Retry |

Second Example (Anthropic Claude 3.5 Sonnet)

2. 40 year old woman with new diagnosis of invasive ductal carcinoma, ER/PR positive, HER2 negative status post modified radical mastectomy and axillary lymph node dissection, final pathology demonstrating 1.3cm carcinoma, well differentiated, no lymphovascular invasion, margins negative by >2mm, Ki-67 50%, and five of ten lymph nodes positive for macrometastases. Presenting for adjuvant radiation recommendations:
**Expected response: Regional Nodal Irradiation**

### General Observations

-Responses #1 & #5 state the axillary region as a target location for regional nodal irradiation (without mention of the infraclavicular fossa), whereas the other responses only suggest the supraclavicular fossa, infraclavicular fossa, and internal mammary nodes
-5/5 accurate

<table>
<tr>
<td>1<br><br>Accurate; regional nodal irradiation is suggested</td>
<td>

Based on the information provided, this patient has several factors that indicate a need for adjuvant radiation therapy. Let's break down the case and discuss the recommendations:

Patient characteristics:

- 40-year-old woman
- New diagnosis of invasive ductal carcinoma
- ER/PR positive, HER2 negative
- Status post modified radical mastectomy and axillary lymph node dissection

Pathology findings:

- 1.3 cm carcinoma
- Well differentiated
- No lymphovascular invasion
- Margins negative by >2mm
- Ki-67 50%
- 5 out of 10 lymph nodes positive for macrometastases

Recommendations for adjuvant radiation therapy:

1. Post-mastectomy radiation therapy (PMRT): This patient is a strong candidate for PMRT due to the following factors:
   - Positive lymph nodes (5 out of 10)
   - Young age (40 years old)

   PMRT has been shown to reduce the risk of locoregional recurrence and improve overall survival in patients with positive lymph nodes after mastectomy.
2. Treatment areas:
   - Chest wall
   - Supraclavicular fossa
   - Internal mammary nodes (consider based on institutional practice)
   - Axillary region (levels I-III, given the extent of nodal involvement)
3. Radiation dose and fractionation:
   - Standard fractionation: 50 Gy in 25 fractions over 5 weeks
   - Consider a boost to the mastectomy scar: 10 Gy in 5 fractions
4. Timing:
   - Ideally, radiation should begin within 4-6 weeks after surgery or completion of adjuvant chemotherapy, if chemotherapy is planned.
5. Additional considerations:
   - Given the ER/PR positive status, endocrine therapy should be discussed.
   - The high Ki-67 (50%) suggests a more aggressive tumor, which further supports the need for comprehensive adjuvant treatment.
   - Genetic counseling may be considered due to the young age at diagnosis.
6. Potential side effects to discuss:
   - Skin reactions
   - Fatigue
   - Potential for lymphedema (given axillary dissection and radiation)
   - Very low risk of pneumonitis or cardiac toxicity (depending on the side affected)

It's important to note that these recommendations should be discussed in a multidisciplinary tumor board setting, taking into account the patient's overall health, preferences, and any planned systemic therapy. The final decision should be made in consultation with the patient, explaining the potential benefits and risks of adjuvant radiation therapy.

Would you like me to elaborate on any specific aspect of these recommendations?

</td>
</tr>
</table>

| | |
|---|---|
| 2<br><br>Accurate; regional nodal irradiation is suggested | Based on the information provided, here's an assessment and recommendation for adjuvant radiation therapy for this patient:<br><br>Patient profile:<br>• 40-year-old woman<br>• Newly diagnosed invasive ductal carcinoma<br>• ER/PR positive, HER2 negative<br>• Status post modified radical mastectomy and axillary lymph node dissection<br><br>Pathological findings:<br>• 1.3cm carcinoma<br>• Well differentiated<br>• No lymphovascular invasion<br>• Margins negative by >2mm<br>• Ki-67 50%<br>• 5/10 lymph nodes positive for macrometastases<br><br>Recommendation:<br>This patient would benefit from adjuvant radiation therapy. The rationale for this recommendation includes:<br>1. Lymph node involvement: With 5 out of 10 lymph nodes positive for macrometastases, the patient has significant nodal involvement. This increases the risk of locoregional recurrence and is a strong indication for post-mastectomy radiation therapy (PMRT).<br>2. Age: At 40 years old, the patient is relatively young, which is associated with a higher risk of recurrence.<br>3. Tumor characteristics: Although the tumor is small (1.3cm) and well-differentiated with negative margins, the high Ki-67 (50%) indicates a high proliferation rate, which is associated with more aggressive behavior.<br><br>Proposed radiation plan:<br>1. Chest wall irradiation<br>2. Supraclavicular and infraclavicular nodal regions<br>3. Internal mammary nodal region (consider based on tumor location and institutional practice)<br><br>Typical dose: 50 Gy in 25 fractions over 5 weeks, with a possible boost to the chest wall scar.<br><br>Additional considerations:<br>1. Timing: Radiation therapy should typically start within 4-6 weeks after surgery or after completion of adjuvant chemotherapy if prescribed.<br>2. Systemic therapy: Given the ER/PR positive status, endocrine therapy should be discussed. The high Ki-67 and nodal involvement may also warrant consideration of adjuvant chemotherapy.<br>3. Side effect management: Discuss potential side effects such as skin reactions, fatigue, and long-term risks, including lymphedema and rare cardiac toxicity.<br>4. Follow-up: Regular follow-up should be scheduled to monitor for recurrence and manage any late effects of treatment.<br><br>It's important to discuss this recommendation with the patient, explaining the benefits and potential risks of adjuvant radiation therapy, and to consider her preferences in the decision-making process. A multidisciplinary tumor board discussion may also be beneficial to optimize the overall treatment plan. |

| | |
|---|---|
| 3<br><br>Accurate; regional nodal irradiation is suggested | Based on the information provided, this patient has several factors that warrant consideration for adjuvant radiation therapy. Let's break down the key elements and discuss the recommendations:<br><br>Patient characteristics:<br>• 40-year-old woman<br>• New diagnosis of invasive ductal carcinoma<br>• ER/PR positive, HER2 negative<br><br>Surgical intervention:<br>• Modified radical mastectomy<br>• Axillary lymph node dissection<br><br>Pathology findings:<br>• Tumor size: 1.3 cm<br>• Well-differentiated<br>• No lymphovascular invasion<br>• Margins negative by >2mm<br>• Ki-67: 50%<br>• Lymph node status: 5 out of 10 positive for macrometastases<br><br>Given these factors, adjuvant radiation therapy is strongly recommended for this patient. The main reasons for this recommendation are:<br><br>1. Positive lymph nodes: With 5 out of 10 lymph nodes positive for macrometastases, this patient has significant nodal involvement. Post-mastectomy radiation therapy (PMRT) has been shown to reduce locoregional recurrence and improve overall survival in patients with positive lymph nodes.<br>2. Young age: At 40 years old, the patient is considered young for breast cancer diagnosis. Younger age is associated with a higher risk of recurrence, which further supports the use of radiation therapy.<br>3. High Ki-67: A Ki-67 of 50% indicates a high proliferation rate, which is associated with a more aggressive tumor behavior and higher risk of recurrence. |

| | |
|---|---|
| | Recommended radiation therapy approach:<br>1. Chest wall irradiation: To reduce the risk of local recurrence.<br>2. Regional nodal irradiation: Including the supraclavicular and infraclavicular regions, as well as the internal mammary nodes. This is particularly important given the extent of nodal involvement.<br>3. Boost to the mastectomy scar: Consider a boost to the mastectomy scar, especially given the patient's young age.<br>The radiation oncologist should discuss the specific radiation doses and fractionation schedule with the patient, taking into account factors such as institutional protocols and patient preferences.<br>It's important to note that adjuvant radiation therapy should be coordinated with other adjuvant treatments, such as chemotherapy and endocrine therapy, which are likely to be recommended given the tumor's ER/PR positive status and the extent of nodal involvement.<br>Lastly, the radiation oncologist should discuss the potential benefits and risks of radiation therapy with the patient, including the impact on recurrence risk, overall survival, and potential side effects.<br>Would you like me to elaborate on any specific aspect of these recommendations or discuss additional considerations for this patient's care? |

| | |
|---|---|
| 4<br><br>Accurate; regional nodal irradiation is suggested | Based on the information provided, this patient has several factors that indicate a need for adjuvant radiation therapy. Let's break down the case and discuss the radiation recommendations:<br><br>Patient characteristics:<br>• 40-year-old woman<br>• New diagnosis of invasive ductal carcinoma<br>• ER/PR positive, HER2 negative<br>• Status post modified radical mastectomy and axillary lymph node dissection<br><br>Pathology findings:<br>• 1.3 cm carcinoma<br>• Well-differentiated<br>• No lymphovascular invasion<br>• Margins negative by >2mm<br>• Ki-67 50%<br>• 5 out of 10 lymph nodes positive for macrometastases<br><br>Radiation therapy recommendations:<br>1. Post-mastectomy radiation therapy (PMRT): This patient is a strong candidate for PMRT due to the following factors:<br> • Positive lymph nodes (5 out of 10)<br> • Young age (40 years old)<br> • High Ki-67 (50%), indicating an aggressive tumor<br>2. Treatment areas:<br> • Chest wall<br> • Supraclavicular fossa<br> • Internal mammary lymph nodes (consideration based on institutional practice)<br>3. Radiation dose and fractionation:<br> • Standard fractionation: 50 Gy in 25 fractions over 5 weeks<br> • Hypofractionation can be considered: 40-42.5 Gy in 15-16 fractions over 3 weeks<br>4. Boost:<br> • A boost to the mastectomy scar is generally not recommended unless there are close margins or other high-risk features<br>5. Timing:<br> • Radiation therapy should typically start within 4-6 weeks after surgery or after completion of adjuvant chemotherapy if indicated<br><br>Additional considerations:<br>• Systemic therapy: Given the ER/PR positive status, endocrine therapy will likely be recommended. The high Ki-67 and positive lymph nodes may also indicate a benefit from adjuvant chemotherapy.<br>• Genetic counseling: Consider referring the patient for genetic counseling due to her young age at diagnosis.<br><br>It's important to note that these recommendations should be discussed in a multidisciplinary tumor board and tailored to the patient's individual circumstances, including her overall health, preferences, and any planned systemic therapies.<br><br>Would you like me to elaborate on any specific aspect of these recommendations? |

| | |
|---|---|
| 5<br><br>Accurate; regional nodal irradiation is suggested | Based on the information provided, this patient has several high-risk features that warrant consideration for adjuvant radiation therapy. Let's break down the key factors and discuss the radiation recommendations:<br><br>Patient and Disease Characteristics:<br>1. 40-year-old woman (younger age is a risk factor)<br>2. Invasive ductal carcinoma<br>3. ER/PR positive, HER2 negative<br>4. Post modified radical mastectomy and axillary lymph node dissection<br>5. Tumor size: 1.3 cm<br>6. Well-differentiated<br>7. No lymphovascular invasion<br>8. Margins negative by >2mm<br>9. Ki-67: 50% (high proliferation index)<br>10. Lymph node status: 5 of 10 positive for macrometastases<br><br>Radiation Therapy Recommendations:<br><br>Given the patient's characteristics and pathology results, adjuvant radiation therapy is strongly recommended. The primary reasons for this recommendation are:<br>1. Positive lymph nodes: With 5 out of 10 lymph nodes positive for macrometastases, this patient has N2 disease. Multiple positive nodes significantly increase the risk of locoregional recurrence and are a strong indication for post-mastectomy radiation therapy (PMRT).<br>2. Young age: At 40 years old, the patient is considered young for breast cancer diagnosis, which is associated with a higher risk of recurrence.<br>3. High Ki-67: A Ki-67 of 50% indicates a high proliferation rate, suggesting a more aggressive tumor behavior. |

Recommended Radiation Fields:

1. Chest wall: To reduce the risk of local recurrence
2. Supraclavicular lymph nodes: Due to the high nodal burden
3. Internal mammary lymph nodes: Consider including based on tumor location and institutional practice
4. Axillary lymph nodes: Given the extensive nodal involvement

Radiation Dose and Fractionation:
The standard approach would be:

- 50 Gy in 25 fractions to all fields
- Consider a boost to the chest wall scar of 10 Gy in 5 fractions

Alternative hypofractionated schedules could be considered based on institutional practices and patient factors.

Additional Considerations:

1. Timing: Radiation therapy should ideally begin within 4-6 weeks after completion of chemotherapy or 4-8 weeks post-surgery if chemotherapy is not given.
2. Systemic Therapy: Given the ER/PR positive status, endocrine therapy should be recommended. The high-risk features also warrant consideration of adjuvant chemotherapy.
3. Side Effect Management: Discuss potential side effects and management strategies with the patient.
4. Follow-up: Regular follow-up should be scheduled to monitor for recurrence and manage any late effects of treatment.

In conclusion, post-mastectomy radiation therapy is strongly recommended for this patient due to the presence of multiple high-risk factors, particularly the positive lymph nodes. A multidisciplinary approach involving surgical oncology, medical oncology, and radiation oncology is crucial for optimal management.

Third Example (Anthropic Claude 3.5 Sonnet)

3. 80 year old woman with new diagnosis of invasive ductal carcinoma, ER/PR positive, HER2 negative status post modified radical mastectomy and axillary lymph node dissection, final pathology demonstrating 1.3cm carcinoma, well differentiated, no lymphovascular invasion, margins negative by >2mm, Ki-67 50%, and zero of ten lymph nodes positive for macrometastases. Presenting for adjuvant radiation recommendations:
**Expected response: No radiation**

### General Observations

-Response #5, similarly to many of ChatGPT 4's responses, display notable concern

about the patient’s high Ki-67 index, suggesting that the aggressive biology of the cancer should be discussed when considering adjuvant radiation therapy, perhaps tipping the decision to favor radiotherapy
-Again, the model erroneously implies or states that adjuvant radiation may be a consideration
-0/5 accurate
-⅖ partially accurate

Phrasing 1:

Phrasing 2:

Phrasing 3:

Chain of Reasoning Prompting:

| | |
|---|---|
| 1<br><br>Partially accurate; although omitting adjuvant radiation is suggested, the model implies that - while not recommended - radiotherapy may be a consideration | Based on the information provided, I'll outline the key factors to consider for adjuvant radiation recommendations for this patient:<br>1. Patient characteristics:<br>• 80-year-old woman<br>• New diagnosis of invasive ductal carcinoma<br>2. Tumor characteristics:<br>• ER/PR positive<br>• HER2 negative<br>• 1.3 cm carcinoma<br>• Well differentiated<br>• No lymphovascular invasion<br>• Margins negative by >2mm<br>• Ki-67 50% (indicating high proliferation rate)<br>3. Lymph node status:<br>• 0/10 lymph nodes positive for macrometastases<br>4. Prior treatment:<br>• Modified radical mastectomy<br>• Axillary lymph node dissection<br>Given these factors, here are the considerations for adjuvant radiation therapy:<br>1. Post-mastectomy radiation therapy (PMRT):<br>• Generally, PMRT is not routinely recommended for patients with T1-2 tumors and negative lymph nodes.<br>• This patient's tumor is small (1.3 cm, T1c) with negative lymph nodes, which suggests a lower risk of locoregional recurrence.<br>2. Age consideration:<br>• At 80 years old, the patient's life expectancy and ability to tolerate treatment should be carefully evaluated.<br>• The potential benefits of radiation therapy should be weighed against the risks and quality of life impact. |

3. Risk factors:
   - The high Ki-67 (50%) indicates a more aggressive tumor biology, which might be considered a risk factor.
   - However, other factors (well-differentiated, negative margins, no lymphovascular invasion) are favorable.
4. Guidelines:
   - Most guidelines (NCCN, ASCO, ASTRO) do not recommend routine PMRT for patients with T1-2 tumors and negative nodes after mastectomy.
5. Alternative considerations:
   - Given the ER/PR positive status, adjuvant endocrine therapy would likely be recommended, which can further reduce the risk of local recurrence.

Recommendation:
Based on the available information, adjuvant radiation therapy is likely not recommended for this patient. The small tumor size, negative lymph nodes, and negative margins suggest a low risk of locoregional recurrence. The potential benefits of radiation therapy may not outweigh the risks and impact on quality of life, especially considering the patient's age.

However, the final decision should be made after:

1. Discussing the potential risks and benefits with the patient
2. Considering the patient's overall health status and life expectancy
3. Evaluating the patient's preferences and goals of care
4. Consulting with a multidisciplinary tumor board, if available

It's important to note that adjuvant endocrine therapy should be strongly considered given the ER/PR positive status, as it can further reduce the risk of recurrence and improve overall survival.

Would you like me to elaborate on any specific aspect of this recommendation or discuss alternative approaches?

Copy Retry

| | |
|---|---|
| 2<br><br>Inaccurate; omitting adjuvant radiation is not suggested as the only viable option | Based on the information provided, let's analyze this case and consider the recommendations for adjuvant radiation therapy:<br><br>Patient profile:<br>• 80-year-old woman<br>• New diagnosis of invasive ductal carcinoma<br>• ER/PR positive, HER2 negative<br>• Post modified radical mastectomy and axillary lymph node dissection<br><br>Pathology findings:<br>• Tumor size: 1.3 cm<br>• Well-differentiated<br>• No lymphovascular invasion<br>• Margins negative by >2mm<br>• Ki-67: 50%<br>• Lymph node status: 0/10 positive for macrometastases<br><br>Given these factors, here are some considerations regarding adjuvant radiation therapy:<br>1. Tumor characteristics: The tumor is relatively small (1.3 cm), well-differentiated, and has no lymphovascular invasion. These are generally favorable prognostic factors.<br>2. Lymph node status: The patient has node-negative disease (0/10 lymph nodes positive), which is associated with a better prognosis.<br>3. Surgical margins: The margins are negative by >2mm, which is considered adequate.<br>4. Age: At 80 years old, the patient's age is a significant factor in decision-making, as the risks and benefits of treatment must be carefully weighed.<br>5. Hormone receptor status: The tumor is ER/PR positive, which generally indicates a better prognosis and responsiveness to hormonal therapy.<br>6. Ki-67: The high Ki-67 (50%) suggests a more rapidly proliferating tumor, which could be a concern. |

Recommendations:

1. Omission of radiation therapy may be considered:
   - The CALGB 9343 trial showed that in women ≥70 years with T1N0 ER-positive breast cancer treated with lumpectomy and tamoxifen, the addition of radiation therapy did not improve overall survival or distant disease-free survival.
   - The patient's age, small tumor size, and node-negative status align with this study population.
2. If radiation therapy is considered:
   - Whole breast irradiation with a hypofractionated regimen could be an option if breast-conserving surgery had been performed.
   - In this case of mastectomy, chest wall irradiation might be considered, but the indications are less clear given the node-negative status and small tumor size.
3. Alternative approach:
   - Given the patient's age and the tumor characteristics, adjuvant endocrine therapy alone might be a reasonable option, focusing on reducing the risk of recurrence while minimizing treatment-related side effects.
4. Multidisciplinary discussion:
   - The decision should involve a multidisciplinary team, including the surgeon, medical oncologist, radiation oncologist, and geriatric specialist if available.
   - The patient's overall health status, life expectancy, and personal preferences should be taken into account.
5. Genomic testing:
   - Consider genomic assays (e.g., Oncotype DX) to further assess the risk of recurrence and potential benefit from chemotherapy, which could influence the decision on radiation therapy.

In conclusion, while adjuvant radiation therapy can be considered, it may be reasonable to omit it in this case, focusing instead on adjuvant endocrine therapy. The decision should be made after careful discussion with the patient, considering her overall health status, preferences, and the potential risks and benefits of treatment.

Would you like me to elaborate on any specific aspect of these recommendations?

| | |
|---|---|
| 3<br><br>Inaccurate; omitting radiation therapy is not suggested as the only viable option | Based on the information provided, I'll discuss the case and provide recommendations for adjuvant radiation therapy for this 80-year-old woman with early-stage breast cancer. Let's break down the key points and then consider the radiation therapy options:<br><br>Key points:<br>1. Age: 80 years old<br>2. Diagnosis: Invasive ductal carcinoma<br>3. Receptor status: ER/PR positive, HER2 negative<br>4. Surgery: Modified radical mastectomy with axillary lymph node dissection<br>5. Pathology:<br>• Tumor size: 1.3 cm<br>• Grade: Well-differentiated<br>• No lymphovascular invasion<br>• Negative margins (>2mm)<br>• Ki-67: 50%<br>• Lymph node status: 0/10 positive<br><br>Given these factors, this patient has stage I (T1N0M0) breast cancer. The primary considerations for adjuvant radiation therapy in this case are:<br>1. Age: At 80 years old, the patient's life expectancy and ability to tolerate treatment should be considered.<br>2. Mastectomy: Post-mastectomy radiation therapy (PMRT) is typically recommended for patients with larger tumors (>5 cm) or positive lymph nodes, neither of which apply in this case.<br>3. Negative lymph nodes and small tumor size: These factors suggest a lower risk of locoregional recurrence.<br>4. Well-differentiated tumor: This is associated with a better prognosis.<br>5. High Ki-67 (50%): This indicates a more proliferative tumor, which could suggest a more aggressive disease despite the small size and negative nodes.<br><br>Recommendations:<br>1. Omission of radiotherapy: Given the patient's age, early-stage disease, negative lymph nodes, and negative margins after mastectomy, it would be reasonable to omit adjuvant radiation therapy. The potential benefits of radiation in this case may not outweigh the risks and burden of treatment for an 80-year-old patient.<br>2. Consider chest wall radiation: If there are concerns about the high Ki-67 or other patient-specific factors that might increase recurrence risk, chest wall radiation could be considered. However, this would be less common in this scenario.<br>3. Discuss alternatives: Instead of radiation therapy, ensure that appropriate systemic therapy (e.g., endocrine therapy given the ER/PR positive status) is prescribed to reduce the risk of distant recurrence and potentially local recurrence.<br>4. Shared decision-making: Discuss the pros and cons of adjuvant radiation therapy with the patient, considering her preferences, overall health status, and quality of life goals.<br>5. Follow-up: Recommend close follow-up with regular clinical exams and imaging as appropriate, regardless of whether radiation is administered.<br><br>In conclusion, based on the available information, omitting adjuvant radiation therapy would be a reasonable approach for this patient. However, the final decision should be made after a thorough discussion with the patient, considering her overall health status, preferences, and a multidisciplinary tumor board review if available.<br><br>Would you like me to elaborate on any specific aspect of these recommendations or discuss alternative approaches? |

| | |
|---|---|
| 4<br><br>Partially accurate; whilst omitting radiation therapy is suggested, the model implies that adjuvant radiotherapy (though not recommended) may be a consideration | Based on the information provided, let's analyze this case and consider the recommendations for adjuvant radiation therapy:<br><br>Key points:<br>1. 80-year-old woman<br>2. Invasive ductal carcinoma<br>3. ER/PR positive, HER2 negative<br>4. Post modified radical mastectomy and axillary lymph node dissection<br>5. Tumor characteristics:<br>• 1.3 cm carcinoma<br>• Well differentiated<br>• No lymphovascular invasion<br>• Margins negative by >2mm<br>• Ki-67 50%<br>6. 0/10 lymph nodes positive for macrometastases<br><br>Analysis:<br>This patient has early-stage breast cancer with favorable prognostic factors, including small tumor size, negative lymph nodes, and hormone receptor positivity. However, the high Ki-67 (50%) suggests a more aggressive tumor biology.<br><br>Recommendations for adjuvant radiation therapy:<br>1. Post-mastectomy radiation therapy (PMRT) is generally not recommended for patients with T1-2 tumors and negative lymph nodes (N0) who have undergone mastectomy. This patient falls into this category.<br>2. The NCCN (National Comprehensive Cancer Network) guidelines typically do not recommend PMRT for patients with T1-2N0 disease.<br>3. However, individual factors should be considered, such as: a. The patient's age (80 years old) b. The high Ki-67 index (50%) c. Overall health status and life expectancy d. Patient preferences<br>4. Given the patient's age and the absence of high-risk features (such as positive lymph nodes or larger tumor size), the potential benefits of radiation therapy may be outweighed by the risks and inconvenience, especially considering life expectancy.<br>5. Instead of radiation therapy, focus should be on adjuvant endocrine therapy, given the ER/PR positive status. This would likely provide more benefit in terms of reducing recurrence risk with less potential for side effects.<br><br>Conclusion:<br>Based on the available information, adjuvant radiation therapy is likely not recommended for this patient. The focus should be on adjuvant endocrine therapy. However, the final decision should be made after a thorough discussion with the patient, considering her overall health status, preferences, and a multidisciplinary tumor board review if available.<br><br>It's important to note that this recommendation is based on general guidelines and the information provided. The treating radiation oncologist should make the final decision after a comprehensive evaluation of the patient and consideration of any additional relevant factors not mentioned here.<br><br>Would you like me to elaborate on any aspect of this recommendation or discuss alternative approaches? |

| | |
|---|---|
| 5<br><br>Inaccurate; omitting adjuvant radiation is not suggested as the only viable option | Thank you for providing this detailed case. Let's break down the key information and consider the recommendations for adjuvant radiation therapy for this patient:<br><br>Patient profile:<br>• 80-year-old woman<br>• New diagnosis of invasive ductal carcinoma<br>• ER/PR positive, HER2 negative<br><br>Surgical intervention:<br>• Modified radical mastectomy<br>• Axillary lymph node dissection<br><br>Pathology findings:<br>• 1.3 cm carcinoma<br>• Well-differentiated<br>• No lymphovascular invasion<br>• Margins negative by >2mm<br>• Ki-67 50%<br>• 0/10 lymph nodes positive for macrometastases<br><br>Given these factors, here are the considerations for adjuvant radiation therapy:<br>1. Age: At 80 years old, the patient's age and overall health status should be taken into account when making treatment decisions.<br>2. Tumor size and node status: The tumor is relatively small (1.3 cm) and node-negative, which are favorable prognostic factors.<br>3. Surgical margins: Negative margins by >2mm reduce the risk of local recurrence.<br>4. Tumor biology: ER/PR positive status suggests a more favorable prognosis, but the high Ki-67 (50%) indicates a more aggressive tumor.<br><br>Recommendations:<br>1. Post-mastectomy radiation therapy (PMRT) is generally not recommended for patients with T1-2N0 breast cancer who have undergone mastectomy. This patient falls into this category (T1N0).<br>2. However, the high Ki-67 and the patient's relatively younger age (for an octogenarian) might warrant a discussion about the potential benefits of radiation therapy to reduce the risk of local recurrence.<br>3. The decision should be made after a multidisciplinary tumor board discussion and should take into account the patient's overall health status, life expectancy, and preferences.<br>4. If radiation therapy is considered, it would likely involve treatment to the chest wall and possibly the regional lymph nodes.<br>5. The potential benefits of radiation therapy should be weighed against the risks and side effects, especially considering the patient's age.<br>6. Adjuvant endocrine therapy (e.g., aromatase inhibitors or tamoxifen) should be strongly considered given the ER/PR positive status.<br>7. A genomic assay (e.g., Oncotype DX) might be helpful in further stratifying the risk and guiding decisions about adjuvant chemotherapy, although the benefit may be limited given the patient's age.<br><br>In conclusion, while post-mastectomy radiation therapy is not routinely recommended for patients with T1N0 breast cancer, the high Ki-67 in this case warrants a thorough discussion of the potential benefits and risks. The final decision should be made in conjunction with the patient, taking into account her overall health status, preferences, and the recommendations of a multidisciplinary team.<br><br>Would you like me to elaborate on any specific aspect of these recommendations? |

| | |
|---|---|
| Phrasing 1 | What specific kind of treatment should be provided for an 80 year old woman with new diagnosis of invasive ductal carcinoma, ER/PR positive, HER2 negative status post modified radical mastectomy and axillary lymph node dissection, final pathology demonstrating 1.3cm carcinoma, well differentiated, no lymphovascular invasion, margins negative by >2mm, Ki-67 50%, and zero of ten lymph nodes positive for macrometastases presenting for adjuvant radiation recommendations? |

| | |
|---|---|
| Phrasing 2 | What would NCCN category 1 recommend for an 80 year old woman with new diagnosis of invasive ductal carcinoma, ER/PR positive, HER2 negative status post modified radical mastectomy and axillary lymph node dissection, final pathology demonstrating 1.3cm carcinoma, well differentiated, no lymphovascular invasion, margins negative by >2mm, Ki-67 50%, and zero of ten lymph nodes positive for macrometastases presenting for adjuvant radiation recommendations? |

| | |
|---|---|
| Phrasing 3 | True or false: an 80 year old woman with new diagnosis of invasive ductal carcinoma, ER/PR positive, HER2 negative status post modified radical mastectomy and axillary lymph |

| | |
|---|---|
| | node dissection, final pathology demonstrating 1.3cm carcinoma, well differentiated, no lymphovascular invasion, margins negative by >2mm, Ki-67 50%, and zero of ten lymph nodes positive for macrometastases presenting for adjuvant radiation recommendations should receive no radiation. |

| | |
|---|---|
| Chain of Reasoning Prompting (Respectively): Question Sequence Ends When Accurate Answer is Given | 1.Considering this woman's age, what would be your adjuvant radiation recommendation?<br>2.Why might you consider omitting adjuvant radiation for this specific woman?<br>3.Based on these considerations, would you still recommend adjuvant radiation for this individual? |

First Example (ChatGPT 4o-Mini)

1. 40 year old woman with new diagnosis of invasive ductal carcinoma, ER/PR positive, HER2 negative status post lumpectomy and sentinel lymph node dissection, final pathology demonstrating 1.3cm carcinoma, well differentiated, no lymphovascular invasion, margins negative by >2mm, Ki-67 50%, and all lymph nodes negative, presenting for adjuvant radiation recommendations: **Expected response: Whole breast with boost**

Focus only on inputting the initial prompt multiple times (5 times) and showing that it does not consistently give the NCCN Category 1 answer.

| |
|---|
| General Observations<br><br>- |

| | |
|---|---|
| 1<br><br>Accurate; whole breast with boost is recommended | Given the patient's diagnosis of invasive ductal carcinoma with the provided details, here's a summary of the key aspects and the typical recommendations for adjuvant radiation therapy:<br><br>**Patient Profile:**<br>• **Age:** 40 years old<br>• **Diagnosis:** Invasive ductal carcinoma (IDC), ER/PR positive, HER2 negative<br>• **Post-surgery status:** Lumpectomy and sentinel lymph node dissection<br>• **Pathology Results:**<br> ◦ **Tumor Size:** 1.3 cm<br> ◦ **Grade:** Well differentiated<br> ◦ **Lymphovascular Invasion:** None<br> ◦ **Margins:** Negative by >2mm<br> ◦ **Ki-67:** 50% (high proliferation index)<br> ◦ **Lymph Nodes:** All negative<br><br>**Adjuvant Radiation Therapy Recommendations:**<br>1. **Whole-Breast Radiation Therapy:**<br> ◦ **Indication:** Given that the patient had a lumpectomy, whole-breast radiation is generally recommended to reduce the risk of local recurrence. This is particularly important in early-stage breast cancer, even with negative margins and no lymphovascular invasion.<br> ◦ **Typical Regimen:** Often, this involves 40-50 Gy delivered in daily fractions over 4-6 weeks.<br>2. **Boost Radiation Therapy:**<br> ◦ **Indication:** Consideration for a radiation boost to the tumor bed is based on the size of the tumor and other risk factors. In this case, given the tumor size of 1.3 cm and high Ki-67, a boost to the lumpectomy site (usually an additional 10-16 Gy) may be recommended to further reduce the risk of recurrence in the area where the tumor was removed. |

| | |
|---|---|
| | 3. **Special Considerations:**<br>• **Ki-67 Index:** The high Ki-67 index (50%) suggests a higher proliferation rate, which can be associated with a higher risk of recurrence. While this doesn't change the fundamental approach to radiation, it might influence discussions about the intensity of additional treatments or surveillance.<br>• **Patient-Specific Factors:** Consideration should be given to the patient's overall health, preferences, and potential side effects.<br>4. **Discussion of Benefits and Risks:**<br>• **Benefits:** Radiation therapy significantly reduces the risk of local recurrence and improves long-term survival in patients undergoing breast-conserving surgery.<br>• **Risks and Side Effects:** Potential side effects include skin irritation, changes in breast appearance, and, less commonly, effects on underlying tissues. These should be discussed thoroughly with the patient.<br>5. **Follow-Up Care:**<br>• **Post-Radiation:** Regular follow-up is essential to monitor for potential late effects of radiation and to ensure that any recurrence or new issues are addressed promptly.<br><br>**Conclusion:** The standard recommendation would be to proceed with whole-breast radiation therapy, potentially including a boost to the tumor bed, given the size of the tumor and high Ki-67 proliferation index. Tailoring the radiation plan to the patient's specific needs and discussing the potential benefits and side effects are crucial for optimal care. |

<table>
<tr>
<td>2<br><br>Accurate; whole breast with boost is recommended</td>
<td>For a 40-year-old woman with a diagnosis of invasive ductal carcinoma, ER/PR positive, HER2 negative, post-lumpectomy and sentinel lymph node dissection with the final pathology demonstrating a 1.3 cm well-differentiated carcinoma, negative lymphovascular invasion, clear margins by >2mm, and a Ki-67 index of 50%, the recommendations for adjuvant radiation therapy would generally include the following:<br><br>
1. Adjuvant Radiation Therapy: Radiation therapy is recommended to reduce the risk of local recurrence following lumpectomy. For early-stage breast cancer with the characteristics you've provided, whole breast irradiation is typically advised.<br>
2. Radiation Field and Dosage:<br>
• Whole-Breast Irradiation (WBI): Generally administered for about 6-7 weeks (5 days a week) with a total dose of approximately 45-50.4 Gy.<br>
• Boost: Depending on specific factors like the patient's age, tumor characteristics, and institutional protocols, a boost to the lumpectomy site (typically an additional 10-16 Gy) may be considered to further reduce the risk of local recurrence, particularly if the tumor is close to the margins.<br>
3. Treatment Planning: The exact details of the radiation plan, including field size, technique (e.g., 3D conformal, IMRT), and any additional boosts, should be tailored to the patient's anatomy and specific needs. The goal is to maximize dose to the tumor while minimizing exposure to surrounding healthy tissue, including the heart and lungs.<br>
4. Considerations:<br>
• Age and Tumor Characteristics: The patient is relatively young and has a high Ki-67 index, which might warrant a more aggressive approach in terms of radiation therapy and possibly systemic therapy.<br>
• Hormone Receptor-Positive Status: Adjuvant endocrine therapy (e.g., tamoxifen or aromatase inhibitors) will also be part of the treatment plan to reduce the risk of recurrence due to the ER/PR positive status.<br>
5. Follow-Up and Monitoring: Regular follow-up visits to monitor for potential side effects and to assess the effectiveness of the treatment will be necessary. This usually includes physical exams, mammograms, and possibly other imaging studies as indicated.<br><br>
Consultation with a radiation oncologist would provide the most individualized plan, taking into account the specifics of the case and any patient-specific considerations.</td>
</tr>
</table>

| | |
|---|---|
| 3<br><br>Accurate; whole breast with boost is recommended | Given the details provided for this 40-year-old woman with invasive ductal carcinoma, here are the recommendations for adjuvant radiation therapy:<br>1. **Radiation Therapy to the Breast:**<br>• **Indication:** Adjuvant radiation therapy is typically recommended following lumpectomy for invasive breast cancer to reduce the risk of local recurrence.<br>• **Dose:** The standard regimen is usually 45-50.4 Gy delivered in daily fractions of 1.8-2.0 Gy over 3-5 weeks.<br>2. **Radiation Therapy to the Regional Lymph Nodes:**<br>• Given that all lymph nodes are negative, regional nodal irradiation may not be necessary. However, considering the high Ki-67 (50%), which indicates a higher proliferation rate and potentially more aggressive disease, a discussion about the potential benefit of regional nodal irradiation should be considered, especially if there are additional risk factors or if the patient is at higher risk for regional recurrence.<br>• **Standard Practice:** For most cases with no lymph node involvement and negative sentinel nodes, radiation to the regional nodes is not typically recommended. However, this decision can be influenced by clinical trials, patient preferences, and discussions with the multidisciplinary team.<br>3. **Boost to the Tumor Bed:**<br>• **Indication:** A boost to the lumpectomy cavity may be considered to further reduce the risk of recurrence in the area where the cancer was removed.<br>• **Dose:** The boost is often 10-16 Gy given in daily fractions, either as part of the initial treatment or sequentially.<br>4. **Overall Considerations:**<br>• The final pathology shows well-differentiated carcinoma with negative margins and no lymphovascular invasion, which is a favorable prognosis factor.<br>• The high Ki-67 index suggests a more aggressive tumor, which may influence the decision to add additional treatments or closer follow-up.<br><br>In summary, adjuvant radiation therapy to the breast is recommended. Whether regional nodal irradiation or a boost to the tumor bed is indicated should be discussed based on the patient's overall risk profile and in consultation with a multidisciplinary team including oncology, radiation oncology, and the patient's preferences. |

| | |
|---|---|
| 4<br><br>Accurate; whole breast with boost is recommended | Given the details provided, here are the recommendations for adjuvant radiation therapy for the patient with invasive ductal carcinoma (IDC) based on current guidelines and practices:<br>1. **Indication for Radiation Therapy:**<br>• **Radiation Therapy:** Adjuvant radiation therapy is typically recommended for all patients who have undergone a lumpectomy for early-stage breast cancer. This is to reduce the risk of local recurrence.<br>• **Tumor Characteristics:** The patient's tumor is 1.3 cm, well-differentiated, and has negative margins, which aligns with criteria for radiation therapy.<br>2. **Radiation Treatment Regimen:**<br>• **Whole Breast Irradiation (WBI):** Standard practice involves whole breast irradiation after lumpectomy. The common regimen is 50 Gy delivered in 25 fractions over 5 weeks.<br>• **Boost Therapy:** A boost to the lumpectomy cavity is often considered to further reduce local recurrence risk, especially if there are additional concerns about margin involvement or higher risk features. In this case, with negative margins and a well-differentiated tumor, a boost may be considered based on institutional protocols and the patient's overall risk profile. The boost dose is usually 10-16 Gy in 5-8 fractions.<br>3. **Patient-Specific Factors:**<br>• **Age:** At 40 years old, the patient is relatively young, which might make her more amenable to adjuvant therapy recommendations given her higher risk of recurrence compared to older patients.<br>• **Hormone Receptor Status:** ER/PR positive status generally indicates a better prognosis and could influence the consideration for hormonal therapy, which is a separate treatment from radiation but important for overall management.<br>4. **Radiation Technique:**<br>• **Technique:** Conformal radiation therapy techniques such as 3D conformal radiation therapy (3D-CRT) or intensity-modulated radiation therapy (IMRT) are commonly used to deliver the radiation while sparing surrounding healthy tissues.<br>5. **Follow-Up and Monitoring:**<br>• **Surveillance:** Regular follow-up with clinical exams and imaging as indicated to monitor for any signs of recurrence or treatment-related side effects is important.<br><br>The exact details and protocols may vary depending on the specific treatment center and any additional patient-specific factors. Discussing these recommendations with the radiation oncologist will help tailor the treatment plan to the patient's individual needs and preferences. |

| | |
|---|---|
| 5<br><br>Accurate; whole breast with boost is recommended | Given the details of your diagnosis and treatment so far, here's a summary of the recommended adjuvant radiation therapy considerations:<br>1. **Tumor Size and Margins:** Your tumor is 1.3 cm and has well-defined margins greater than 2 mm. This is a positive indicator for localized treatment, as clear margins reduce the likelihood of residual disease at the edges.<br>2. **Lymph Nodes:** All sentinel lymph nodes were negative, which is encouraging as it suggests no regional spread of cancer.<br>3. **ER/PR Positive, HER2 Negative:** Your cancer is estrogen receptor (ER) and progesterone receptor (PR) positive, which means hormonal therapy (such as tamoxifen or an aromatase inhibitor) will likely be part of your treatment regimen. HER2 negative status also influences your treatment options but doesn't directly impact radiation decisions.<br>4. **Ki-67:** A Ki-67 of 50% is relatively high, indicating a higher proliferation rate. This can be a factor in considering the intensity and duration of additional treatments, including radiation.<br><br>**Adjuvant Radiation Therapy Recommendations:**<br>• **Whole-Breast Radiation Therapy (WBRT):** Typically recommended for invasive ductal carcinoma of this size and characteristics. The standard regimen usually involves 3-6 weeks of radiation therapy, often delivered in daily fractions.<br>• **Boost to Tumor Bed:** Given the size of the tumor and the fact that it's ER/PR positive with a relatively high Ki-67, a boost to the tumor bed (the area where the tumor was removed) might be recommended. This additional radiation helps target any potential residual cancer cells in that area.<br>• **Treatment Area:** Radiation usually includes the whole breast and the tumor bed, and sometimes the regional lymph nodes, depending on other factors like the potential risk of lymph node involvement (even though your sentinel nodes are negative, this is a precautionary measure in some cases).<br>• **Technique:** Techniques such as 3D conformal radiation therapy (3D-CRT) or intensity-modulated radiation therapy (IMRT) might be used to better target the tumor bed while sparing surrounding healthy tissues.<br><br>**Considerations:**<br>• **Hormonal Therapy:** Ensure you're also discussing and starting adjuvant hormonal therapy with your oncologist, as it's an important part of treatment for ER/PR positive cancers.<br>• **Follow-Up:** Regular follow-up appointments will be crucial to monitor for any side effects of radiation and to ensure the effectiveness of the overall treatment plan.<br><br>Discuss these recommendations in detail with your radiation oncologist, as they can provide a tailored treatment plan based on your individual case and preferences. |

Second Example (ChatGPT 4o-Mini)

2. 40 year old woman with new diagnosis of invasive ductal carcinoma, ER/PR positive, HER2 negative status post modified radical mastectomy and axillary lymph node dissection, final pathology demonstrating 1.3cm carcinoma, well differentiated, no lymphovascular invasion, margins negative by >2mm, Ki-67 50%, and five of ten lymph nodes positive for macrometastases. Presenting for adjuvant radiation recommendations:
**Expected response: Regional Nodal Irradiation**

| General Observations |
| --- |
| - |

| | |
| --- | --- |
| 1<br><br>Accurate; regional nodal irradiation is recommended | For a 40-year-old woman with the described characteristics following a modified radical mastectomy and axillary lymph node dissection, here are the general recommendations for adjuvant radiation therapy:<br><br>**1. Breast Radiation:**<br>• **Indication:** Given the presence of invasive ductal carcinoma, the size of the tumor (1.3 cm), and the fact that she had a mastectomy, radiation therapy to the chest wall is typically recommended. This helps to reduce the risk of local recurrence.<br>• **Technique:** Whole-breast irradiation (WBI) is generally used if any breast tissue remains or if there's a concern for local recurrence in the residual breast tissue. In the case of a mastectomy, chest wall radiation is used. The usual dose is 50 Gy in 25 fractions over 5 weeks.<br><br>**2. Regional Nodal Radiation:**<br>• **Indication:** With five out of ten axillary lymph nodes positive for macrometastases, regional nodal irradiation is recommended. This approach targets the axilla, supraclavicular, and potentially the internal mammary nodes, as there is a higher risk of residual or recurrent disease in these regions.<br>• **Technique:** Regional nodal radiation typically involves doses of 45-50.4 Gy to the axilla and supraclavicular areas, with additional considerations for internal mammary nodes depending on the exact nodal involvement and the patient's specific anatomy.<br><br>**3. Special Considerations:**<br>• **Ki-67 Index:** A high Ki-67 index (50%) suggests a high proliferation rate, which is associated with a more aggressive tumor. This supports the need for thorough adjuvant therapy to minimize the risk of recurrence.<br>• **Margins and Lymphovascular Invasion:** The fact that margins are negative by >2mm and there is no lymphovascular invasion are favorable prognostic factors, which is positive, but the number of involved nodes still indicates a need for adjuvant therapy.<br><br>**4. Treatment Planning:**<br>• **Simulation and Planning:** Proper simulation is essential to ensure accurate delivery of radiation therapy while minimizing exposure to surrounding healthy tissues. This often includes imaging and contouring to plan the precise areas to be irradiated.<br>• **Follow-Up:** Regular follow-up visits are necessary to monitor for side effects and assess the response to treatment.<br><br>Consultation with a radiation oncologist is crucial to tailor the treatment plan to the individual patient's needs, considering all aspects of her clinical scenario and preferences. |

| | |
|---|---|
| 2<br><br>Accurate; regional nodal irradiation is recommended | Given the details of the case—40-year-old woman with invasive ductal carcinoma, ER/PR positive, HER2 negative, with a final pathology report indicating a well-differentiated 1.3 cm carcinoma, negative margins, a high Ki-67 index of 50%, and five of ten lymph nodes positive for macrometastases—here are recommendations for adjuvant radiation therapy:<br>1. **Chest Wall Radiation:** Post-mastectomy radiation is typically indicated due to the presence of five positive lymph nodes. This helps to reduce the risk of local recurrence. The radiation should cover the entire chest wall, including the supraclavicular and axillary regions.<br>2. **Regional Nodal Irradiation:** Considering the involvement of multiple lymph nodes, regional nodal irradiation should be part of the treatment plan. This usually includes the supraclavicular and axillary nodes.<br>3. **Radiation Dose:** A standard dose regimen is often 50 Gy in 25 fractions to the chest wall and regional nodes, followed by a boost to the tumor bed if applicable (usually 10-16 Gy in 5-8 fractions). However, the boost is less common in modern practice, particularly if the margins are negative and there is no evidence of residual disease in the resection margins.<br>4. **Treatment Planning:** Ensure careful planning to minimize radiation exposure to the surrounding healthy tissues, especially the heart and lungs. Advanced techniques such as intensity-modulated radiation therapy (IMRT) or volumetric modulated arc therapy (VMAT) may be used to optimize the dose distribution and reduce side effects.<br>5. **Consideration of Adjuvant Therapy:** Given the high Ki-67 index (50%), which indicates a high proliferation rate, consider whether additional systemic adjuvant therapy (such as chemotherapy) is indicated. This decision would depend on a detailed review of the patient's overall risk profile and potentially the use of gene expression assays or other predictive markers.<br><br>**Follow-Up:** Regular follow-up is essential to monitor for any potential side effects of radiation therapy and to evaluate for signs of recurrence. This typically includes physical examinations, imaging studies, and possibly additional markers.<br><br>It's crucial to work closely with a multidisciplinary team, including a radiation oncologist, medical oncologist, and surgical oncologist, to tailor the treatment plan to the patient's specific needs and preferences. |

<table>
<tr>
<td>3<br><br>Accurate; regional nodal irradiation is recommended</td>
<td>

Given the diagnosis and treatment history, here's a summary of the considerations for adjuvant radiation therapy:

1. **Type of Cancer and Initial Treatment:**
   - **Invasive Ductal Carcinoma:** ER/PR positive, HER2 negative.
   - **Post-Surgical Status:** Modified radical mastectomy with axillary lymph node dissection. Final pathology shows a 1.3 cm carcinoma, well differentiated, no lymphovascular invasion, margins >2mm, Ki-67 50%, and five of ten lymph nodes positive for macrometastases.
2. **Radiation Therapy Recommendations:**
   - **Chest Wall Radiation:** Given the size of the tumor (1.3 cm) and the fact that the patient underwent a mastectomy, radiation to the chest wall is generally recommended. This helps to reduce the risk of local recurrence. The typical radiation dose is around 50 Gy in 25 fractions over 5 weeks.
   - **Regional Nodal Radiation:** Since there are five positive lymph nodes, radiation to the regional lymph nodes is usually recommended. This may include the supraclavicular, axillary, and internal mammary nodes. This approach is intended to address any residual microscopic disease and improve outcomes. The dose can vary, but a common regimen is around 45-50 Gy to the regional nodes, often delivered in conjunction with chest wall radiation.
3. **Rationale for Radiation:**
   - **Local Control:** Radiation therapy aims to reduce the risk of recurrence in the chest wall and nearby regions.
   - **Lymph Node Involvement:** With five positive lymph nodes, there is a higher risk of residual disease in the regional lymph nodes, which justifies regional nodal irradiation.
4. **Additional Considerations:**
   - **Systemic Therapy:** Given the high Ki-67 (50%), the patient may benefit from adjuvant chemotherapy in addition to radiation and hormonal therapy.
   - **Hormonal Therapy:** Since the cancer is ER/PR positive, endocrine therapy (e.g., tamoxifen or aromatase inhibitors) will be a crucial component of the adjuvant treatment plan.
   - **Follow-up and Surveillance:** Regular follow-up with imaging and clinical evaluations to monitor for any recurrence or late effects of treatment.

These recommendations should be tailored based on the patient's overall health, preferences, and any additional clinical factors. It's important for the patient to discuss these options with their oncology team to make an informed decision based on their specific situation.

</td>
</tr>
</table>

| | |
|---|---|
| 4<br><br>Accurate; regional nodal irradiation is recommended | For a 40-year-old woman with invasive ductal carcinoma (IDC) who has undergone a modified radical mastectomy and axillary lymph node dissection with the following pathology results, the recommended adjuvant radiation therapy (RT) approach generally includes the following considerations:<br><br>1. **Radiation to the Chest Wall:** Given that she has undergone a modified radical mastectomy, adjuvant radiation to the chest wall is typically recommended to reduce the risk of local recurrence. This is especially important given the size of the tumor (1.3 cm), the positive lymph nodes (5 of 10 with macrometastases), and the high Ki-67 proliferation index (50%), which suggests a more aggressive tumor biology.<br>2. **Regional Nodal Irradiation:** Considering that 5 of 10 axillary lymph nodes are positive for macrometastases, regional nodal irradiation (RNI) is often advised. This includes:<br>• **Axillary Nodal Irradiation:** Direct radiation to the axilla.<br>• **Supraclavicular Nodal Irradiation:** Radiation to the supraclavicular region is generally recommended due to the positive lymph nodes, as it helps address potential microscopic disease in the supraclavicular lymph nodes.<br>• **Internal Mammary Nodal Irradiation:** Depending on institutional protocols and risk assessment, internal mammary nodal irradiation might be considered, especially if there are additional risk factors or if the patient is at higher risk for internal mammary node involvement.<br>3. **Dose and Fractionation:** Standard radiation therapy for the chest wall typically involves a dose of around 45-50.4 Gy in daily fractions of 1.8-2.0 Gy over approximately 5-6 weeks. For regional nodal irradiation, the dose might be similar, with adjustments based on the specific regional areas treated.<br>4. **Radiation Planning and Technique:** Intensity-modulated radiation therapy (IMRT) or 3D conformal radiation therapy may be employed to improve target coverage and minimize dose to surrounding healthy tissue. A careful planning process is essential to optimize the therapeutic ratio, especially given the involvement of regional nodes.<br>5. **Patient-Specific Considerations:** Discussions with the radiation oncologist will also take into account any patient-specific factors such as overall health, prior treatments, and personal preferences. For example, considerations regarding potential side effects or the patient's baseline health status may influence the final radiation plan.<br><br>In summary, for this patient, adjuvant radiation therapy would likely include chest wall irradiation, regional nodal irradiation (including axillary and supraclavicular regions), and potentially internal mammary nodes based on further evaluation. The exact regimen and approach would be tailored to her specific case in consultation with her radiation oncology team. |

| | |
|---|---|
| 5<br><br>Accurate; regional nodal irradiation is recommended | For a 40-year-old woman with invasive ductal carcinoma who has undergone a modified radical mastectomy and axillary lymph node dissection, the adjuvant radiation therapy recommendations would typically be based on the following considerations:<br>1. **Radiation to the Chest Wall:** Given the presence of invasive ductal carcinoma with lymph node involvement, radiation therapy to the chest wall is recommended. The goal is to reduce the risk of local recurrence. The standard approach is usually a dose of 45-50.4 Gy to the chest wall, typically delivered in 1.8-2.0 Gy fractions.<br>2. **Axillary Radiation:** Since the patient had five of ten axillary lymph nodes positive for macrometastases, axillary radiation may be considered to reduce the risk of regional recurrence. This is especially relevant given the higher nodal involvement. Axillary radiation typically targets the axillary lymph nodes as well as the supraclavicular and possibly the infraclavicular regions, depending on the extent of nodal involvement.<br>3. **Regional Nodal Irradiation:** In cases with multiple positive lymph nodes, regional nodal irradiation is often advised. This includes treating the supraclavicular nodes, and sometimes the infraclavicular nodes, in addition to the chest wall. The total dose for regional nodes often ranges from 45-50.4 Gy, delivered in daily fractions.<br>4. **Radiation Therapy Planning:** It's crucial that the radiation therapy plan is tailored to minimize exposure to healthy tissues, particularly the heart and lungs, while effectively targeting the cancerous areas. Modern techniques such as intensity-modulated radiation therapy (IMRT) or three-dimensional conformal radiation therapy (3D-CRT) may be used to achieve this.<br>5. **Adjuvant Therapy Considerations:** The high Ki-67 index (50%) suggests a high proliferation rate, which could indicate a more aggressive tumor. This, along with the node-positive status, supports the use of adjuvant radiation to address potential residual disease.<br>6. **Follow-up and Monitoring:** Post-radiation therapy, the patient will need regular follow-ups to monitor for any signs of recurrence and to manage any potential side effects from the treatment.<br><br>It's essential that the specifics of the radiation therapy plan are determined by a multidisciplinary team, including an oncologist, radiation therapist, and surgeon, considering the individual patient's situation and preferences. |

Third Example (ChatGPT 4o-Mini)

3. 80 year old woman with new diagnosis of invasive ductal carcinoma, ER/PR positive, HER2 negative status post modified radical mastectomy and axillary lymph node dissection, final pathology demonstrating 1.3cm carcinoma, well differentiated, no lymphovascular invasion, margins negative by >2mm, Ki-67 50%, and zero of ten lymph nodes positive for macrometastases. Presenting for adjuvant radiation recommendations: **Expected response: No radiation**

| |
|---|
| General Observations<br><br>- |

| |
|---|
| Phrasing 1:<br><br>Phrasing 2:<br><br>Phrasing 3:<br><br>Chain of Reasoning Prompting: |

| | |
|---|---|
| 1<br><br>Inaccurate; omitting adjuvant radiation is not recommended | For an 80-year-old woman with a new diagnosis of invasive ductal carcinoma, ER/PR positive, HER2 negative, who has undergone a modified radical mastectomy with negative margins and no lymph node involvement, here are some considerations for adjuvant radiation therapy:<br><br>1. **Indication for Adjuvant Radiation Therapy:**<br>• Given that the patient had a modified radical mastectomy, radiation therapy is generally recommended to reduce the risk of local recurrence. This is especially pertinent in cases with high-risk features or when the primary tumor is ≥1 cm in size.<br>2. **Tumor Characteristics:**<br>• **Size:** The tumor is 1.3 cm, which qualifies it for consideration of radiation therapy.<br>• **Grade:** The carcinoma is well-differentiated, which typically suggests a lower risk, but radiation is still often recommended.<br>• **Ki-67:** The Ki-67 proliferation index of 50% is relatively high, indicating a higher risk of recurrence which supports the use of radiation therapy.<br>3. **Radiation Therapy Recommendations:**<br>• **Radiation to the Chest Wall:** Typically, adjuvant radiation therapy is directed to the chest wall in patients who have undergone a mastectomy. This is aimed at reducing the risk of local recurrence in the breast area.<br>• **Dose and Schedule:** Standard treatment involves delivering approximately 50 Gy (Gray) in 25 fractions over 5 weeks. Adjustments may be made based on individual patient factors and specific clinical protocols.<br>• **Regional Nodes:** Given the absence of positive lymph nodes, routine radiation to the regional nodes is generally not indicated unless there are other high-risk features or clinical trials suggest otherwise.<br>4. **Patient-Specific Considerations:**<br>• **Age and Comorbidities:** At 80 years old, it's important to consider the patient's overall health, functional status, and comorbidities. If the patient has significant comorbidities or functional limitations, these factors might influence the decision-making process. However, age alone should not preclude her from receiving radiation therapy if it is deemed beneficial.<br>• **Patient Preferences:** Discussing the benefits and potential side effects of radiation therapy with the patient is essential. It's also important to consider her personal preferences and quality of life.<br><br>In summary, based on the provided clinical details, adjuvant radiation therapy to the chest wall is recommended to reduce the risk of local recurrence. The treatment regimen typically involves 50 Gy delivered in 25 fractions over 5 weeks. The patient's age and overall health should be considered in the treatment planning process, but radiation therapy remains a standard recommendation for this scenario. |

| | |
|---|---|
| 2<br><br>Inaccurate; omitting adjuvant radiation is not recommended | Given the details of the case, here are some recommendations for adjuvant radiation therapy:<br>1. **Target Area for Radiation:**<br>• **Chest Wall:** Since the patient has had a modified radical mastectomy, radiation to the chest wall is typically recommended to reduce the risk of local recurrence. This is especially pertinent in cases with higher risk features or where there's a concern about residual microscopic disease.<br>• **Regional Lymph Nodes:** While the patient's axillary lymph nodes were negative for macrometastases, radiation to the regional lymph nodes (axillary, supraclavicular, and internal mammary nodes) can be considered based on the patient's overall risk profile. For patients with negative nodes and well-differentiated tumors, the decision to include regional nodes can be individualized based on additional risk factors and patient preferences.<br>2. **Radiation Dose and Schedule:**<br>• **Chest Wall Radiation:** Typically, the dose is 45-50.4 Gy in 1.8-2.0 Gy fractions. The specific dose can be adjusted based on institutional protocols and the patient's tolerance.<br>• **Regional Lymph Nodes:** If included, the dose is usually similar to that of the chest wall but may be adjusted based on the target volumes and potential overlap.<br>3. **Considerations for Radiation Therapy:**<br>• **Age and Health:** Given the patient's age, it's important to carefully weigh the benefits of radiation therapy against potential risks and side effects. In some cases, older patients may have a lower threshold for more intensive treatments if they have other significant health issues.<br>• **Ki-67 Index:** A high Ki-67 index (50% in this case) indicates a higher proliferative rate and can be a factor in considering adjuvant treatments. High Ki-67 might influence the decision to provide radiation to reduce the risk of recurrence, though this is more commonly considered in the context of chemotherapy rather than radiation therapy alone.<br>4. **Patient Preferences and Quality of Life:** Discuss the potential benefits and side effects of radiation therapy with the patient. Involvement in the decision-making process is crucial, especially in older patients where quality of life considerations are significant.<br><br>In summary, adjuvant radiation therapy to the chest wall is typically recommended for this patient, and consideration of regional lymph node irradiation might be discussed based on additional risk factors. The specific plan should be tailored to the patient's overall health, preferences, and the potential benefits of treatment. |

| | |
|---|---|
| 3<br><br>Inaccurate; omitting adjuvant radiation is not recommended | 2. **Considerations for Radiation Therapy:**<br>• **Tumor Size and Grade:** While the tumor is small and well-differentiated, the high Ki-67 index suggests a more aggressive nature, which could justify adjuvant radiation.<br>• **Margins:** Negative margins by >2mm are reassuring but not always definitive in avoiding recurrence.<br>• **Age and Overall Health:** At 80 years old, the patient's overall health and potential for treatment-related side effects must be considered. Adjuvant radiation therapy should balance the benefits in reducing recurrence with potential risks.<br><br>**Specific Recommendations:**<br>• **Radiation Therapy to Chest Wall:** Given the tumor size and high Ki-67 index, radiation to the chest wall is typically recommended to reduce the risk of local recurrence.<br>• **Axillary Radiation:** Since all lymph nodes were negative for macrometastases, radiation to the axilla is usually not required. However, this may vary based on additional risk factors or institutional protocols.<br><br>**Final Decision:**<br>Given the patient's tumor characteristics and the potential benefits of radiation therapy in reducing the risk of local recurrence, radiation therapy to the chest wall should be strongly considered. The decision will ultimately depend on a discussion with the patient about the potential benefits and risks, as well as her overall health and preferences.<br><br>In summary, for this patient, adjuvant radiation therapy to the chest wall is generally recommended based on the high Ki-67 index and small but significant tumor size. A detailed discussion about the benefits and risks of radiation therapy, considering her age and health, should guide the final treatment plan. |

| | |
|---|---|
| 4<br><br>Inaccurate: omitting adjuvant radiation is not recommended | Given the details of the patient's diagnosis and treatment, here's a summary of considerations for adjuvant radiation therapy:<br><br>1. **Stage and Prognosis:**<br>• The patient has invasive ductal carcinoma (IDC) with a well-differentiated tumor (grade 1), measuring 1.3 cm, which is a favorable characteristic.<br>• Lymphovascular invasion is absent, and all ten axillary lymph nodes are negative for macrometastases, indicating that the disease has not spread to the lymph nodes.<br>2. **Margin Status:**<br>• Surgical margins are negative by more than 2 mm, which suggests that the tumor was adequately removed.<br>3. **Ki-67 Index:**<br>• The Ki-67 index of 50% indicates a higher proliferation rate, which can be associated with a higher risk of recurrence, even though other factors are favorable.<br>4. **ER/PR and HER2 Status:**<br>• The tumor is ER/PR positive and HER2 negative, which typically suggests a more favorable prognosis and responsiveness to endocrine therapy.<br>5. **Adjuvant Radiation Therapy Considerations:**<br>• For early-stage, ER-positive, HER2-negative breast cancer, radiation therapy is generally recommended if the patient underwent a breast-conserving surgery (lumpectomy). However, since the patient had a modified radical mastectomy, the primary considerations for radiation therapy include:<br> • **Local Recurrence Risk:** Even with a mastectomy, radiation may be considered if there are high-risk features or if there is concern about local recurrence.<br> • **Proliferation Rate:** The high Ki-67 index (50%) suggests a more aggressive tumor, potentially increasing the risk of local recurrence.<br>6. **Recommendations:**<br>• **Radiation to Chest Wall:** Given the tumor's size, the absence of lymph node involvement, and the high Ki-67 index, adjuvant radiation to the chest wall is generally recommended to reduce the risk of local recurrence.<br>• **Regional Nodes:** Considering the lack of lymph node involvement, routine radiation to the axilla or supraclavicular nodes may not be necessary unless there were other high-risk features or findings.<br>7. **Patient-Specific Factors:**<br>• **Age:** At 80 years old, the patient's overall health and ability to tolerate radiation should be assessed. For older patients, radiation therapy may need to be carefully weighed against the potential benefits, considering the patient's life expectancy and quality of life.<br>• **Endocrine Therapy:** Given the ER-positive status, endocrine therapy (such as an aromatase inhibitor or tamoxifen) should be a part of the adjuvant treatment plan to address any residual risk of recurrence.<br><br>**Summary:** For this patient, adjuvant radiation therapy to the chest wall is typically recommended due to the high Ki-67 index and the need to minimize the risk of local recurrence. The final decision should consider the patient's overall health, preferences, and specific clinical context. It's also important to coordinate with the oncology team to ensure a comprehensive approach that includes endocrine therapy and any additional supportive care as needed. |

<table>
<tr>
<td>5<br><br>Inaccurate; omitting adjuvant radiation is not recommended</td>
<td>
Given the patient's profile and the final pathology results, here's a detailed approach to adjuvant radiation recommendations:
<ol>
<li>Tumor Characteristics:
<ul>
<li>Size: 1.3 cm, which is less than 2 cm.</li>
<li>Grade: Well differentiated.</li>
<li>Lymphovascular Invasion: None.</li>
<li>Margins: Negative by >2 mm.</li>
<li>Ki-67: 50% (indicates a high proliferation rate, which can influence the decision for adjuvant therapy).</li>
<li>Lymph Nodes: Zero of ten nodes positive for macrometastases.</li>
</ul>
</li>
<li>General Radiation Therapy Recommendations:
<ul>
<li>Whole Breast Radiation Therapy (WBRT): For early-stage, ER/PR positive, HER2 negative breast cancer with the characteristics described, adjuvant whole breast radiation therapy is typically recommended. This reduces the risk of local recurrence.</li>
<li>Post-Mastectomy Radiation Therapy (PMRT): Given the patient's age and well-differentiated cancer with negative margins, the use of post-mastectomy radiation therapy is generally considered based on a few factors:
<ul>
<li>Size of the Tumor: While PMRT is often considered for tumors greater than 5 cm or with involved lymph nodes, this patient has a small, well-differentiated tumor with no positive nodes, which might lower the urgency of PMRT.</li>
<li>Lymph Node Involvement: The absence of positive lymph nodes is favorable but, in some guidelines, PMRT may still be considered based on other risk factors or institutional practices.</li>
</ul>
</li>
<li>Boost Radiation: A radiation boost to the tumor bed might be considered, especially if the patient had a close or positive margin. However, with margins >2 mm and a small, well-differentiated tumor, a boost might not be necessary unless there are additional risk factors.</li>
</ul>
</li>
<li>Personalized Considerations:
<ul>
<li>Age: Being 80 years old, the patient's overall health, life expectancy, and preferences should be considered. Older patients often have different risk-benefit profiles for radiation therapy.</li>
<li>Patient Preference: The decision should involve a discussion with the patient regarding the benefits and potential side effects of radiation therapy.</li>
</ul>
</li>
<li>Consultation and Final Decision:
<ul>
<li>Multidisciplinary Team: A thorough discussion with a multidisciplinary team including oncologists, radiologists, and the patient's primary care provider will help tailor the radiation therapy plan to the individual's specific case and preferences.</li>
</ul>
</li>
</ol>
In summary, for this patient, whole breast radiation therapy is typically recommended based on her cancer characteristics and the aim to reduce the risk of local recurrence. Post-mastectomy radiation therapy might be considered, but given her specific tumor characteristics and age, this decision should be individualized and discussed thoroughly with the patient.
</td>
</tr>
</table>
